# The Association of Stay-at-Home Orders and the Spread of COVID-19 in Rural and Urban United States

**DOI:** 10.1101/2021.07.13.21260476

**Authors:** David H. Jiang, Darius J. Roy, Benjamin D. Pollock, Nilay D. Shah, Rozalina G. McCoy

**Affiliations:** Division of Health Care Delivery Research, Robert D. and Patricia E. Kern Center for the Science of Health Care Delivery, Mayo Clinic; Rochester, MN; Division of Cardiovascular Medicine, Brigham and Women’s Hospital; Boston, MA; Department of Quality, Experience, and Affordability, Mayo Clinic; Rochester, MN; Division of Community Internal Medicine, Department of Medicine, Mayo Clinic; Rochester, MN

**Keywords:** Stay at home, COVID-19, urban-rural, disparities, health policy

## Abstract

**Background:** Throughout the spring of 2020, stay-at-home orders were imposed to curb the spread of COVID-19. There is limited data on the effectiveness of stay-at-home orders, particularly in rural as compared to urban areas.

**Objective:** To examine the association between stay-at-home order implementation and the incidence of COVID-19 in rural vs. urban counties.

**Design:** Interrupted time series analysis using a mixed effects zero-inflated Poisson model.

**Participants:** 3,142 U.S. counties.

**Interventions:** Stay-at-home orders.

**Main Measures:** COVID-19 daily incidence (primary) and mobility (secondary and intermediate measure of stay-at-home effectiveness)

**Key Results:** Stay-at-home orders were implemented later (median March 30 vs. March 28) and were shorter (median 35 vs. 54 days) in rural than urban counties. Indoor mobility was, on average, 2.6-6.9% higher in rural than urban counties both during and after stay-at-home orders. Compared to the baseline (pre-stay-at-home) period, the number of new COVID-19 cases increased under stay-at-home by IRR 1.60 (95% CI, 1.57-1.64) in rural and 1.36 (95% CI, 1.30-1.42) in urban counties. For each day under stay-at-home orders, the number of new cases changed by a factor of 0.982 (95% CI 0.981-0.982) in rural and 0.952 (95% CI, 0.951-0.953) in urban counties compared to prior to stay-at-home. Each day after stay-at-home orders expired, the number of new cases changed by a factor of 0.995 (95% CI, 0.994-0.995) in rural and 0.997 (95% CI, 0.995-0.999) in urban counties compared to prior to stay-at-home.

**Conclusion:** Stay-at-home orders decreased mobility and slowed the spread of COVID-19, but less effectively in rural than in urban counties. This necessitates a critical reevaluation of how stay-at-home orders are designed, communicated, and implemented in rural areas.

## Introduction

As cases of coronavirus disease-2019 (COVID-19) emerged in the United States in early 2020, pressure grew on federal, state, and local governments to implement public health guidelines aimed at reducing its impact on the healthcare system and society. These measures included school and non-essential business closures, bans on social gatherings, and stay-at-home orders. However, the U.S. is not monolithic, and in the absence of a unified federal strategy,^1^ the timing, duration, stringency, and enforcement of stay-at-home orders varied widely across municipalities.^2^ Nearly one-fifth of the U.S. population resides in rural areas.^3^ Resource availability, political affiliation, and attitudes toward COVID-19 all differ between rural and urban areas,^4^ and, as such, public health guidance and implementation may need to be adapted to the unique needs and circumstances of each setting. Thus, while emerging evidence has demonstrated an overall effectiveness of physical distancing measures as enforced by stay-at-home orders,^5-11^ potential differences in their effectiveness and durability (i.e., continued effectiveness even after the stay-at-home orders are lifted) of stay-at-home orders between rural and urban areas have not been explored. Identification of such differences would call for closer examination of barriers to optimal implementation and effectiveness with the goal of developing more effective, adaptive, and scalable public health guidelines going forward. With the more transmissible B.1.617.2 (Delta) variant of SARS-CoV-2 virus now the dominant strain in the U.S. and the resurgence of cases,^12^ there are discussions of new stay-at-home orders in many jurisdictions.^13^ As such, it is more important to consider the difference in efficacy of stay-at-home orders between geographical regions.

We focus on rural areas specifically because as COVID-19 first emerged in urban centers^14^ and only later spread throughout the country, rural perception of the pandemic and response to it may have differed from urban areas.^4^ As a result, rural residents may feel that public health regulations stemming from urban experiences do not equally apply to rural populations, which in turn, may have led to differences in the adoption of stay-at-home orders, the fidelity to physical distancing, and ultimately to the spread of COVID-19.

Stay-at-home orders are only effective if they translate to greater physical distancing and reduced mobility, particularly in indoor settings that pose the highest risk for COVID-19 transmission. Several studies have correlated changes in population-level mobility with COVID-19 infection rates^6 15^ and demonstrated reduced mobility with the passage of stay-at-home orders.^2 16 17^ Yet, public intent to comply with physical distancing regulations has varied widely across the U.S., influenced by religious affiliation,^18^ income,^19^ political ideology,^20^ and local/state political leadership.^21^ We hypothesize that it also differed between rural and urban areas, though this has not heretofore been explicitly assessed.

Better understanding of the spread of COVID-19 and of the effectiveness and durability of stay-at-home orders in rural areas, specifically, is important because rural populations are highly susceptible to severe manifestations of COVID-19.^22^ Rates of obesity,^23^ heart disease,^24 25^ chronic lung disease,^24^ diabetes,^25^ smoking,^26^ and multimorbidity are major risk factors for serious COVID-19 illness and are all higher in rural than urban settings.^26^ Rural residents are also older than urban residents^26^ and age is a major determinant of COVID-19 severity and mortality.^27^ Furthermore, rural areas may have lower resiliency against the pandemic, with fewer intensive care unit (ICU) beds^26^ and many small regional and critical access hospitals and clinics already facing financial hardship.^28^

Thus, in the face of these unique challenges facing rural residents, identifying opportunities to improve the effectiveness of stay-at-home orders is critical as the nation faces resurgence of case numbers and new strains of COVID-19.^29^ The recent increases in the case numbers in many states and across the nation have led policy makers to consider reimplementing or fully reinstating stay-at-home orders.^30^ In order to optimize the current iteration of stay-at-home orders and inform future infection control efforts, we examine the effectiveness of such orders in rural and urban areas of the United States. Specifically, we examine the mobility patterns and COVID-19 infection rates in rural and urban counties during and after stay-at-home orders, focusing specifically on how stay-at-home orders impacted rates of new infections and mobility in these areas.

## Methods

### Data Sources and Study Population

County-level COVID-19 case data was acquired from the Centers for Disease Control and Prevention, courtesy of USAFacts.org, a nonpartisan, nonprofit government data repository.^31^ The dataset contained cumulative county-level cases for all 3,142 U.S. counties and county-equivalents between January 22 and June 10, 2020. Each county was categorized as urban or rural using 2013 county-level Rural-Urban Continuum Codes (RUCC) per the Economic Research Service of the U.S. Department of Agriculture and the Office of Management and Budget.^32^ Urban counties were defined as having RUCC of 1-3 and rural counties as RUCC of 4-7.

Start and end dates of stay-at-home orders were identified from data compiled and published by the *New York Times*,^33 34^ which was then manually verified for completeness and accuracy by the study team via internet search and review of state executive orders. A binary variable was created to indicate each day individual counties were under stay-at-home. When stay-at-home orders were issued by state governments, it was assumed that all counties in that state fell under that order. If individual counties issued their own stay-at-home orders (e.g. Davis County, Utah^35^), county-specific start and end dates were identified and applied. If stay-at-home orders were declared by individual cities (e.g. Jackson, Wyoming,^36^ Oklahoma City, Oklahoma^37^), those stay-at-home order dates were applied to the rest of that county, as cities comprised the majority of that county’s population.

County-level mobility trends were identified from COVID-19 Community Mobility Reports between February 15 and June 14, 2020, available from Google, LLC.^38^ This data presents how the visitors to or time spent in specific categories of places change each day compared to a baseline day. The baseline day is represented by a normal value for that day of the week based on the median value from the 5-week period between January 3 and February 6, 2020.

### Outcomes

The primary outcome was the number of new cases of COVID-19 per day per 100,000 people in the county recorded 14 days from the current date. A lag of 14 days was selected based on the duration of susceptibility to a new diagnosis of COVID-19 after an initial exposure.^39 40^ Sensitivity analyses were performed for 5-day and 10-day lagged cases. Secondary outcomes were percent changes in county-level mobility for the following categories: grocery/pharmacy, retail/recreation, residential, and workplace. Transit and parks categories were not included due to insufficient data for all counties.

### Independent Variables

Independent variables included in the models were the primary exposure of county classification (rural vs. urban), stay-at-home order status (before, during, or after), days since start of follow-up (to account for time), days under stay-at-home orders, and days after stay-at-home orders.

### Statistical Analysis

Interrupted time series analysis of county-level, 14-day lagged COVID-19 daily new cases examined the impact of stay-at-home orders on rural and urban counties. Models included the independent variables outlined above and interaction terms between each of the variables and rural/urban status. To account for differences between counties, we used a mixed effects models with random intercept by county.

A variety of mixed effects count data models were compared on the basis of model diagnostics, Akaike information criterion (AIC), and parsimony: Poisson, zero-inflated Poisson, zero-inflated Poisson with random intercept and slope, negative binomial, negative binomial with random intercept and slope, and zero-inflated negative binomial. All used the same variables for the fixed effects to account for the time varying nature of stay-at-home orders and were offset by county population divided by 100,000 to standardize by population. Models were run using the glmmTMB package in R,^41^ while model diagnostics examined the simulated quantile scaled residuals using the DHARMa package in R.^42^ Models were assessed for over-dispersion, zero-inflation, and distribution of residuals. The mixed effects zero-inflated Poisson model with random intercept by county was chosen as the best model, as it was not significantly zero-inflated, not over-dispersed, did not contain outliers, and had the expected distribution of residuals. It was temporally autocorrelated by the Durbin-Watson test, but this is unavoidable (additions of variance-covariance structures led to significant over-dispersion) and did not have a significant effect on the results because of the long follow-up time, the significance of the results, and the large number of counties.^43^ Using offsets calculated from rural and urban population averages, we visualized the impact of stay-at-home orders by estimating the number of new COVID-19 cases in rural and urban counties for the start, end, and duration of stay-at-home orders. We conducted sensitivity analysis surrounding the lag time for our regression. We conducted a five-day and a ten-day lag and compared the results to that of our original 14-day lag. Detailed description of the methods is available in the Supplement.

Google Community Mobility Reports were analyzed for differences in during- and post-stay-at-home mobility between urban and rural counties using repeated measures ANOVA via the rstatix package in R.^44^ Before testing for significance, all of the mobility data were examined for outliers and normality. Outliers were classified as observations outside of 1.5 times the interquartile range (IQR) of their respective distribution (mobility type and rurality). Grocery/pharmacy and workplace were the only categories with outliers, with 8 outliers (4 days) and 2 outliers (1 day) removed for these categories, respectively.

Difference in stay-at-home duration between county type was assessed using Wilcoxon rank sum test with continuity correction.

Figure 2 was generated by inputting the estimates of fixed effects and the urban and rural averages of stay-at-home orders start and end dates. The outcome was divided by the offset to standardize the results per 100,000 population. The respective offsets for urban and rural counties were calculated using urban and rural counties respective population averages. Similarly, the extrapolations were generated by using the conditional model only with intercept and variables: Rurality, Days, and Rurality*Days. The extrapolations represent continuation of the before stay-at-home order trends.

## Results

We analyzed data for all 1,976 rural (62.9%) and 1,166 urban (37.1%) U.S. counties, home to 46,063,061 (14.0%) and 282,176,462 (86.0%) people, respectively (**Table 1**). As of June 10, 2020, there were 1,786,886 cases of COVID-19 in the U.S., of which 9.0% (N=161,452) were in rural counties. Adjusted for population, this translates to 350.5 per 100,000 persons in rural counties and 576.0 per 100,000 persons in urban counties. During the study period, 1854 (93.8%) rural counties and 1075 (92.2%) urban counties were covered by a stay-at-home order. The median start date for stay-at-home orders was March 30 for rural counties and March 28 for urban counties. The median duration of stay-at-home orders was 35 days (IQR 28-68) for rural counties and 54 days (IQR 29-70) for urban counties (p <0.001).

**Table 1.**
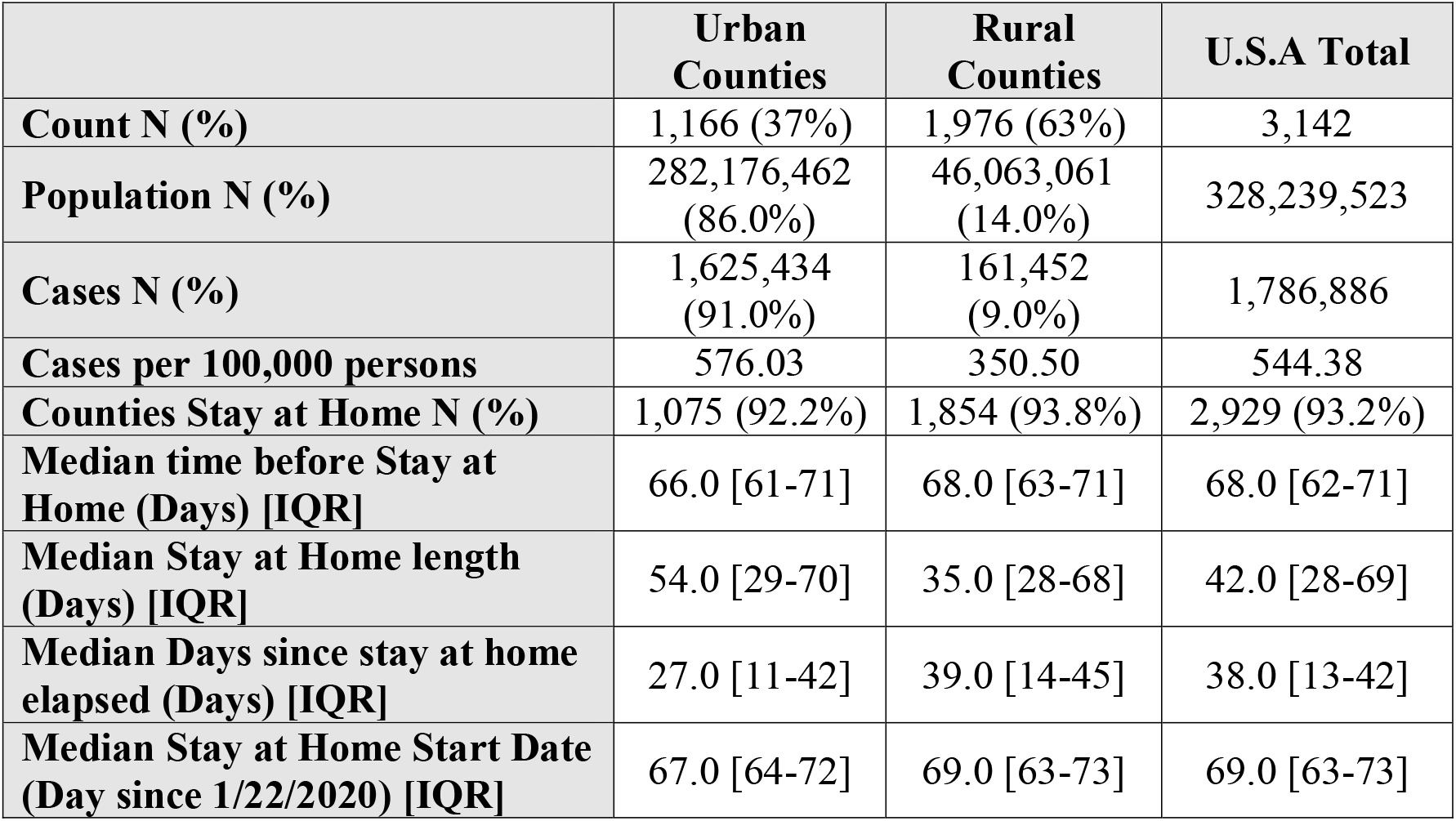
Study Cohort and Stay-at-Home Implementation.

|  | **Urban**<br>**Counties** | **Rural**<br>**Counties** | **U.S.A Total** |
| --- | --- | --- | --- |
| **Count N (%)** | 1,166 (37%) | 1,976 (63%) | 3,142 |
| **Population N (%)** | 282,176,462<br>(86.0%) | 46,063,061<br>(14.0%) | 328,239,523 |
| **Cases N (%)** | 1,625,434<br>(91.0%) | 161,452<br>(9.0%) | 1,786,886 |
| **Cases per 100,000 persons** | 576.03 | 350.50 | 544.38 |
| **Counties Stay at Home N (%)** | 1,075 (92.2%) | 1,854 (93.8%) | 2,929 (93.2%) |
| **Median time before Stay at Home (Days) [IQR]** | 66.0 [61-71] | 68.0 [63-71] | 68.0 [62-71] |
| **Median Stay at Home length (Days) [IQR]** | 54.0 [29-70] | 35.0 [28-68] | 42.0 [28-69] |
| **Median Days since stay at home elapsed (Days) [IQR]** | 27.0 [11-42] | 39.0 [14-45] | 38.0 [13-42] |
| **Median Stay at Home Start Date (Day since 1/22/2020) [IQR]** | 67.0 [64-72] | 69.0 [63-73] | 69.0 [63-73] |

### County-Level Mobility Trends

Mobility across all categories prior to the implementation of stay-at-home orders was similar in rural and urban counties (**Figure 1**). There was an approximately 25% increase in grocery/pharmacy mobility prior to implementation of stay-at-home orders, potentially reflecting anticipatory shopping prior to sheltering in place. This was preceded by a 15% increase and subsequent decline in retail/recreation mobility. The increase in grocery/pharmacy mobility coincided with a 25% decrease in work-place mobility and a 10% increase in residential mobility, consistent with transition to working from home.

**Figure 1.**
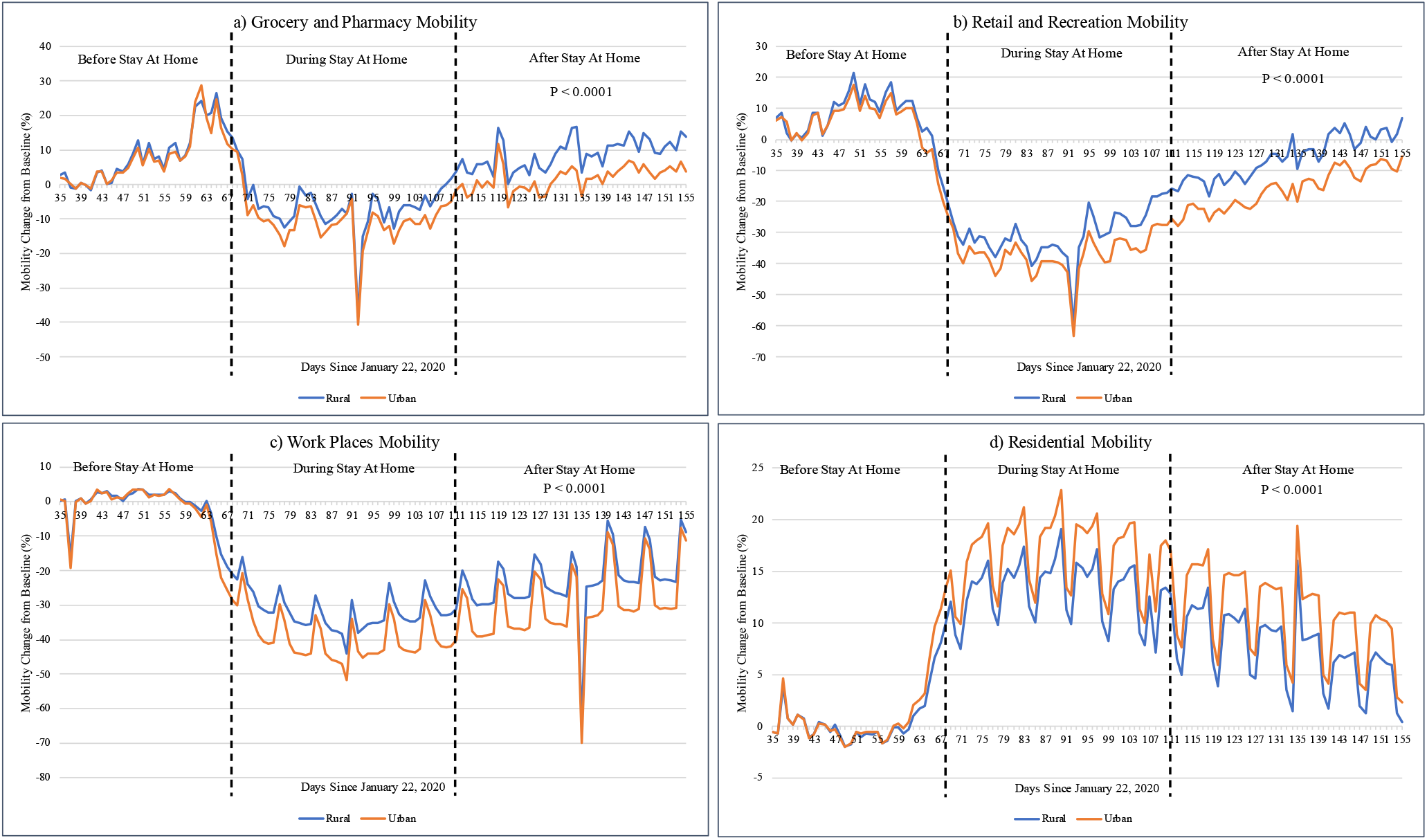
Changes in Mobility in Rural and Urban Counties. Percent changes in mobility in (A) grocery and pharmacy, (B) retail and recreation, (C) work place, and (D) residential areas were calculated for rural and urban counties compared to the referent period of January 3 and February 6, 2020. P-values report the results of repeated measure ANOVA analysis.

After implementation of stay-at-home orders, mobility in grocery/pharmacy, retail/recreation, and workplace decreased 10-40%, while residential mobility increased 10-20%. These reductions in mobility were significantly more pronounced in urban compared to rural counties, with the average absolute differences of 6.89%, 4.43%, 5.76%, and 2.67% for retail/recreation, grocery/pharmacy, workplace, and residential locations, respectively (p<0.0001). After stay-at-home orders elapsed, all mobility began to increase toward baseline levels, more rapidly in urban than rural areas. Grocery/pharmacy mobility ultimately exceeded baseline mobility in rural areas.

### County-Level Case Trends

Estimated numbers of new COVID-19 cases in rural and urban counties before, during, and after stay-at-home orders are depicted in **Figure 2**, using median dates of stay-at-home order initiation and termination for visual demonstration. In rural counties, implementation of stay-at-home orders decreased the growth of daily new COVID-19 cases from 2.1% (95% CI, 2.1%-2.2%) growth in cases per day to 0.3% (95% CI, 0.2%-0.4%) growth per day. After stay-at-home orders expired, the daily new case increase was slower than before they were put in place, with 0.2% (95% CI, -0.4% to -0.1%) decline in new cases per day. However, stay-at-home orders were more effective at preventing the spread of COVID-19 in urban counties, where the changes in the number of new daily COVID-19 cases were 4.3% (95% CI, 4.2%-4.4%) increase per day before stay-at-home, 0.7% (95% CI, -0.9% to -0.5%) decrease per day during stay-at-home, and 1% (95% CI, -1.4% to -0.6%) decrease per day after the stay-at-home order period.

**Figure 2.**
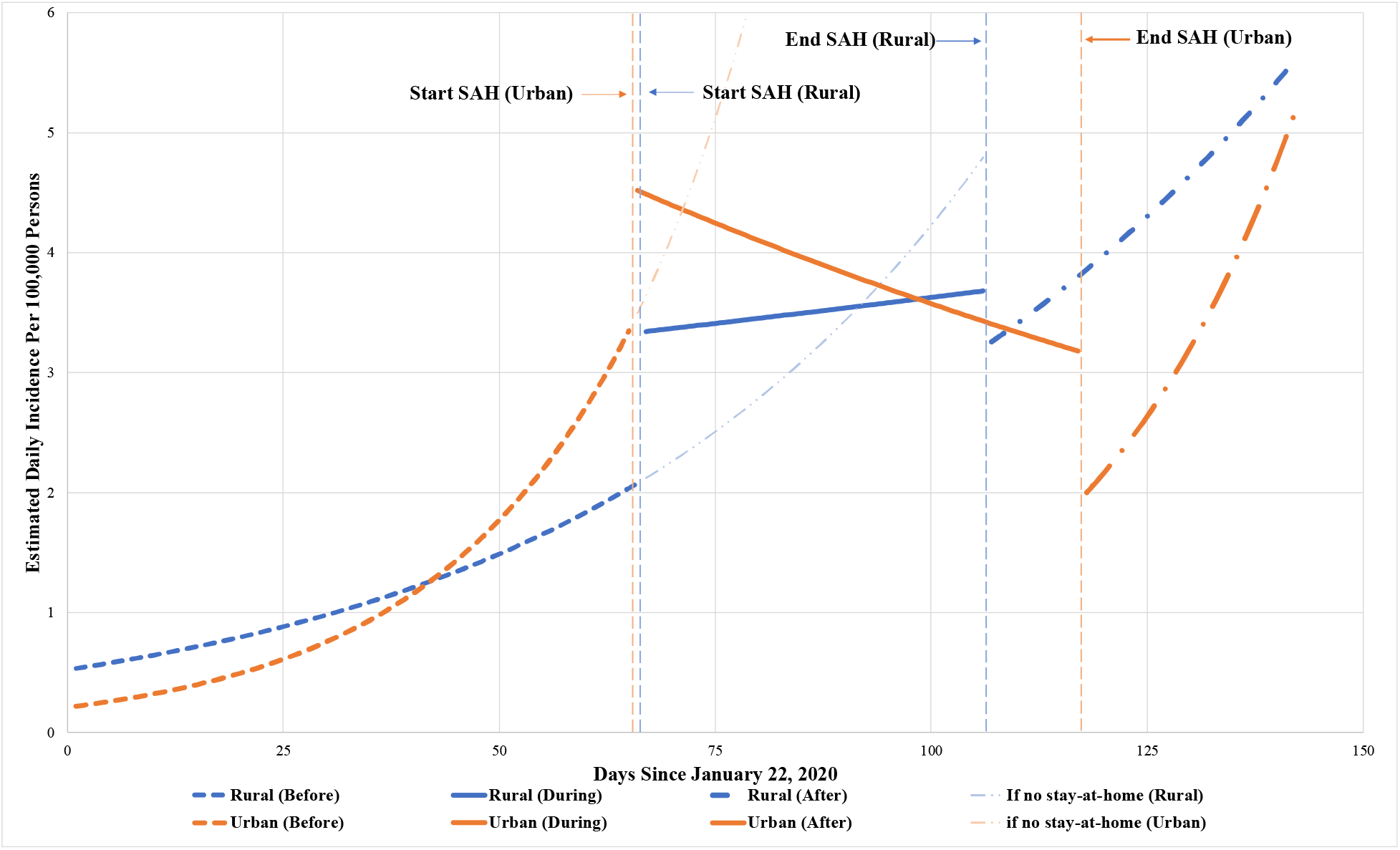
Estimated Population-Standardized Daily New Cases of COVID-19 in Rural and Urban Counties. Estimated numbers of new COVID-19 cases per day were modeled using median dates for the start and end of stay-at-home orders in rural and urban counties. We also extrapolated the predicted numbers of new daily COVID-19 infections in rural and urban counties had stay-at-home orders not been implemented to demonstrate the potential impact of these orders.

The number of new COVID-19 cases increased from baseline to during stay-at-home orders by 60.4% (IRR 1.60; 95% CI, 1.57 – 1.64) in rural counties and by 35.9% (IRR 1.36; 95% CI, 1.30 – 1.42) in urban counties; **Table 2**. For each day under stay-at-home orders, the number of daily new cases slowed by a factor of 0.982 (95% CI 0.981 – 0.982) in rural counties and 0.952 (95% CI, 0.951 – 0.953) in urban counties, i.e., slowing down more in urban counties each day under stay-at-home.

**Table 2.** Model Estimates.

| **Variable** | **Estimate (IRR)** | **2.5% CI (IRR)** | **97.5% CI (IRR)** | **P-Value** |
| --- | --- | --- | --- | --- |
| **Intercept** | -0.645 | -0.712 | -0.578 | <0.0001 |
|  | (0.525) | (0.491) | (0.561) |  |
| **Urban (vs. rural)** | -0.905 | -1.003 | -0.808 | <0.0001 |
|  | (0.404) | (0.367) | (0.446) |  |
| **Period during stay-at-home (vs. before)** | 0.473 | 0.451 | 0.494 | <0.0001 |
|  | (1.604) | (1.570) | (1.640) |  |
| **Period after stay-at-home (vs. before)** | 0.335 | 0.305 | 0.365 | <0.0001 |
|  | (1.398) | (1.357) | (1.440) |  |
| **Days elapsed since 1/22/2020** | 0.021 | 0.020 | 0.021 | <0.0001 |
|  | (1.021) | (1.021) | (1.022) |  |
| **Per day under stay-at-home orders** | -0.018 | -0.019 | -0.018 | <0.0001 |
|  | (0.982) | (0.981) | (0.982) |  |
| **Per day after stay-at-home orders** | -0.005 | -0.006 | -0.005 | <0.0001 |
|  | (0.995) | (0.994) | (0.995) |  |
| **Urban county interaction with period during stay-at-home (vs. before)** | -0.166 | -0.189 | -0.143 | <0.0001 |
|  | (0.847) | (0.828) | (0.866) |  |
| **Urban county interaction with period after stay-at-home (vs. before)** | -0.531 | -0.562 | -0.499 | <0.0001 |
|  | (0.588) | (0.570) | (0.607) |  |
| **Urban county interaction with days elapsed since 1/22/2020** | 0.022 | 0.021 | 0.022 | <0.0001 |
|  | (1.022) | (1.021) | (1.022) |  |
| **Urban county interaction with per day under stay-at-home orders** | -0.031 | -0.032 | -0.030 | <0.0001 |
|  | (0.970) | (0.969) | (0.970) |  |
| **Urban county interaction with per day after stay-at-home orders** | 0.002 | 0.001 | 0.003 | <0.0001 |
|  | (1.002) | (1.001) | (1.003) |  |
| **Urban county period during stay-at-home (vs. before)** | 0.307 | 0.262 | 0.351 | <0.0001<sup>1</sup> |
|  | (1.359) | (1.300) | (1.421) |  |
| **Urban county period after stay-at-home (vs. before)** | -0.196 | -0.257 | -0.135 | <0.0001<sup>1</sup> |
|  | (0.822) | (0.773) | (0.874) |  |
| **Urban county days elapsed since 1/22/2020** | 0.043 | 0.041 | 0.044 | <0.0001<sup>1</sup> |
|  | (1.043) | (1.042) | (1.044) |  |
| **Urban county per day under stay-at-home orders** | -0.049 | -0.051 | -0.048 | <0.0001<sup>1</sup> |
|  | (0.952) | (0.951) | (0.953) |  |
| **Urban county per day after stay-at-home orders** | -0.003 | -0.005 | -0.001 | <0.001<sup>1</sup> |
|  | (0.997) | (0.995) | (0.999) |  |

<sup>1</sup>. These variables are linear combinations meant to show the effect of multiple covariates. The p-values here were calculated using the confidence intervals and are not unique terms in the model.

After stay-at-home orders were lifted, rates of new COVID-19 infections were 39.8% greater (IRR 1.40; 95% CI, 1.36 – 1.44) than before they had been implemented in rural counties and 17.8% lower (IRR 0.82; 95% CI, 0.77 – 0.87) in urban counties, reinforcing the greater effectiveness of stay-at-home orders in urban than rural counties. Each additional day following the expiration of stay-at-home orders saw a reduction in the number of new COVID-19 cases by a factor of 0.995 in rural counties (95% CI, 0.994 – 0.995) and 0.997 in urban counties (95% CI, 0.995 – 0.999). That is to say that the rates of new cases were decreasing by 0.5% and 0.3% per day in rural and urban counties, respectively, after stay-at-home orders ended.

### Sensitivity Analysis

We examined the impact of stay-at-home orders on 5-day and 10-day lagged new cases of COVID-19 (Supplement p.). None of the study conclusions for inferences changed with either of these two different lags.

## Discussion

Physical distancing and other infection control efforts are essential to containing the spread of COVID-19. Our analysis of COVID-19 infection spread in rural and urban areas of the United States before, during, and after the implementation of stay-at-home orders revealed important successes and opportunities for improvement of this public health approach. Urban areas, which, on average, implemented stay-at-home orders earlier and maintained them longer, were able to effectively slow the growth of COVID-19 cases both while stay-at-home orders were in place and after they expired. Implementation of stay-at-home orders in rural areas was also associated with slowed the spread of COVID-19, but the observed decrease in case growth in rural areas was significantly smaller than in urban areas and the COVID-19 case load rebounded much faster after stay-at-home orders expired. These differences in the effectiveness and durability of stay-at-home orders between rural and urban areas may be driven by greater mobility of rural as compared to urban residents that we observed both during and after the stay-at-home period.

Stay-at-home orders, while they were in place, were associated with higher population-adjusted daily new cases in rural than urban areas, driven by both delayed implementation of stay-at-home orders and their shorter duration. Our analysis suggests that each additional day under stay-at-home restrictions was associated with a significant decrease in the rate of new COVID-19 infections. After stay-at-home orders expired, urban areas continued to see a much slower rate of COVID-19 spread than before restrictions were implemented, while rural areas returned near to the pre-stay-at-home baseline. This is likely driven by a range of individual, systemic, and political factors. Recent research suggests that the strongest determinants of the timing, issuance, enforcement, and adherence of stay-at-home orders are political affiliations and geography.^16 21 45-48^ Rural areas are more likely to be politically conservative and under Republican leadership^49-51^ leading to stay-at-home orders being delayed, cut short, or both, and with less enforcement of mobility restrictions.

Moreover, for stay-at-home orders to be effective and have a durable effect on infection control, individuals need to alter their daily routines and reduce high risk behaviors. Residents of rural areas consistently had higher mobility at high-risk indoor locations such as grocery stores, pharmacies, and places of retail and recreation, rendering stay-at-home orders less efficacious. This is consistent with an earlier study showing that pandemic-era declines in restaurant visits was nearly double in urban areas as compared to rural areas.^52^ Political affiliation of rural residents may have dampened their adherence to physical distancing regulations.^16 21 48^ Local news coverage of COVID-19 as a problem mainly affecting urban areas may have also diminished rural citizens’ intentions to comply.^53^ Finally, rural residents are frequently employed in fields not amenable to telework, such as agriculture, manufacturing, and service industries,^54^ and hence may be unable to fully comply with stay-at-home orders.^55^

As states are considering implementing a new round of stay-at-home orders, our findings reveal several ways to improve the implementation, enforcement, and adherence. First, they should be implemented earlier and maintained longer for optimal effectiveness. There was marked heterogeneity in the timing and duration of stay-at-home orders and our data reinforces the need for multi-jurisdictional, ideally federal, infection control mandates.^56^ Any new stay-at-home orders should be gradually implemented to avoid the pre-stay-at-home surge of mobility and subsequent spike in COVID-19 cases. To better encourage and facilitate compliance, leaders at all levels need to use scientific evidence to advocate for the importance of stay-at-home orders, set personal examples, and develop employment, housing, educational, and healthcare assistance for the most vulnerable.^55^

While our study is the first to examine the effects of stay-at-home orders on rural and urban areas, it has limitations. We focused on stay-at-home orders without considering the heterogeneity of what constituted these orders on the local level and did not separately weigh the impacts of additional measures such as school closures, nonessential business closures, prohibition of large gatherings, and mandatory masking. However, it would be difficult to separate these effects as most restrictions were implemented concurrent with, and worked in tandem to, stay-at-home orders. COVID-19 case data may be biased by differences in testing availability. However, because testing was more limited in rural than urban areas,^57^ our findings are likely an underestimate of the true difference in infection rates between urban and rural counties. We were not able to examine hospitalization and ICU utilization rates in rural and urban counties as these data are not uniformly available at the county-level. Finally, our study does not take into account potential residual or unmeasured confounders that may explain the difference in infection rates between rural and urban counties outside of stay-at-home order implementation. We accounted for this by using a random intercept in our analysis, but due to the absence of granular data at the county-level, potential confounding would be impossible to eliminate completely.

High rates of COVID-19 in rural counties, along with the suppressed effect of stay-at-home orders relative to urban counties, are very concerning. Residents of rural counties are older and have higher rates of chronic health conditions that place them at high risk for severe COVID-19 disease and death.^26 27 58^ Rural areas also lack the resources of urban areas to care for patients with COVID-19, with fewer hospitals, ICU beds, infectious disease specialists, and public health professionals.^22 28^ They may also have fewer support systems for people disabled by post-COVID-19 complications,^59^ leading to longer-term disability and personal and financial hardship. Additional research is needed to examine whether there is differential impact of stay-at-home orders on mortality and case fatality rates in rural and urban areas.

## Conclusion

Shorter duration and lower effectiveness of stay-at-home orders in rural areas have led to the greater spread of COVID-19 in rural as compared to urban areas when standardized for population. This calls for urgent reevaluation of how stay-at-home orders are designed, communicated, and implemented in rural areas and throughout the United States.

## Data Availability

All data used for this study are publicly available. The daily COVID-19 case data can be found at USAFacts, available at https://usafacts.org/visualizations/coronavirus-covid-19-spread-map/. Mobility data can be found from Google COVID-19 Community Mobility Data, available at https://www.google.com/covid19/mobility/. Stay-at-home orders were manually collected from news releases and official executive orders, this data is available from the authors upon request.

## Disclosures

### Conflict of Interest

The authors have no conflicts of interest to declare. In the past 36 months, Dr. McCoy also received support from an AARP^®^ Quality Measure Innovation Grant, the Mayo Clinic Center for Health Equity and Community Engagement Research, and the Mayo Clinic Robert D. and Patricia E. Kern Center for the Science of Health Care Delivery. In the past 36 months, Dr. Shah has received research support through Mayo Clinic from the Food and Drug Administration to establish Yale-Mayo Clinic Center for Excellence in Regulatory Science and Innovation (CERSI) program (U01FD005938); the Centers of Medicare and Medicaid Innovation under the Transforming Clinical Practice Initiative (TCPI); the Agency for Healthcare Research and Quality (1U19HS024075; R01HS025164; R01HS025402; R03HS025517); the National Heart, Lung and Blood Institute of the National Institutes of Health (NIH) (R56HL130496; R01HL131535); the National Science Foundation; and the Patient Centered Outcomes Research Institute (PCORI) to develop a Clinical Data Research Network (LHSNet).

### Funding

This effort was funded by the National Institute of Health National Institute of Diabetes and Digestive and Kidney Diseases grant K23DK114497 (McCoy) and the Mayo Clinic Research Pipeline K2R Program Award (McCoy). Study contents are the sole responsibility of the authors and do not necessarily represent the official views of NIH.

### Authors’ Contributions

D.H.J. and D.J.R. are the guarantors of the manuscript. They had full access to the data in the study and takes responsibility for the integrity of the data and accuracy of the data analysis. D.H.J. and D.J.R. designed the study, analyzed and interpreted the results and drafted the manuscript. B.D.P and N.D.S edited the manuscript and contributed to the discussion. R.G.M. supervised the study, interpreted the results, and reviewed/edited the manuscript.

## Notes

### Author Declarations

IRB approval was not required for this study.

