## Supplement for "The Association of Stay-at-Home Orders and the Spread of COVID-19 in Rural and Urban United States"

**Supplementary Materials**

**The Association of Stay-at-Home Orders and the Spread of COVID-19 in Rural and Urban United States: An interrupted time series study**

### Model Regression

#### eMethods

Analysis of the data was approached utilizing the following mixed effects count data models: Poisson (lm1glmmrelev); zero-inflated Poisson (lm2relev, lm2catziprelev, lm2catziprelev_cs_cdate, lm2catziprelev_toep_cdate, lm2catziprelev_toep_date2, and lm2catziprelev_us_date2); zero-inflated Poisson with random intercept and slope (lm2catziprelev_randslope_cdate and lm2catziprelev_randslope_date2); negative binomial (lm3glmmrelev); negative binomial with random intercept and slope (lm3glmmRandslope); zero-inflated negative binomial (lm4catziprelev). “Catzip” refers to only using the categories of dates (during and after stay-at-home orders) and their interactions with county type for the zero inflated model, instead of all of the variables used in the conditional model.

All models used the same variables for the fixed effects, as all are necessary to account for the time varying nature of stay-at-home orders. In addition, all models were offset by the population of the county divided by 100,000 to standardize the results per 100,000 people. All models were ran using the glmmTMB package in R.^1^Summary results of each model are detailed below, where URBinary represents the rurality status (a dummy variable that is 0 for rural counties and 1 for urban counties), c_daterelevduring SaH represents the stay-at-home order status (a dummy variable that is 0 for not during stay-at-home orders and 1 for during stay-at-home orders), c_daterelevafter SaH represents another indication of the stay-at-home order status (a dummy variable that is 0 for after stay-at-home orders and 1 for after stay-at-home orders), Date2 represents the number of days since January 22, 2020, dsahcarried represents the number of days under stay-at-home orders at a given time and the total number of days under stay-at-home orders while c_daterelevafter SaH is 1, asahcarried represents the number of days since the end of stay-at-home orders, URBinary:c_daterelevduring SaH represents the interaction term between the rurality status and stay-at-home order status (a dummy variable that is 0 for rural counties and for urban counties not under stay-at-home orders, and 1 for urban counties under stay-at-home orders), URBinary:c_daterelevafter SaH represents another interaction term between the rurality status and stay-at-home order status (a dummy variable that is 0 for rural counties and for urban counties not after stay-at-home orders, and 1 for urban counties after stay-at-home orders), URBinary:Date2 represents the interaction term between the number of days since January 22, 2020 and the rurality status (0 for rural counties and 1 through 142 for urban counties), URBinary:dsahcarried represents the interaction term between the number of days under stay-at-home orders and the rurality status (0 for rural counties and 0 for urban counties before stay-at-home orders), URBinary:asahcarried represents the interaction term between the number of days after stay-at-home orders and the rurality status (0 for rural counties and 0 for urban counties before the end of stay-at-home orders).

The models were compared on the basis of model diagnostics, Akaike information criterion (AIC), and parsimony (preferring non zero-inflated models where appropriate and prioritizing model diagnostics). All models were consistent in terms of estimate signs and significance.

Model diagnostics were performed examining the model’s simulated quantile scaled residuals using the DHARMA package in R.^2^ The models were assessed for over-dispersion, zero-inflation, and expected distribution of the residuals. The mixed effects negative binomial model with random intercept by county was found to be statistically significantly not zero-inflated and having normally distributed residuals, but over-dispersed and having outliers. To examine if this over-dispersion was due to the presence of outliers, the model was rerun after outliner counties (369 of 3142) were removed, but this restricted model was still over-dispersed. The models were also assessed for temporal autocorrelation using the Durbin-Watson test in the DHARMa package.^2^ The zero inflated Poisson model (lm2catziprelev) was found to only be temporally auto correlated and thus was chosen to be the best model. It was examined further using variance-covariance structures in an attempt to remove the temporal autocorrelation (lm2catziprelev_cs_cdate, lm2catziprelev_toep_cdate, lm2catziprelev_toep_date2, and lm2catziprelev_us_date2). Compound symmetry (cs_cdate) and Toeplitz (toep_cdate) structures where the only structures out of AR(1), compound symmetry, Toeplitz, and unstructured to converge using categorical date. Similarly, Toeplitz (toep_date2) and unstructured (us_date2) were the only structures able to converge using days since January 22^nd^. All attempts to remove temporal autocorrelation were inadequate and detrimental to the overall fit of the model. Temporal autocorrelation was thus deemed unavoidable. Moreover, it did not have a significant effect on the results because of the long follow-up time, the significance of the results, and the large number of counties.^3^

The final model chosen was the zero inflated Poisson model using the categories of dates and their interactions with county type for the zero inflation model (lm2catziprelev). The equations of the final model are:

$\Pr\left( Y_{ij}=y_{ij} \right)=\left\{ \begin{aligned} \pi_{ij}+\left( 1-\pi_{ij} \right)\exp\left( -\mu_{ij} \right), if y_{ij}=0 \\ \left( 1-\pi_{ij} \right)\frac{\mu_{ij}^{y_{ij}}\exp\left( -\mu_{ij} \right)}{y_{ij}!}, if y_{ij}>0 \end{aligned} \right.$	(Equation 1)

$logit\left( \pi_{ij} \right)=a_{0}+ a_{1}{Rurality}_{i}+ a_{2}{Under\_SAH}_{ij}+ a_{3}{After\_SAH}_{ij}+ a_{4}{Rurality}_{i}*{Under_{SAH}}_{ij}+a_{5}{Rurality}_{i}*{After_{SAH}}_{ij}$	(Equation 2)

$\mathrm{Log} \left( \mu_{ij} \right)=\log\left( \frac{{Population}_{i}}{100,000} \right)+ \beta_{0}+ \beta_{1}{Rurality}_{i}+ \beta_{2}{Under\_SAH}_{ij}+ \beta_{3}{After\_SAH}_{ij}+\beta_{4}{Days}_{ij} + \beta_{5}{Days\_Under\_SAH}_{ij}+ \beta_{6}{Days\_After\_SAH}_{ij}+ \beta_{7}{Rurality}_{i}*{Under\_SAH}_{ij}+\beta_{8}{Rurality}_{i}*{After\_SAH}_{ij}+\beta_{9}{Rurality}_{i}*{Days}_{ij}+ \beta_{10}{Rurality}_{i}*{Days\_Under\_SAH}_{ij}+ \beta_{11}{Rurality}_{i}*{Days\_After\_SAH}_{ij}+ b_{1i}$	(Equation 3)

where Equation 1 is the probability distribution, Equation 2 is the zero inflation model, and Equation 3 is the Poisson model. $Y_{ij}$ represents the 14-day lagged incidence of COVID-19 in the *i^th^* county on the *j^th^* day (technically the *(j+14)^th^* day) represents the probability of being 0 for the *i^th^* county on the *j^th^* day,$\mu_{ij}$ represents the 14-day lagged incidence of COVID-19 in the *i^th^* county on the *j^th^* day, $b_{i}$ represents the random effect of the *i^th^* county, *Population_i_* represents the population of the *i^th^* county, Rurality*_i_* represents the rurality status of the *i^th^* county (a dummy variable that is 0 for rural counties and 1 for urban counties), Under_SAH*_ij_* represents the stay-at-home order status of the *i^th^* county on the *j^th^* day (a dummy variable that is 0 for not during stay-at-home orders and 1 for during stay-at-home orders), After_SAH*_ij_* represents another indication of the stay-at-home order status of the *i^th^* county on the *j^th^* day (a dummy variable that is 0 for after stay-at-home orders and 1 for after stay-at-home orders), Days*_ij_* represents the number of days since January 22, 2020 for the *i^th^* county on the *j^th^* day, Days_Under_SAH*_ij_* represents the number of days under stay-at-home orders for the *i^th^* county on the *j^th^* day, Days_After_SAH*_ij_* represents the number of days since the end of stay-at-home orders for the *i^th^* county on the *j^th^* day, Rurality*Under_SAH*_ij_* represents the interaction term between the rurality status of the *i^th^* county and stay-at-home order status for the *i^th^* county on the *j^th^* day (a dummy variable that is 0 for rural counties and for urban counties not under stay-at-home orders, and 1 for urban counties under stay-at-home orders), Rurality*After_SAH*_ij_* represents another interaction term between the rurality status of the *i^th^* county and stay-at-home order status for the *i^th^* county on the *j^th^* day (a dummy variable that is 0 for rural counties and for urban counties not after stay-at-home orders, and 1 for urban counties after stay-at-home orders), Rurality*Days*_ij_* represents the interaction term between the number of days since January 22, 2020 and the rurality status for the *i^th^* county on the *j^th^* day (0 for rural counties and 1 through 142 for urban counties), Rurality*Days_Under_SAH*_ij_* represents the interaction term between the number of days under stay-at-home orders and the rurality status for the *i^th^* county on the *j^th^* day (0 for rural counties and 0 for urban counties before stay-at-home orders), Rurality*Days_After_SAH*_ij_* represents the interaction term between the number of days after stay-at-home orders and the rurality status for the *i^th^* county on the *j^th^* day (0 for rural counties and 0 for urban counties before the end of stay-at-home orders).

Therefore, $a_{0}$ represents the baseline log odds of being a “zero” day for a typical county at *j* = 0 (in that the zero inflated model assumes two zero generating processes, the first generating zeros, the top half of equation 1, and the second a Poisson process that generates counts including zeros, the bottom half of equation 1. In this case a “zero” day is one that never had the chance of being a count), $a_{1}$ represents the change in the log odds of being a zero for urban counties,$a_{2}$ represents the change in the log odds during stay-at-home orders,$a_{3}$ represents the change in the log odds after stay-at-home orders, $a_{4}$ represents the additional change in the log odds during stay-at-home orders for urban counties,$a_{5}$ represents the additional change in the log odds after stay-at-home orders for urban counties, $\beta_{0}$ represents the baseline outcome (i.e. 14-day lagged new daily cases of COVID-19) for a typical county at *j* = 0, $b_{i}$ represents the random effects (the random intercept) which is the change in baseline outcome from the typical county for the *i^th^* county (that is$\beta_{0}+b_{1}$ is the baseline outcome for the 1*^st^* county), $\beta_{1}$represents the change in the outcome for urban counties, $\beta_{2}$ represents the change in the outcome during stay-at-home orders, $\beta_{3}$ represents the change in the outcome after stay-at-home orders, $\beta_{4}$ represents the change in the outcome for each day since *j* =0 (January 22, 2020), $\beta_{5}$ represents the change in the outcome for each day a county was under stay-at-home orders, $\beta_{6}$ represents the change in the outcome for each day a county was out of stay-at-home orders, $\beta_{7}$ represents the additional change in the outcome for urban counties during stay-at-home orders (that is for urban counties the “actual $\beta_{2}$” is $\beta_{2}$ + $\beta_{7}$), $\beta_{8}$ represents the additional change in the outcome for urban counties after stay-at-home orders, $\beta_{9}$ represents the additional change in the outcome for each day since *j* =0 (January 22, 2020), $\beta_{10}$ represents the additional change in the outcome for urban counties for each day it was under stay-at-home orders, $\beta_{11}$ represents the additional change in the outcome for urban counties for each day it was out of stay-at-home orders.

#### Figure 2 Generation

Figure 2 was generated by inputting the estimates of fixed effects and the urban and rural averages of stay-at-home orders start and end dates. The outcome was divided by the offset to standardize the results per 100,000 population. The respective offsets for urban and rural counties were calculated using urban and rural counties respective population averages. Similarly, the extrapolations were generated by using the conditional model only with intercept and variables: Rurality, Days, and Rurality*Days. The extrapolations represent continuation of the before stay-at-home order trends.

#### summary(lm1glmmrelev)

```
##  Family: poisson  ( log )
## Formula:
## newcase_nst_14 ~ offset(popoff) + URBinary * c_daterelev + URBinary *
##     Date2 + URBinary * dsahcarried + URBinary * asahcarried +      (1 | c_FIPS)
## Data: df_14
##
##      AIC      BIC   logLik deviance df.resid
##  2575878  2576022 -1287926  2575852   446151
##
## Random effects:
##
## Conditional model:
##  Groups Name        Variance Std.Dev.
##  c_FIPS (Intercept) 1.904    1.38
## Number of obs: 446164, groups:  c_FIPS, 3142
##
## Conditional model:
##                                  Estimate Std. Error z value Pr(>|z|)
## (Intercept)                    -2.6640807  0.0345759  -77.05   <2e-16 ***
## URBinary                       -1.0865939  0.0539059  -20.16   <2e-16 ***
## c_daterelevafter SaH            0.7165783  0.0142383   50.33   <2e-16 ***
## c_daterelevduring SaH           0.8961563  0.0102393   87.52   <2e-16 ***
## Date2                           0.0336172  0.0001930  174.18   <2e-16 ***
## dsahcarried                    -0.0276698  0.0002731 -101.30   <2e-16 ***
## asahcarried                    -0.0170237  0.0003703  -45.97   <2e-16 ***
## URBinary:c_daterelevafter SaH  -1.0547728  0.0150346  -70.16   <2e-16 ***
## URBinary:c_daterelevduring SaH -0.7052632  0.0106686  -66.11   <2e-16 ***
## URBinary:Date2                  0.0386741  0.0002361  163.83   <2e-16 ***
## URBinary:dsahcarried           -0.0511247  0.0003085 -165.74   <2e-16 ***
## URBinary:asahcarried           -0.0151175  0.0004177  -36.19   <2e-16 ***
## ---
## Signif. codes:  0 '***' 0.001 '**' 0.01 '*' 0.05 '.' 0.1 ' ' 1
```

#### summary(lm2relev)

```
##  Family: poisson  ( log )
## Formula:
## newcase_nst_14 ~ offset(popoff) + URBinary * c_daterelev + URBinary *
##     Date2 + URBinary * dsahcarried + URBinary * asahcarried + (1 | c_FIPS)
## Zero inflation:
## ~URBinary * c_daterelev + URBinary * Date2 + URBinary * dsahcarried +
##     URBinary * asahcarried
## Data: df_14
##
##      AIC      BIC   logLik deviance df.resid
##  2185560  2185835 -1092755  2185510   446139
##
## Random effects:
##
## Conditional model:
##  Groups Name        Variance Std.Dev.
##  c_FIPS (Intercept) 1.493    1.222
## Number of obs: 446164, groups:  c_FIPS, 3142
##
## Conditional model:
##                                  Estimate Std. Error z value Pr(>|z|)
## (Intercept)                     0.3294486  0.0345318    9.54  < 2e-16 ***
## URBinary                       -1.7134201  0.0507236  -33.78  < 2e-16 ***
## c_daterelevafter SaH            0.3334240  0.0149715   22.27  < 2e-16 ***
## c_daterelevduring SaH           0.4153541  0.0104818   39.63  < 2e-16 ***
## Date2                           0.0072637  0.0002595   27.99  < 2e-16 ***
## dsahcarried                    -0.0068868  0.0003361  -20.49  < 2e-16 ***
## asahcarried                     0.0074896  0.0004262   17.57  < 2e-16 ***
## URBinary:c_daterelevafter SaH  -0.4997899  0.0157299  -31.77  < 2e-16 ***
## URBinary:c_daterelevduring SaH -0.0822104  0.0108929   -7.55 4.45e-14 ***
## URBinary:Date2                  0.0319725  0.0003020  105.88  < 2e-16 ***
## URBinary:dsahcarried           -0.0393398  0.0003725 -105.61  < 2e-16 ***
## URBinary:asahcarried           -0.0074082  0.0004742  -15.62  < 2e-16 ***
## ---
## Signif. codes:  0 '***' 0.001 '**' 0.01 '*' 0.05 '.' 0.1 ' ' 1
##
## Zero-inflation model:
##                                  Estimate Std. Error z value Pr(>|z|)
## (Intercept)                     3.9325320  0.0368018  106.86  < 2e-16 ***
## URBinary                        0.2730539  0.0524306    5.21 1.91e-07 ***
## c_daterelevafter SaH           -0.2081402  0.0469575   -4.43 9.31e-06 ***
## c_daterelevduring SaH          -0.4189844  0.0285833  -14.66  < 2e-16 ***
## Date2                          -0.0524315  0.0006947  -75.47  < 2e-16 ***
## dsahcarried                     0.0372385  0.0007767   47.95  < 2e-16 ***
## asahcarried                     0.0428863  0.0014264   30.07  < 2e-16 ***
## URBinary:c_daterelevafter SaH   1.5684013  0.0823941   19.04  < 2e-16 ***
## URBinary:c_daterelevduring SaH  0.8701813  0.0440532   19.75  < 2e-16 ***
## URBinary:Date2                 -0.0353790  0.0010536  -33.58  < 2e-16 ***
## URBinary:dsahcarried            0.0132444  0.0012072   10.97  < 2e-16 ***
## URBinary:asahcarried            0.0426762  0.0024530   17.40  < 2e-16 ***
## ---
## Signif. codes:  0 '***' 0.001 '**' 0.01 '*' 0.05 '.' 0.1 ' ' 1
```

#### summary(lm2catziprelev)

```
##  Family: poisson  ( log )
## Formula:
## newcase_nst_14 ~ offset(popoff) + URBinary * c_daterelev + URBinary *
##     Date2 + URBinary * dsahcarried + URBinary * asahcarried + (1 | c_FIPS)
## Zero inflation:                  ~URBinary * c_daterelev
## Data: df_14
##
##      AIC      BIC   logLik deviance df.resid
##  2220521  2220730 -1110242  2220483   446145
##
## Random effects:
##
## Conditional model:
##  Groups Name        Variance Std.Dev.
##  c_FIPS (Intercept) 1.389    1.179
## Number of obs: 446164, groups:  c_FIPS, 3142
##
## Conditional model:
##                                  Estimate Std. Error z value Pr(>|z|)
## (Intercept)                    -0.6452101  0.0342097  -18.86  < 2e-16 ***
## URBinary                       -0.9052841  0.0496647  -18.23  < 2e-16 ***
## c_daterelevafter SaH            0.3348040  0.0152451   21.96  < 2e-16 ***
## c_daterelevduring SaH           0.4726759  0.0111321   42.46  < 2e-16 ***
## Date2                           0.0208674  0.0002576   81.02  < 2e-16 ***
## dsahcarried                    -0.0184094  0.0003317  -55.50  < 2e-16 ***
## asahcarried                    -0.0054122  0.0004206  -12.87  < 2e-16 ***
## URBinary:c_daterelevafter SaH  -0.5307952  0.0159890  -33.20  < 2e-16 ***
## URBinary:c_daterelevduring SaH -0.1659420  0.0115200  -14.40  < 2e-16 ***
## URBinary:Date2                  0.0215742  0.0003019   71.46  < 2e-16 ***
## URBinary:dsahcarried           -0.0309239  0.0003699  -83.60  < 2e-16 ***
## URBinary:asahcarried            0.0023423  0.0004701    4.98 6.28e-07 ***
## ---
## Signif. codes:  0 '***' 0.001 '**' 0.01 '*' 0.05 '.' 0.1 ' ' 1
##
## Zero-inflation model:
##                                Estimate Std. Error z value Pr(>|z|)
## (Intercept)                     0.83343    0.01385   60.19   <2e-16 ***
## URBinary                       -0.59437    0.01740  -34.15   <2e-16 ***
## c_daterelevafter SaH           -1.37483    0.02111  -65.13   <2e-16 ***
## c_daterelevduring SaH          -1.08682    0.01874  -57.99   <2e-16 ***
## URBinary:c_daterelevafter SaH  -0.55237    0.03219  -17.16   <2e-16 ***
## URBinary:c_daterelevduring SaH -0.80902    0.02630  -30.77   <2e-16 ***
## ---
## Signif. codes:  0 '***' 0.001 '**' 0.01 '*' 0.05 '.' 0.1 ' ' 1
```

#### summary(lm2catziprelev_cs_cdate)

```
##  Family: poisson  ( log )
## Formula:
## newcase_nst_14 ~ offset(popoff) + URBinary * c_daterelev + URBinary *
##     Date2 + URBinary * dsahcarried + URBinary * asahcarried +
##     (1 | c_FIPS) + cs(c_daterelev + 0 | c_FIPS)
## Zero inflation:                  ~URBinary * c_daterelev
## Data: df_14
##
##       AIC       BIC    logLik  deviance  df.resid
## 1886145.8 1886399.0 -943049.9 1886099.8    446141
##
## Random effects:
##
## Conditional model:
##  Groups   Name                  Variance  Std.Dev. Corr
##  c_FIPS   (Intercept)           4.359e-06 0.002088
##  c_FIPS.1 c_daterelevbefore SaH 1.395e+00 1.181049 0.56 (cs)
## Number of obs: 446164, groups:  c_FIPS, 3142
##
## Conditional model:
##                                  Estimate Std. Error z value Pr(>|z|)
## (Intercept)                    -1.1492470  0.0370040  -31.06  < 2e-16 ***
## URBinary                       -0.8032034  0.0521819  -15.39  < 2e-16 ***
## c_daterelevafter SaH            0.5686569  0.0400956   14.18  < 2e-16 ***
## c_daterelevduring SaH           0.5825936  0.0381258   15.28  < 2e-16 ***
## Date2                           0.0226415  0.0002548   88.85  < 2e-16 ***
## dsahcarried                    -0.0211272  0.0003418  -61.81  < 2e-16 ***
## asahcarried                    -0.0076120  0.0004400  -17.30  < 2e-16 ***
## URBinary:c_daterelevafter SaH  -0.4869172  0.0550463   -8.85  < 2e-16 ***
## URBinary:c_daterelevduring SaH -0.2393014  0.0533266   -4.49 7.21e-06 ***
## URBinary:Date2                  0.0224831  0.0003010   74.68  < 2e-16 ***
## URBinary:dsahcarried           -0.0307955  0.0003805  -80.94  < 2e-16 ***
## URBinary:asahcarried           -0.0024864  0.0004938   -5.03 4.78e-07 ***
## ---
## Signif. codes:  0 '***' 0.001 '**' 0.01 '*' 0.05 '.' 0.1 ' ' 1
##
## Zero-inflation model:
##                                Estimate Std. Error z value Pr(>|z|)
## (Intercept)                     0.49083    0.01631   30.09   <2e-16 ***
## URBinary                       -0.43144    0.02007  -21.49   <2e-16 ***
## c_daterelevafter SaH           -1.23339    0.02466  -50.01   <2e-16 ***
## c_daterelevduring SaH          -0.99054    0.02247  -44.09   <2e-16 ***
## URBinary:c_daterelevafter SaH  -0.62784    0.03622  -17.33   <2e-16 ***
## URBinary:c_daterelevduring SaH -0.94321    0.03091  -30.52   <2e-16 ***
## ---
## Signif. codes:  0 '***' 0.001 '**' 0.01 '*' 0.05 '.' 0.1 ' ' 1
```

#### summary(lm2catziprelev_toep_cdate)

```
##  Family: poisson  ( log )
## Formula:
## newcase_nst_14 ~ offset(popoff) + URBinary * c_daterelev + URBinary *
##     Date2 + URBinary * dsahcarried + URBinary * asahcarried +
##     (1 | c_FIPS) + toep(c_daterelev + 0 | c_FIPS)
## Zero inflation:                  ~URBinary * c_daterelev
## Data: df_14
##
##       AIC       BIC    logLik  deviance  df.resid
## 1886153.2 1886417.4 -943052.6 1886105.2    446140
##
## Random effects:
##
## Conditional model:
##  Groups   Name                  Variance  Std.Dev. Corr
##  c_FIPS   (Intercept)           3.133e-06 0.00177
##  c_FIPS.1 c_daterelevbefore SaH 1.388e+00 1.17807
##           c_daterelevafter SaH  1.590e+00 1.26104  0.55
##           c_daterelevduring SaH 1.828e+00 1.35185  0.59 0.55
## Number of obs: 446164, groups:  c_FIPS, 3142
##
## Conditional model:
##                                  Estimate Std. Error z value Pr(>|z|)
## (Intercept)                    -1.1466523  0.0369283  -31.05  < 2e-16 ***
## URBinary                       -0.8003107  0.0520796  -15.37  < 2e-16 ***
## c_daterelevafter SaH            0.5676059  0.0401813   14.13  < 2e-16 ***
## c_daterelevduring SaH           0.5815646  0.0374309   15.54  < 2e-16 ***
## Date2                           0.0226389  0.0002548   88.85  < 2e-16 ***
## dsahcarried                    -0.0211143  0.0003418  -61.77  < 2e-16 ***
## asahcarried                    -0.0076127  0.0004397  -17.31  < 2e-16 ***
## URBinary:c_daterelevafter SaH  -0.4894416  0.0553070   -8.85  < 2e-16 ***
## URBinary:c_daterelevduring SaH -0.2472263  0.0521449   -4.74 2.13e-06 ***
## URBinary:Date2                  0.0224768  0.0003010   74.67  < 2e-16 ***
## URBinary:dsahcarried           -0.0307992  0.0003804  -80.96  < 2e-16 ***
## URBinary:asahcarried           -0.0024785  0.0004936   -5.02 5.13e-07 ***
## ---
## Signif. codes:  0 '***' 0.001 '**' 0.01 '*' 0.05 '.' 0.1 ' ' 1
##
## Zero-inflation model:
##                                Estimate Std. Error z value Pr(>|z|)
## (Intercept)                     0.49144    0.01631   30.13   <2e-16 ***
## URBinary                       -0.43017    0.02007  -21.44   <2e-16 ***
## c_daterelevafter SaH           -1.23361    0.02467  -50.00   <2e-16 ***
## c_daterelevduring SaH          -0.99132    0.02247  -44.12   <2e-16 ***
## URBinary:c_daterelevafter SaH  -0.62941    0.03622  -17.38   <2e-16 ***
## URBinary:c_daterelevduring SaH -0.94446    0.03090  -30.56   <2e-16 ***
## ---
## Signif. codes:  0 '***' 0.001 '**' 0.01 '*' 0.05 '.' 0.1 ' ' 1
```

#### summary(lm2catziprelev_toep_date2)

```
##  Family: poisson  ( log )
## Formula:
## newcase_nst_14 ~ offset(popoff) + URBinary * c_daterelev + URBinary *
##     Date2 + URBinary * dsahcarried + URBinary * asahcarried +
##     (1 | c_FIPS) + toep(Date2 + 0 | c_FIPS)
## Zero inflation:                  ~URBinary * c_daterelev
## Data: df_14
##
##       AIC       BIC    logLik  deviance  df.resid
## 1675467.5 1675687.7 -837713.8 1675427.5    446144
##
## Random effects:
##
## Conditional model:
##  Groups   Name        Variance Std.Dev. Corr
##  c_FIPS   (Intercept) 7.092575 2.66319
##  c_FIPS.1 Date2       0.001084 0.03292
## Number of obs: 446164, groups:  c_FIPS, 3142
##
## Conditional model:
##                                  Estimate Std. Error z value Pr(>|z|)
## (Intercept)                    -4.6939201  0.0779133  -60.25  < 2e-16 ***
## URBinary                       -1.0536716  0.1119397   -9.41  < 2e-16 ***
## c_daterelevafter SaH           -0.2636837  0.0177526  -14.85  < 2e-16 ***
## c_daterelevduring SaH          -0.0369320  0.0130893   -2.82  0.00478 **
## Date2                           0.0800563  0.0010290   77.80  < 2e-16 ***
## dsahcarried                    -0.0876383  0.0009001  -97.36  < 2e-16 ***
## asahcarried                    -0.0918231  0.0009535  -96.30  < 2e-16 ***
## URBinary:c_daterelevafter SaH  -0.2738945  0.0185531  -14.76  < 2e-16 ***
## URBinary:c_daterelevduring SaH -0.0194567  0.0134725   -1.44  0.14869
## URBinary:Date2                  0.0314464  0.0014375   21.88  < 2e-16 ***
## URBinary:dsahcarried           -0.0376001  0.0009373  -40.12  < 2e-16 ***
## URBinary:asahcarried            0.0072034  0.0010036    7.18 7.08e-13 ***
## ---
## Signif. codes:  0 '***' 0.001 '**' 0.01 '*' 0.05 '.' 0.1 ' ' 1
##
## Zero-inflation model:
##                                Estimate Std. Error z value Pr(>|z|)
## (Intercept)                    -0.03036    0.01702   -1.78   0.0745 .
## URBinary                       -0.77990    0.02329  -33.49  < 2e-16 ***
## c_daterelevafter SaH           -0.72481    0.02506  -28.92  < 2e-16 ***
## c_daterelevduring SaH          -0.40245    0.02194  -18.34  < 2e-16 ***
## URBinary:c_daterelevafter SaH  -0.30267    0.03817   -7.93  2.2e-15 ***
## URBinary:c_daterelevduring SaH -0.56191    0.03156  -17.80  < 2e-16 ***
## ---
## Signif. codes:  0 '***' 0.001 '**' 0.01 '*' 0.05 '.' 0.1 ' ' 1
```

#### summary(lm2catziprelev_us_date2)

```
##  Family: poisson  ( log )
## Formula:
## newcase_nst_14 ~ offset(popoff) + URBinary * c_daterelev + URBinary *
##     Date2 + URBinary * dsahcarried + URBinary * asahcarried +
##     (1 | c_FIPS) + us(Date2 + 0 | c_FIPS)
## Zero inflation:                  ~URBinary * c_daterelev
## Data: df_14
##
##       AIC       BIC    logLik  deviance  df.resid
## 1675467.5 1675687.7 -837713.8 1675427.5    446144
##
## Random effects:
##
## Conditional model:
##  Groups   Name        Variance Std.Dev.
##  c_FIPS   (Intercept) 7.092575 2.66319
##  c_FIPS.1 Date2       0.001084 0.03292
## Number of obs: 446164, groups:  c_FIPS, 3142
##
## Conditional model:
##                                  Estimate Std. Error z value Pr(>|z|)
## (Intercept)                    -4.6939201  0.0779133  -60.25  < 2e-16 ***
## URBinary                       -1.0536716  0.1119397   -9.41  < 2e-16 ***
## c_daterelevafter SaH           -0.2636837  0.0177526  -14.85  < 2e-16 ***
## c_daterelevduring SaH          -0.0369320  0.0130893   -2.82  0.00478 **
## Date2                           0.0800563  0.0010290   77.80  < 2e-16 ***
## dsahcarried                    -0.0876383  0.0009001  -97.36  < 2e-16 ***
## asahcarried                    -0.0918231  0.0009535  -96.30  < 2e-16 ***
## URBinary:c_daterelevafter SaH  -0.2738945  0.0185531  -14.76  < 2e-16 ***
## URBinary:c_daterelevduring SaH -0.0194567  0.0134725   -1.44  0.14869
## URBinary:Date2                  0.0314464  0.0014375   21.88  < 2e-16 ***
## URBinary:dsahcarried           -0.0376001  0.0009373  -40.12  < 2e-16 ***
## URBinary:asahcarried            0.0072034  0.0010036    7.18 7.08e-13 ***
## ---
## Signif. codes:  0 '***' 0.001 '**' 0.01 '*' 0.05 '.' 0.1 ' ' 1
##
## Zero-inflation model:
##                                Estimate Std. Error z value Pr(>|z|)
## (Intercept)                    -0.03036    0.01702   -1.78   0.0745 .
## URBinary                       -0.77990    0.02329  -33.49  < 2e-16 ***
## c_daterelevafter SaH           -0.72481    0.02506  -28.92  < 2e-16 ***
## c_daterelevduring SaH          -0.40245    0.02194  -18.34  < 2e-16 ***
## URBinary:c_daterelevafter SaH  -0.30267    0.03817   -7.93  2.2e-15 ***
## URBinary:c_daterelevduring SaH -0.56191    0.03156  -17.80  < 2e-16 ***
## ---
## Signif. codes:  0 '***' 0.001 '**' 0.01 '*' 0.05 '.' 0.1 ' ' 1
```

#### summary(lm2catziprelev_randslope_cdate)

```
##  Family: poisson  ( log )
## Formula:
## newcase_nst_14 ~ offset(popoff) + URBinary * c_daterelev + URBinary *
##     Date2 + URBinary * dsahcarried + URBinary * asahcarried +
##     (1 + c_daterelev | c_FIPS)
## Zero inflation:                  ~URBinary * c_daterelev
## Data: df_14
##
##       AIC       BIC    logLik  deviance  df.resid
## 1885839.9 1886104.1 -942895.9 1885791.9    446140
##
## Random effects:
##
## Conditional model:
##  Groups Name                  Variance Std.Dev. Corr
##  c_FIPS (Intercept)           1.327    1.152
##         c_daterelevafter SaH  1.862    1.365    -0.51
##         c_daterelevduring SaH 1.425    1.194    -0.30  0.67
## Number of obs: 446164, groups:  c_FIPS, 3142
##
## Conditional model:
##                                  Estimate Std. Error z value Pr(>|z|)
## (Intercept)                    -1.1358486  0.0365213  -31.10  < 2e-16 ***
## URBinary                       -0.8211116  0.0513008  -16.01  < 2e-16 ***
## c_daterelevafter SaH            0.5781164  0.0437979   13.20  < 2e-16 ***
## c_daterelevduring SaH           0.5572114  0.0381538   14.60  < 2e-16 ***
## Date2                           0.0226873  0.0002546   89.11  < 2e-16 ***
## dsahcarried                    -0.0212549  0.0003420  -62.14  < 2e-16 ***
## asahcarried                    -0.0075466  0.0004414  -17.10  < 2e-16 ***
## URBinary:c_daterelevafter SaH  -0.4409249  0.0621032   -7.10 1.25e-12 ***
## URBinary:c_daterelevduring SaH -0.2174602  0.0534720   -4.07 4.77e-05 ***
## URBinary:Date2                  0.0224540  0.0003008   74.64  < 2e-16 ***
## URBinary:dsahcarried           -0.0306887  0.0003806  -80.62  < 2e-16 ***
## URBinary:asahcarried           -0.0025590  0.0004951   -5.17 2.35e-07 ***
## ---
## Signif. codes:  0 '***' 0.001 '**' 0.01 '*' 0.05 '.' 0.1 ' ' 1
##
## Zero-inflation model:
##                                Estimate Std. Error z value Pr(>|z|)
## (Intercept)                     0.48961    0.01632   30.01   <2e-16 ***
## URBinary                       -0.43090    0.02008  -21.46   <2e-16 ***
## c_daterelevafter SaH           -1.22573    0.02522  -48.60   <2e-16 ***
## c_daterelevduring SaH          -0.99062    0.02246  -44.10   <2e-16 ***
## URBinary:c_daterelevafter SaH  -0.63294    0.03662  -17.29   <2e-16 ***
## URBinary:c_daterelevduring SaH -0.94362    0.03090  -30.54   <2e-16 ***
## ---
## Signif. codes:  0 '***' 0.001 '**' 0.01 '*' 0.05 '.' 0.1 ' ' 1
```

#### summary(lm2catziprelev_randslope_date2)

```
##  Family: poisson  ( log )
## Formula:
## newcase_nst_14 ~ offset(popoff) + URBinary * c_daterelev + URBinary *
##     Date2 + URBinary * dsahcarried + URBinary * asahcarried +
##     (1 + Date2 | c_FIPS)
## Zero inflation:                  ~URBinary * c_daterelev
## Data: df_14
##
##       AIC       BIC    logLik  deviance  df.resid
## 1672172.5 1672403.7 -836065.3 1672130.5    446143
##
## Random effects:
##
## Conditional model:
##  Groups Name        Variance Std.Dev. Corr
##  c_FIPS (Intercept) 7.896127 2.81000
##         Date2       0.001253 0.03539  -0.84
## Number of obs: 446164, groups:  c_FIPS, 3142
##
## Conditional model:
##                                  Estimate Std. Error z value Pr(>|z|)
## (Intercept)                    -4.6156791  0.0800240  -57.68  < 2e-16 ***
## URBinary                       -1.1205871  0.1164587   -9.62  < 2e-16 ***
## c_daterelevafter SaH           -0.2686742  0.0178768  -15.03  < 2e-16 ***
## c_daterelevduring SaH          -0.0401562  0.0132186   -3.04  0.00238 **
## Date2                           0.0816669  0.0010739   76.05  < 2e-16 ***
## dsahcarried                    -0.0898446  0.0009347  -96.12  < 2e-16 ***
## asahcarried                    -0.0951357  0.0009879  -96.31  < 2e-16 ***
## URBinary:c_daterelevafter SaH  -0.2696256  0.0186693  -14.44  < 2e-16 ***
## URBinary:c_daterelevduring SaH -0.0166123  0.0135937   -1.22  0.22168
## URBinary:Date2                  0.0299902  0.0015192   19.74  < 2e-16 ***
## URBinary:dsahcarried           -0.0355063  0.0009702  -36.60  < 2e-16 ***
## URBinary:asahcarried            0.0103444  0.0010368    9.98  < 2e-16 ***
## ---
## Signif. codes:  0 '***' 0.001 '**' 0.01 '*' 0.05 '.' 0.1 ' ' 1
##
## Zero-inflation model:
##                                Estimate Std. Error z value Pr(>|z|)
## (Intercept)                    -0.02154    0.01712   -1.26    0.208
## URBinary                       -0.78216    0.02335  -33.50   <2e-16 ***
## c_daterelevafter SaH           -0.73417    0.02516  -29.18   <2e-16 ***
## c_daterelevduring SaH          -0.42532    0.02208  -19.27   <2e-16 ***
## URBinary:c_daterelevafter SaH  -0.29925    0.03823   -7.83    5e-15 ***
## URBinary:c_daterelevduring SaH -0.54751    0.03164  -17.30   <2e-16 ***
## ---
## Signif. codes:  0 '***' 0.001 '**' 0.01 '*' 0.05 '.' 0.1 ' ' 1
```

#### summary(lm3glmmrelev)

```
##  Family: nbinom2  ( log )
## Formula:
## newcase_nst_14 ~ offset(popoff) + URBinary * c_daterelev + URBinary *
##     Date2 + URBinary * dsahcarried + URBinary * asahcarried +      (1 | c_FIPS)
## Data: df_14
##
##       AIC       BIC    logLik  deviance  df.resid
##  986461.7  986615.8 -493216.9  986433.7    446150
##
## Random effects:
##
## Conditional model:
##  Groups Name        Variance Std.Dev.
##  c_FIPS (Intercept) 1.963    1.401
## Number of obs: 446164, groups:  c_FIPS, 3142
##
## Overdispersion parameter for nbinom2 family (): 0.599
##
## Conditional model:
##                                  Estimate Std. Error z value Pr(>|z|)
## (Intercept)                    -3.6817667  0.0419566  -87.75   <2e-16 ***
## URBinary                       -1.5409535  0.0656975  -23.46   <2e-16 ***
## c_daterelevafter SaH            0.2696026  0.0283545    9.51   <2e-16 ***
## c_daterelevduring SaH           0.5163268  0.0182598   28.28   <2e-16 ***
## Date2                           0.0499635  0.0004014  124.48   <2e-16 ***
## dsahcarried                    -0.0398040  0.0005698  -69.85   <2e-16 ***
## asahcarried                    -0.0263350  0.0008609  -30.59   <2e-16 ***
## URBinary:c_daterelevafter SaH  -0.9275987  0.0386698  -23.99   <2e-16 ***
## URBinary:c_daterelevduring SaH -0.6635130  0.0244069  -27.19   <2e-16 ***
## URBinary:Date2                  0.0459769  0.0006779   67.83   <2e-16 ***
## URBinary:dsahcarried           -0.0488390  0.0008528  -57.27   <2e-16 ***
## URBinary:asahcarried           -0.0384419  0.0012898  -29.80   <2e-16 ***
## ---
## Signif. codes:  0 '***' 0.001 '**' 0.01 '*' 0.05 '.' 0.1 ' ' 1
```

#### summary(lm3glmmRandslope)

```
##  Family: nbinom2  ( log )
## Formula:
## newcase_nst_14 ~ offset(popoff) + URBinary * c_daterelev + URBinary *
##     Date2 + URBinary * dsahcarried + URBinary * asahcarried +
##     (1 + c_daterelev | c_FIPS)
## Data: df_14
##
##       AIC       BIC    logLik  deviance  df.resid
##  965591.5  965800.6 -482776.7  965553.5    446145
##
## Random effects:
##
## Conditional model:
##  Groups Name                  Variance Std.Dev. Corr
##  c_FIPS (Intercept)           1.714    1.309
##         c_daterelevafter SaH  2.044    1.430    -0.51
##         c_daterelevduring SaH 1.559    1.249    -0.26  0.73
## Number of obs: 446164, groups:  c_FIPS, 3142
##
## Overdispersion parameter for nbinom2 family (): 0.742
##
## Conditional model:
##                                  Estimate Std. Error z value Pr(>|z|)
## (Intercept)                    -3.6799074  0.0422226  -87.16   <2e-16 ***
## URBinary                       -1.6822882  0.0641426  -26.23   <2e-16 ***
## c_daterelevafter SaH            0.5330086  0.0504697   10.56   <2e-16 ***
## c_daterelevduring SaH           0.5997640  0.0417863   14.35   <2e-16 ***
## Date2                           0.0481627  0.0003801  126.70   <2e-16 ***
## dsahcarried                    -0.0408738  0.0005746  -71.13   <2e-16 ***
## asahcarried                    -0.0245185  0.0008627  -28.42   <2e-16 ***
## URBinary:c_daterelevafter SaH  -0.7504941  0.0717474  -10.46   <2e-16 ***
## URBinary:c_daterelevduring SaH -0.5525770  0.0580217   -9.52   <2e-16 ***
## URBinary:Date2                  0.0471780  0.0006432   73.35   <2e-16 ***
## URBinary:dsahcarried           -0.0526096  0.0008429  -62.42   <2e-16 ***
## URBinary:asahcarried           -0.0401380  0.0012818  -31.31   <2e-16 ***
## ---
## Signif. codes:  0 '***' 0.001 '**' 0.01 '*' 0.05 '.' 0.1 ' ' 1
```

#### summary(lm4catziprelev)

\## Family: nbinom2 ( log )
\## Formula:
\## newcase_nst_14 ~ offset(popoff) + URBinary * c_daterelev + URBinary *
\##	Date2 + URBinary * dsahcarried + URBinary * asahcarried +	(1 | c_FIPS)
\## Zero inflation: ~URBinary * c_daterelev
\## Data: df_14
\##
\##	AIC	BIC	logLik deviance df.resid
\## 983260.7 983480.9 -491610.4 983220.7	446144
\##
\## Random effects:
\##
\## Conditional model:
\## Groups Name	Variance Std.Dev.
\## c_FIPS (Intercept) 1.915	1.384
\## Number of obs: 446164, groups: c_FIPS, 3142
\##
\## Overdispersion parameter for nbinom2 family (): 0.717
\##
\## Conditional model:
\##	Estimate Std. Error z value Pr(>|z|)
\## (Intercept)	-3.3261455 0.0449990 -73.92	<2e-16 ***
\## URBinary	-1.5427435 0.0690553 -22.34	<2e-16 ***
\## c_daterelevafter SaH	0.0112667 0.0301700	0.37	0.709
\## c_daterelevduring SaH	0.2390741 0.0212071	11.27	<2e-16 ***
\## Date2	0.0502714 0.0004456 112.83	<2e-16 ***
\## dsahcarried	-0.0406409 0.0006016 -67.55	<2e-16 ***
\## asahcarried	-0.0261313 0.0008803 -29.69	<2e-16 ***
\## URBinary:c_daterelevafter SaH -0.7592662 0.0391172 -19.41	<2e-16 ***
\## URBinary:c_daterelevduring SaH -0.5058538 0.0264575 -19.12	<2e-16 ***
\## URBinary:Date2	0.0425168 0.0007429	57.23	<2e-16 ***
\## URBinary:dsahcarried	-0.0458900 0.0008931 -51.38	<2e-16 ***
\## URBinary:asahcarried	-0.0350415 0.0012934 -27.09	<2e-16 ***
\## ---
\## Signif. codes: 0 '***' 0.001 '**' 0.01 '*' 0.05 '.' 0.1 ' ' 1
\##
\## Zero-inflation model:
\##	Estimate Std. Error z value Pr(>|z|)
\## (Intercept)	-0.83882	0.03160 -26.546	<2e-16 ***
\## URBinary	-0.82464	0.04518 -18.252	<2e-16 ***
\## c_daterelevafter SaH	-1.44175	0.07414 -19.448	<2e-16 ***
\## c_daterelevduring SaH	-1.57486	0.08458 -18.619	<2e-16 ***
\## URBinary:c_daterelevafter SaH -13.90946 187.85997 -0.074	0.941
\## URBinary:c_daterelevduring SaH -16.20421 236.47785 -0.069	0.945
\## ---
\## Signif. codes: 0 '***' 0.001 '**' 0.01 '*' 0.05 '.' 0.1 ' ' 1

### Simulated Quantile Scaled Residual Plots

#### SimOut_lm1glmmrelev

SimOut_lm1glmmrelev <- simulateResiduals(fittedModel = lm1glmmrelev, plot = T)

\## DHARMa:plot used testOutliers with type = binomial for computational reasons (nObs > 500). Note that this method may not have inflated Type I error rates for integer-valued distributions. To get a more exact result, it is recommended to re-run testOutliers with type = 'bootstrap'. See ?testOutliers for details

plot(SimOut_lm1glmmrelev)

\## DHARMa:plot used testOutliers with type = binomial for computational reasons (nObs > 500). Note that this method may not have inflated Type I error rates for integer-valued distributions. To get a more exact result, it is recommended to re-run testOutliers with type = 'bootstrap'. See ?testOutliers for details

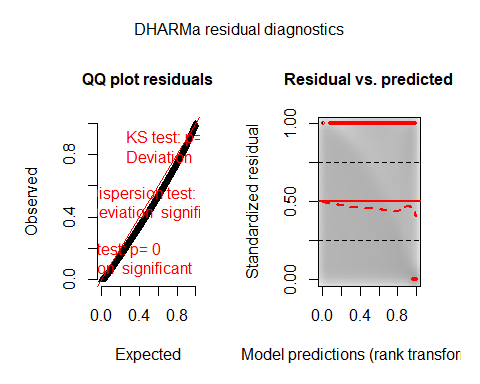

testZeroInflation(SimOut_lm1glmmrelev)

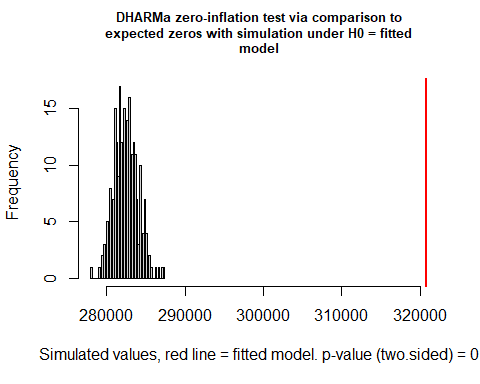

```
##
##  DHARMa zero-inflation test via comparison to expected zeros with
##  simulation under H0 = fitted model
##
## data:  simulationOutput
## ratioObsSim = 1.1354, p-value < 2.2e-16
## alternative hypothesis: two.sided
testOutliers(SimOut_lm1glmmrelev, type= 'bootstrap')
```

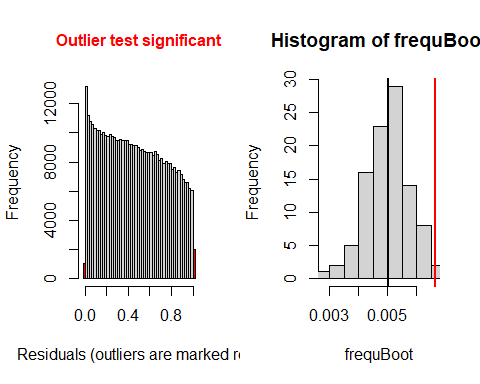

```
##
##  DHARMa bootstrapped outlier test
##
## data:  SimOut_lm1glmmrelev
## outliers at both margin(s) = 2959, observations = 446164, p-value <
## 2.2e-16
## alternative hypothesis: two.sided
##  percent confidence interval:
##  0.003519446 0.006204781
## sample estimates:
## outlier frequency (expected: 0.00500728431697761 )
##                                         0.00663209
simoutrecalc <- recalculateResiduals(SimOut_lm1glmmrelev, group = df_14$Date2)
testTemporalAutocorrelation(simoutrecalc, time = unique(df_14$Date2))
```

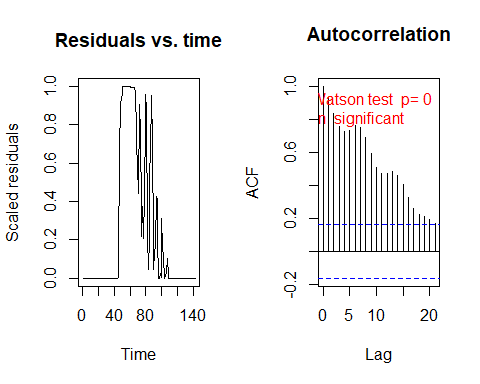

```
##
##  Durbin-Watson test
##
## data:  simulationOutput$scaledResiduals ~ 1
## DW = 0.12709, p-value < 2.2e-16
## alternative hypothesis: true autocorrelation is not 0
```

#### SimOut_lm2relev

SimOut_lm2relev <- simulateResiduals(fittedModel = lm2relev, plot = T)

\## DHARMa:plot used testOutliers with type = binomial for computational reasons (nObs > 500). Note that this method may not have inflated Type I error rates for integer-valued distributions. To get a more exact result, it is recommended to re-run testOutliers with type = 'bootstrap'. See ?testOutliers for details

plot(SimOut_lm2relev)

\## DHARMa:plot used testOutliers with type = binomial for computational reasons (nObs > 500). Note that this method may not have inflated Type I error rates for integer-valued distributions. To get a more exact result, it is recommended to re-run testOutliers with type = 'bootstrap'. See ?testOutliers for details

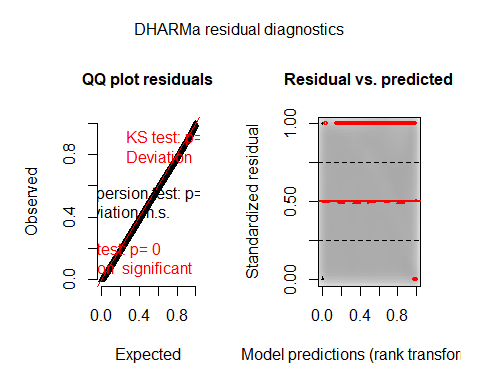

testZeroInflation(SimOut_lm2relev)

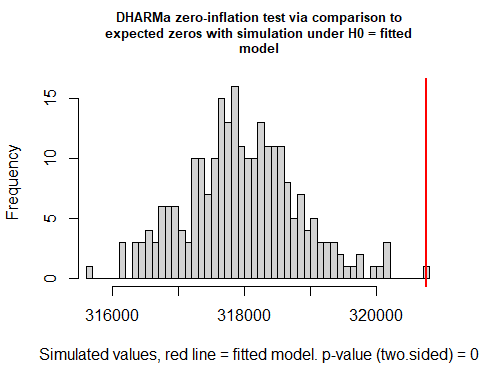

```
##
##  DHARMa zero-inflation test via comparison to expected zeros with
##  simulation under H0 = fitted model
##
## data:  simulationOutput
## ratioObsSim = 1.0088, p-value < 2.2e-16
## alternative hypothesis: two.sided
testOutliers(SimOut_lm2relev, type= 'bootstrap')
```

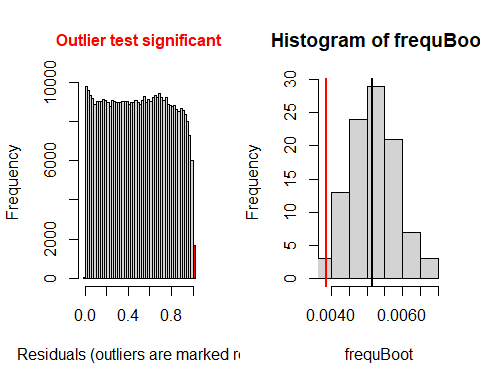

```
##
##  DHARMa bootstrapped outlier test
##
## data:  SimOut_lm2relev
## outliers at both margin(s) = 1716, observations = 446164, p-value =
## 0.04
## alternative hypothesis: two.sided
##  percent confidence interval:
##  0.003935997 0.006475029
## sample estimates:
## outlier frequency (expected: 0.00515005692973884 )
##                                        0.003846119
simoutrecalc <- recalculateResiduals(SimOut_lm2relev, group = df_14$Date2)
testTemporalAutocorrelation(simoutrecalc, time = unique(df_14$Date2))
```

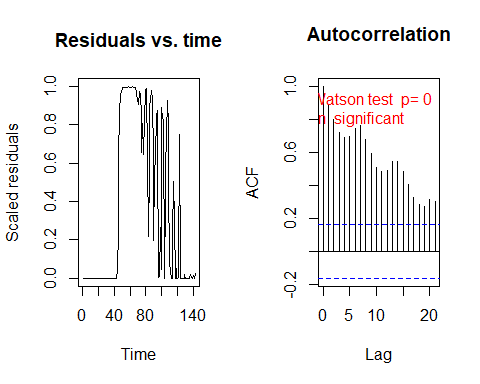

```
##
##  Durbin-Watson test
##
## data:  simulationOutput$scaledResiduals ~ 1
## DW = 0.20601, p-value < 2.2e-16
## alternative hypothesis: true autocorrelation is not 0
#


```

#### SimOut_lm2catziprelev

SimOut_lm2catziprelev <- simulateResiduals(fittedModel = lm2catziprelev, plot = T)

\## DHARMa:plot used testOutliers with type = binomial for computational reasons (nObs > 500). Note that this method may not have inflated Type I error rates for integer-valued distributions. To get a more exact result, it is recommended to re-run testOutliers with type = 'bootstrap'. See ?testOutliers for details

plot(SimOut_lm2catziprelev)

\## DHARMa:plot used testOutliers with type = binomial for computational reasons (nObs > 500). Note that this method may not have inflated Type I error rates for integer-valued distributions. To get a more exact result, it is recommended to re-run testOutliers with type = 'bootstrap'. See ?testOutliers for details

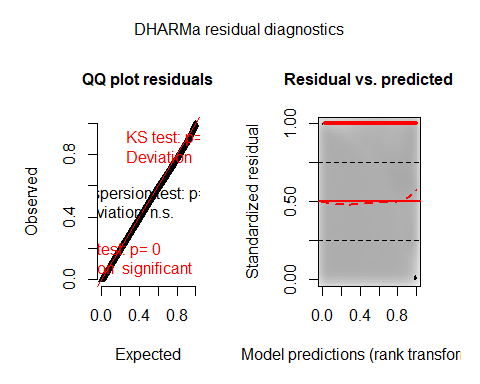

testZeroInflation(SimOut_lm2catziprelev)

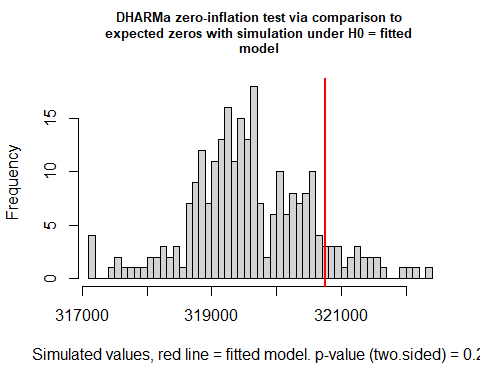

```
##
##  DHARMa zero-inflation test via comparison to expected zeros with
##  simulation under H0 = fitted model
##
## data:  simulationOutput
## ratioObsSim = 1.0037, p-value = 0.2
## alternative hypothesis: two.sided
testOutliers(SimOut_lm2catziprelev, type= 'bootstrap')
```

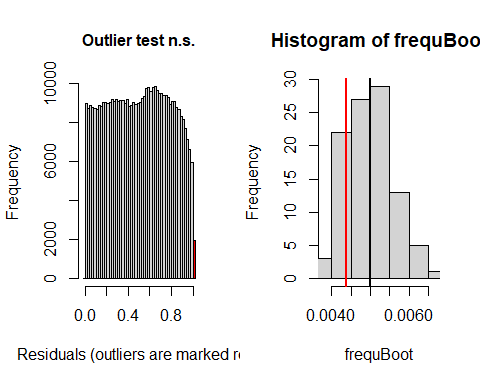

```
##
##  DHARMa bootstrapped outlier test
##
## data:  SimOut_lm2catziprelev
## outliers at both margin(s) = 1953, observations = 446164, p-value =
## 0.28
## alternative hypothesis: two.sided
##  percent confidence interval:
##  0.003989508 0.006183937
## sample estimates:
## outlier frequency (expected: 0.00499551734339839 )
##                                        0.004377314
simoutrecalc <- recalculateResiduals(SimOut_lm2catziprelev, group = df_14$Date2)
testTemporalAutocorrelation(simoutrecalc, time = unique(df_14$Date2))
```

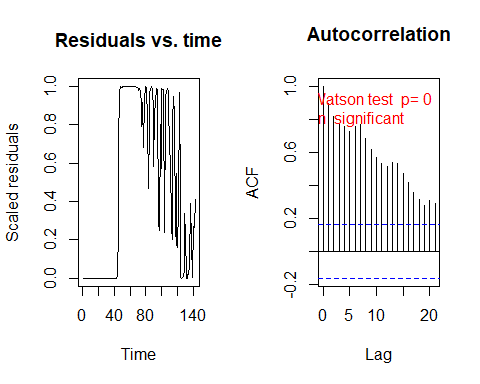

```
##
##  Durbin-Watson test
##
## data:  simulationOutput$scaledResiduals ~ 1
## DW = 0.19769, p-value < 2.2e-16
## alternative hypothesis: true autocorrelation is not 0
#


```

#### SimOut_lm2catziprelev_cs

SimOut_lm2catziprelev_cs <- simulateResiduals(fittedModel = lm2catziprelev_cs_cdate, plot = T)

\## DHARMa:plot used testOutliers with type = binomial for computational reasons (nObs > 500). Note that this method may not have inflated Type I error rates for integer-valued distributions. To get a more exact result, it is recommended to re-run testOutliers with type = 'bootstrap'. See ?testOutliers for details

plot(SimOut_lm2catziprelev_cs)

\## DHARMa:plot used testOutliers with type = binomial for computational reasons (nObs > 500). Note that this method may not have inflated Type I error rates for integer-valued distributions. To get a more exact result, it is recommended to re-run testOutliers with type = 'bootstrap'. See ?testOutliers for details

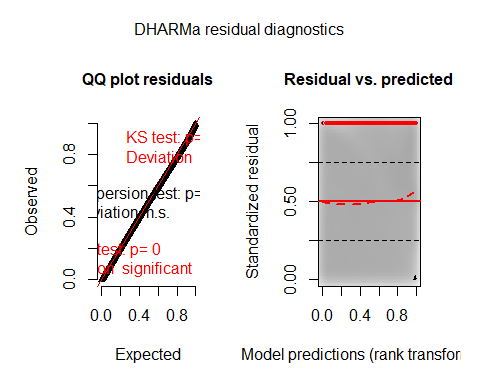

testZeroInflation(SimOut_lm2catziprelev_cs)

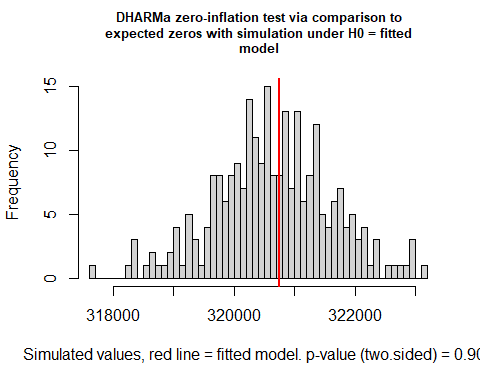

```
##
##  DHARMa zero-inflation test via comparison to expected zeros with
##  simulation under H0 = fitted model
##
## data:  simulationOutput
## ratioObsSim = 1.0003, p-value = 0.904
## alternative hypothesis: two.sided
testOutliers(SimOut_lm2catziprelev_cs, type= 'bootstrap')
```

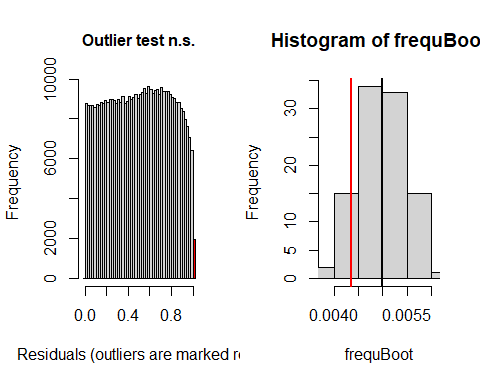

```
##
##  DHARMa bootstrapped outlier test
##
## data:  SimOut_lm2catziprelev_cs
## outliers at both margin(s) = 1943, observations = 446164, p-value =
## 0.22
## alternative hypothesis: two.sided
##  percent confidence interval:
##  0.004055235 0.005848858
## sample estimates:
## outlier frequency (expected: 0.00498182282748048 )
##                                        0.004354901
simoutrecalc <- recalculateResiduals(SimOut_lm2catziprelev_cs, group = df_14$Date2)
testTemporalAutocorrelation(simoutrecalc, time = unique(df_14$Date2))
```

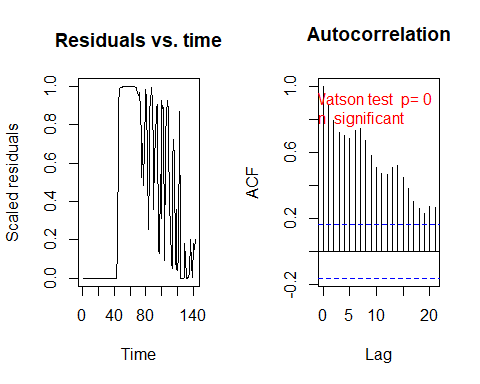

```
##
##  Durbin-Watson test
##
## data:  simulationOutput$scaledResiduals ~ 1
## DW = 0.22157, p-value < 2.2e-16
## alternative hypothesis: true autocorrelation is not 0
```

#### SimOut_lm2catziprelev_tpc

SimOut_lm2catziprelev_tpc <- simulateResiduals(fittedModel = lm2catziprelev_toep_cdate, plot = T)

\## DHARMa:plot used testOutliers with type = binomial for computational reasons (nObs > 500). Note that this method may not have inflated Type I error rates for integer-valued distributions. To get a more exact result, it is recommended to re-run testOutliers with type = 'bootstrap'. See ?testOutliers for details

plot(SimOut_lm2catziprelev_tpc)

\## DHARMa:plot used testOutliers with type = binomial for computational reasons (nObs > 500). Note that this method may not have inflated Type I error rates for integer-valued distributions. To get a more exact result, it is recommended to re-run testOutliers with type = 'bootstrap'. See ?testOutliers for details

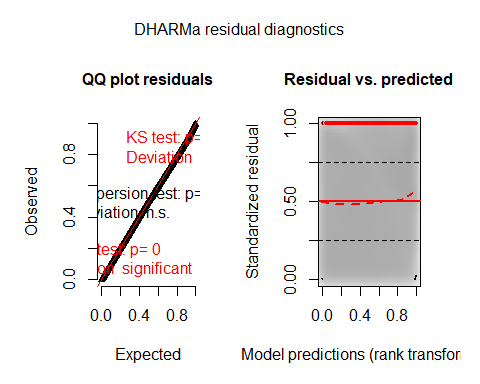

testZeroInflation(SimOut_lm2catziprelev_tpc)

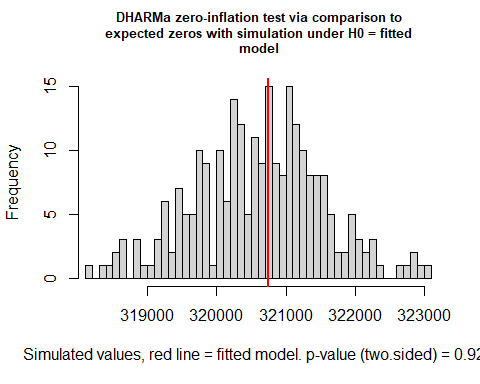

```
##
##  DHARMa zero-inflation test via comparison to expected zeros with
##  simulation under H0 = fitted model
##
## data:  simulationOutput
## ratioObsSim = 1.0004, p-value = 0.928
## alternative hypothesis: two.sided
testOutliers(SimOut_lm2catziprelev_tpc, type= 'bootstrap')
```

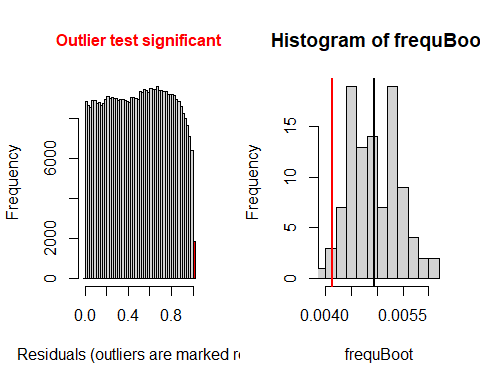

```
##
##  DHARMa bootstrapped outlier test
##
## data:  SimOut_lm2catziprelev_tpc
## outliers at both margin(s) = 1843, observations = 446164, p-value =
## 0.04
## alternative hypothesis: two.sided
##  percent confidence interval:
##  0.004147074 0.005899512
## sample estimates:
## outlier frequency (expected: 0.00493818416546382 )
##                                        0.004130768
simoutrecalc <- recalculateResiduals(SimOut_lm2catziprelev_tpc, group = df_14$Date2)
testTemporalAutocorrelation(simoutrecalc, time = unique(df_14$Date2))
```

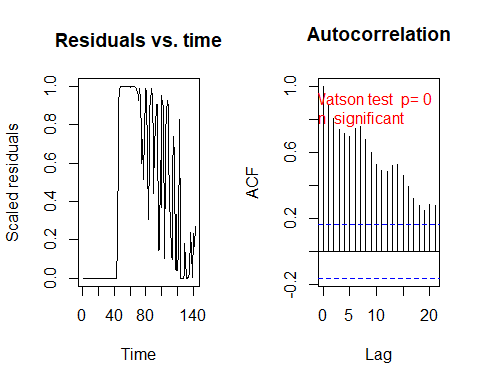

```
##
##  Durbin-Watson test
##
## data:  simulationOutput$scaledResiduals ~ 1
## DW = 0.20724, p-value < 2.2e-16
## alternative hypothesis: true autocorrelation is not 0
##

```

#### SimOut_lm2catziprelev_rsc

SimOut_lm2catziprelev_rsc <- simulateResiduals(fittedModel = lm2catziprelev_randslope_cdate, plot = T)

\## DHARMa:plot used testOutliers with type = binomial for computational reasons (nObs > 500). Note that this method may not have inflated Type I error rates for integer-valued distributions. To get a more exact result, it is recommended to re-run testOutliers with type = 'bootstrap'. See ?testOutliers for details

plot(SimOut_lm2catziprelev_rsc)

\## DHARMa:plot used testOutliers with type = binomial for computational reasons (nObs > 500). Note that this method may not have inflated Type I error rates for integer-valued distributions. To get a more exact result, it is recommended to re-run testOutliers with type = 'bootstrap'. See ?testOutliers for details

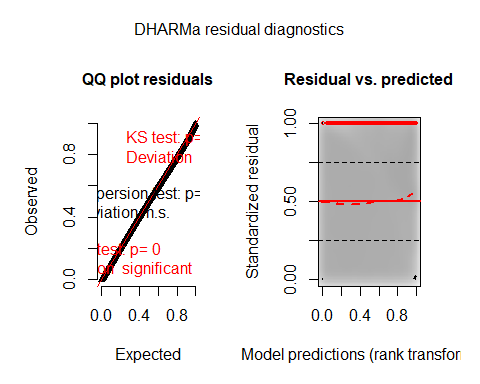

testZeroInflation(SimOut_lm2catziprelev_rsc)

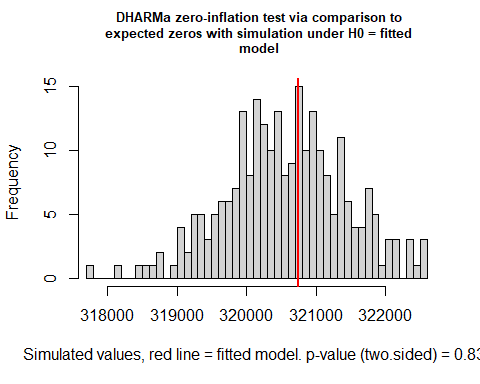

```
##
##  DHARMa zero-inflation test via comparison to expected zeros with
##  simulation under H0 = fitted model
##
## data:  simulationOutput
## ratioObsSim = 1.0006, p-value = 0.832
## alternative hypothesis: two.sided
testOutliers(SimOut_lm2catziprelev_rsc, type= 'bootstrap')
```

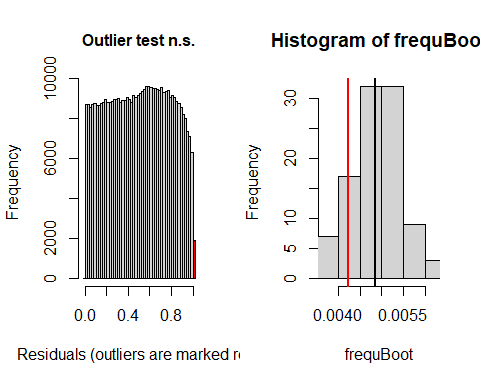

```
##
##  DHARMa bootstrapped outlier test
##
## data:  SimOut_lm2catziprelev_rsc
## outliers at both margin(s) = 1886, observations = 446164, p-value =
## 0.28
## alternative hypothesis: two.sided
##  percent confidence interval:
##  0.003860296 0.005998691
## sample estimates:
## outlier frequency (expected: 0.00486076868595404 )
##                                        0.004227145
simoutrecalc <- recalculateResiduals(SimOut_lm2catziprelev_rsc, group = df_14$Date2)
testTemporalAutocorrelation(simoutrecalc, time = unique(df_14$Date2))
```

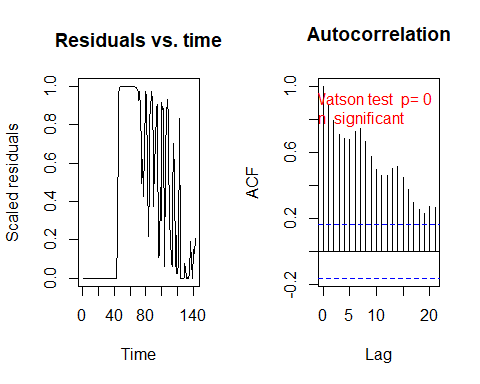

```
##
##  Durbin-Watson test
##
## data:  simulationOutput$scaledResiduals ~ 1
## DW = 0.21202, p-value < 2.2e-16
## alternative hypothesis: true autocorrelation is not 0
```

#### SimOut_lm2catziprelev_rsd

SimOut_lm2catziprelev_rsd <- simulateResiduals(fittedModel = lm2catziprelev_randslope_date2, plot = T)

\## DHARMa:plot used testOutliers with type = binomial for computational reasons (nObs > 500). Note that this method may not have inflated Type I error rates for integer-valued distributions. To get a more exact result, it is recommended to re-run testOutliers with type = 'bootstrap'. See ?testOutliers for details

plot(SimOut_lm2catziprelev_rsd)

\## DHARMa:plot used testOutliers with type = binomial for computational reasons (nObs > 500). Note that this method may not have inflated Type I error rates for integer-valued distributions. To get a more exact result, it is recommended to re-run testOutliers with type = 'bootstrap'. See ?testOutliers for details

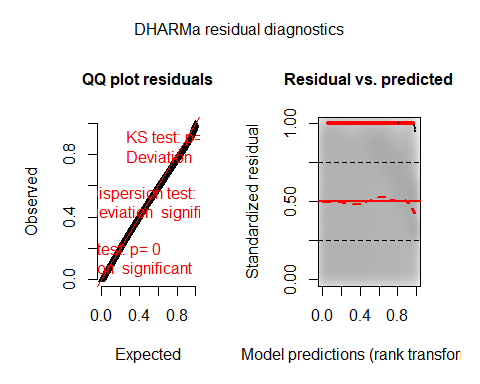

testZeroInflation(SimOut_lm2catziprelev_rsd)

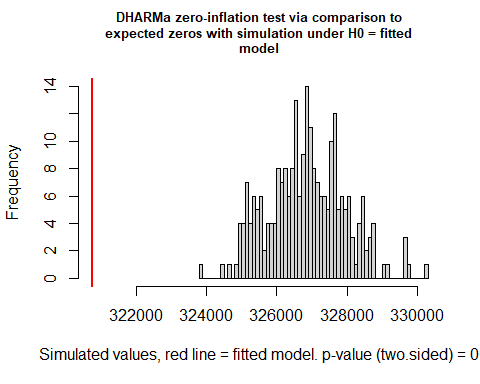

```
##
##  DHARMa zero-inflation test via comparison to expected zeros with
##  simulation under H0 = fitted model
##
## data:  simulationOutput
## ratioObsSim = 0.98132, p-value < 2.2e-16
## alternative hypothesis: two.sided
testOutliers(SimOut_lm2catziprelev_rsd, type= 'bootstrap')
```

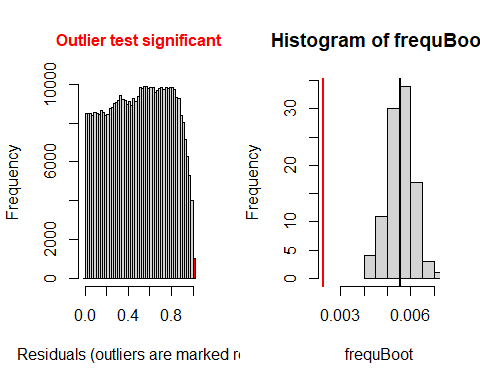

```
##
##  DHARMa bootstrapped outlier test
##
## data:  SimOut_lm2catziprelev_rsd
## outliers at both margin(s) = 1002, observations = 446164, p-value <
## 2.2e-16
## alternative hypothesis: two.sided
##  percent confidence interval:
##  0.004446179 0.006592979
## sample estimates:
## outlier frequency (expected: 0.00556387337391632 )
##                                        0.002245811
simoutrecalc <- recalculateResiduals(SimOut_lm2catziprelev_rsd, group = df_14$Date2)
testTemporalAutocorrelation(simoutrecalc, time = unique(df_14$Date2))
```

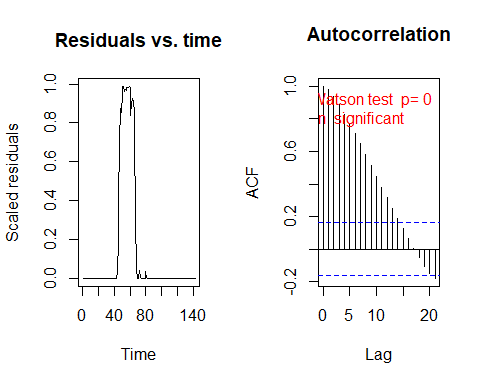

```
##
##  Durbin-Watson test
##
## data:  simulationOutput$scaledResiduals ~ 1
## DW = 0.034811, p-value < 2.2e-16
## alternative hypothesis: true autocorrelation is not 0
###

```

#### SimOut_lm3glmmrelev

SimOut_lm3glmmrelev <- simulateResiduals(fittedModel = lm3glmmrelev, plot = T)

\## DHARMa:plot used testOutliers with type = binomial for computational reasons (nObs > 500). Note that this method may not have inflated Type I error rates for integer-valued distributions. To get a more exact result, it is recommended to re-run testOutliers with type = 'bootstrap'. See ?testOutliers for details

plot(SimOut_lm3glmmrelev)

\## DHARMa:plot used testOutliers with type = binomial for computational reasons (nObs > 500). Note that this method may not have inflated Type I error rates for integer-valued distributions. To get a more exact result, it is recommended to re-run testOutliers with type = 'bootstrap'. See ?testOutliers for details

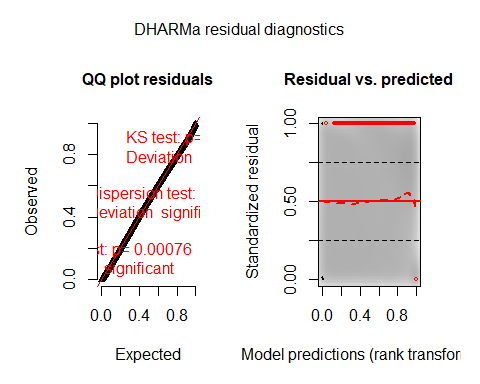

testZeroInflation(SimOut_lm3glmmrelev)

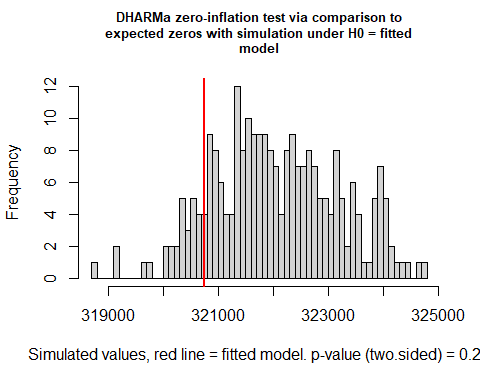

```
##
##  DHARMa zero-inflation test via comparison to expected zeros with
##  simulation under H0 = fitted model
##
## data:  simulationOutput
## ratioObsSim = 0.99595, p-value = 0.24
## alternative hypothesis: two.sided
testOutliers(SimOut_lm3glmmrelev, type= 'bootstrap')
```

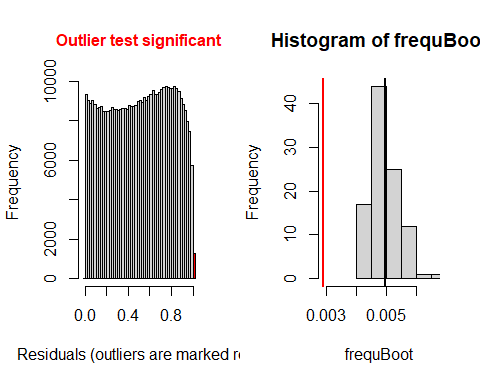

```
##
##  DHARMa bootstrapped outlier test
##
## data:  SimOut_lm3glmmrelev
## outliers at both margin(s) = 1286, observations = 446164, p-value <
## 2.2e-16
## alternative hypothesis: two.sided
##  percent confidence interval:
##  0.004172513 0.005960925
## sample estimates:
## outlier frequency (expected: 0.00496371289480998 )
##                                        0.002882348
simoutrecalc <- recalculateResiduals(SimOut_lm3glmmrelev, group = df_14$Date2)
testTemporalAutocorrelation(simoutrecalc, time = unique(df_14$Date2))
```

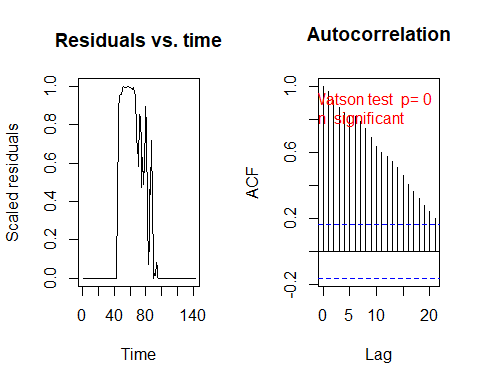

```
##
##  Durbin-Watson test
##
## data:  simulationOutput$scaledResiduals ~ 1
## DW = 0.061153, p-value < 2.2e-16
## alternative hypothesis: true autocorrelation is not 0
```

#### SimOut_lm3glmmRandSlope

SimOut_lm3glmmRandSlope <- simulateResiduals(fittedModel = lm3glmmRandslope, plot = T)

\## DHARMa:plot used testOutliers with type = binomial for computational reasons (nObs > 500). Note that this method may not have inflated Type I error rates for integer-valued distributions. To get a more exact result, it is recommended to re-run testOutliers with type = 'bootstrap'. See ?testOutliers for details

plot(SimOut_lm3glmmRandSlope)

\## DHARMa:plot used testOutliers with type = binomial for computational reasons (nObs > 500). Note that this method may not have inflated Type I error rates for integer-valued distributions. To get a more exact result, it is recommended to re-run testOutliers with type = 'bootstrap'. See ?testOutliers for details

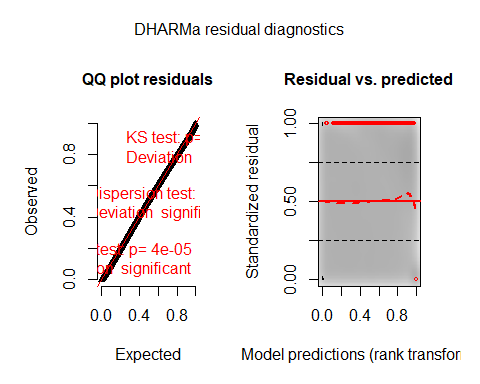

testZeroInflation(SimOut_lm3glmmRandSlope)

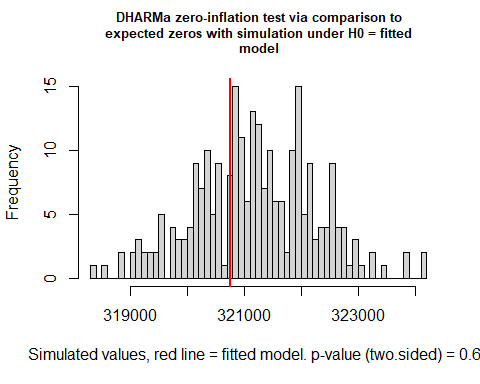

```
##
##  DHARMa zero-inflation test via comparison to expected zeros with
##  simulation under H0 = fitted model
##
## data:  simulationOutput
## ratioObsSim = 0.99862, p-value = 0.64
## alternative hypothesis: two.sided
testOutliers(SimOut_lm3glmmRandSlope, type= 'bootstrap')
```

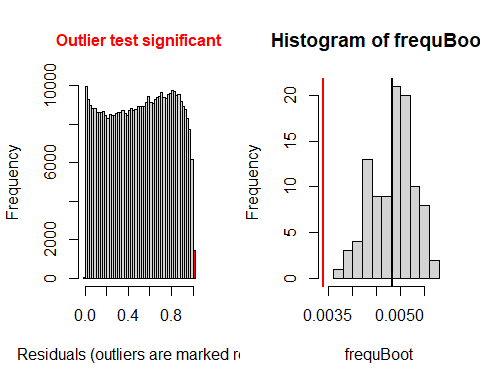

```
##
##  DHARMa bootstrapped outlier test
##
## data:  SimOut_lm3glmmRandSlope
## outliers at both margin(s) = 1508, observations = 446164, p-value <
## 2.2e-16
## alternative hypothesis: two.sided
##  percent confidence interval:
##  0.003982504 0.005508569
## sample estimates:
## outlier frequency (expected: 0.0048313848719305 )
##                                       0.003379923
simoutrecalc <- recalculateResiduals(SimOut_lm3glmmRandSlope, group = df_14$Date2)
testTemporalAutocorrelation(simoutrecalc, time = unique(df_14$Date2))
```

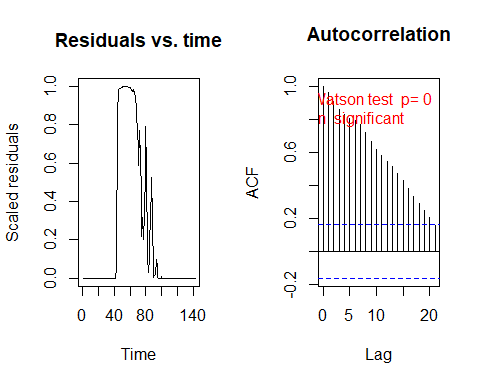

```
##
##  Durbin-Watson test
##
## data:  simulationOutput$scaledResiduals ~ 1
## DW = 0.072204, p-value < 2.2e-16
## alternative hypothesis: true autocorrelation is not 0
##

```

#### SimOut_lm4catziprelev

SimOut_lm4catziprelev <- simulateResiduals(fittedModel = lm4catziprelev, plot = T)

\## DHARMa:plot used testOutliers with type = binomial for computational reasons (nObs > 500). Note that this method may not have inflated Type I error rates for integer-valued distributions. To get a more exact result, it is recommended to re-run testOutliers with type = 'bootstrap'. See ?testOutliers for details

plot(SimOut_lm4catziprelev)

\## DHARMa:plot used testOutliers with type = binomial for computational reasons (nObs > 500). Note that this method may not have inflated Type I error rates for integer-valued distributions. To get a more exact result, it is recommended to re-run testOutliers with type = 'bootstrap'. See ?testOutliers for details

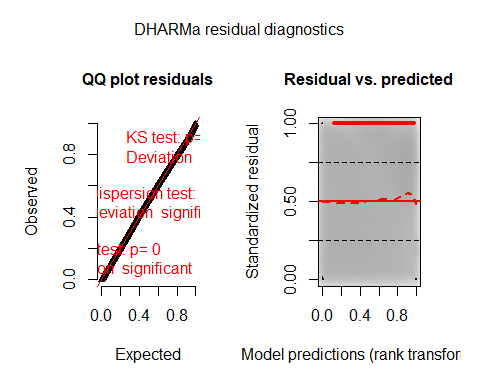

testZeroInflation(SimOut_lm4catziprelev)

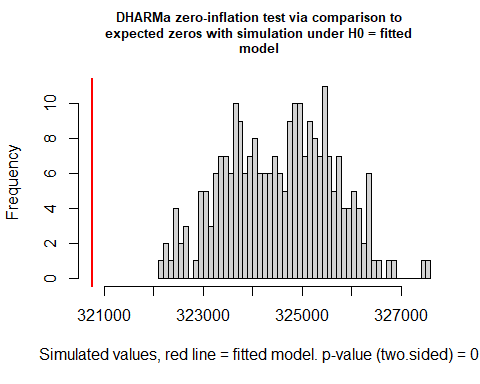

```
##
##  DHARMa zero-inflation test via comparison to expected zeros with
##  simulation under H0 = fitted model
##
## data:  simulationOutput
## ratioObsSim = 0.9883, p-value < 2.2e-16
## alternative hypothesis: two.sided
testOutliers(SimOut_lm4catziprelev, type= 'bootstrap')
```

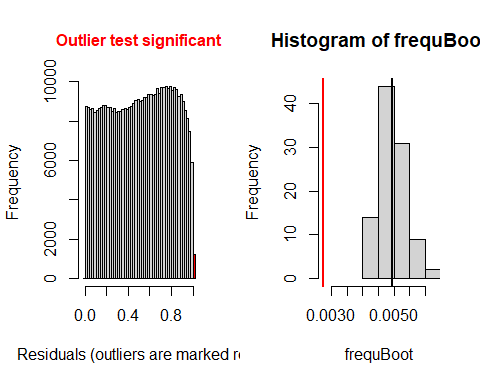

```
##
##  DHARMa bootstrapped outlier test
##
## data:  SimOut_lm4catziprelev
## outliers at both margin(s) = 1219, observations = 446164, p-value <
## 2.2e-16
## alternative hypothesis: two.sided
##  percent confidence interval:
##  0.004161026 0.005833057
## sample estimates:
## outlier frequency (expected: 0.00496077675473593 )
##                                        0.002732179
simoutrecalc <- recalculateResiduals(SimOut_lm4catziprelev, group = df_14$Date2)
testTemporalAutocorrelation(simoutrecalc, time = unique(df_14$Date2))
```

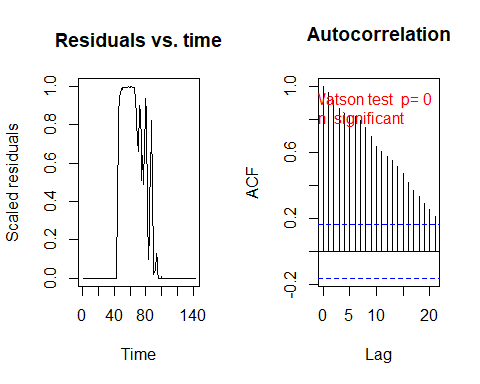

```
##
##  Durbin-Watson test
##
## data:  simulationOutput$scaledResiduals ~ 1
## DW = 0.063356, p-value < 2.2e-16
## alternative hypothesis: true autocorrelation is not 0
```

### Removing Outliers

#### Removing Outliers lm3glmm

```
# REMOVING OUTLIERS LM3GLMM
r <- which(residuals(SimOut_lm3glmmrelev) == 1 | residuals(SimOut_lm3glmmrelev) == 0)

df_14$row <- c(1:446164)

`%notin%` <- Negate(`%in%`)
#removing the counties
outcount <- df_14$c_FIPS[df_14$row %in% r]
outcount <- unique(outcount)
df_14outremcount <- df_14[df_14$c_FIPS %notin% outcount,]

# REMOVING OUTLIERS LM3GLMM
summary(lm3glmmrelevoutcount)##  Family: nbinom2  ( log )
## Formula:
## newcase_nst_14 ~ offset(popoff) + URBinary * c_daterelev + URBinary *
##     Date2 + URBinary * dsahcarried + URBinary * asahcarried +      (1 | c_FIPS)
## Data: df_14outremcount
##
##       AIC       BIC    logLik  deviance  df.resid
##  810243.1  810395.5 -405107.6  810215.1    393752
##
## Random effects:
##
## Conditional model:
##  Groups Name        Variance Std.Dev.
##  c_FIPS (Intercept) 1.471    1.213
## Number of obs: 393766, groups:  c_FIPS, 2773
##
## Overdispersion parameter for nbinom2 family (): 0.712
##
## Conditional model:
##                                  Estimate Std. Error z value Pr(>|z|)
## (Intercept)                    -3.6664114  0.0415560  -88.23  < 2e-16 ***
## URBinary                       -1.7572238  0.0635405  -27.66  < 2e-16 ***
## c_daterelevafter SaH            0.1984690  0.0310583    6.39 1.66e-10 ***
## c_daterelevduring SaH           0.4720351  0.0200622   23.53  < 2e-16 ***
## Date2                           0.0461757  0.0004144  111.42  < 2e-16 ***
## dsahcarried                    -0.0375113  0.0005965  -62.89  < 2e-16 ***
## asahcarried                    -0.0150326  0.0009235  -16.28  < 2e-16 ***
## URBinary:c_daterelevafter SaH  -0.9250872  0.0403586  -22.92  < 2e-16 ***
## URBinary:c_daterelevduring SaH -0.6737951  0.0257428  -26.17  < 2e-16 ***
## URBinary:Date2                  0.0504486  0.0006895   73.17  < 2e-16 ***
## URBinary:dsahcarried           -0.0517760  0.0008711  -59.44  < 2e-16 ***
## URBinary:asahcarried           -0.0503087  0.0013246  -37.98  < 2e-16 ***
## ---
## Signif. codes:  0 '***' 0.001 '**' 0.01 '*' 0.05 '.' 0.1 ' ' 1
SimOut_lm3glmmrelevoutcount <- simulateResiduals(fittedModel = lm3glmmrelevoutcount, plot = T)
## DHARMa:plot used testOutliers with type = binomial for computational reasons (nObs > 500). Note that this method may not have inflated Type I error rates for integer-valued distributions. To get a more exact result, it is recommended to re-run testOutliers with type = 'bootstrap'. See ?testOutliers for details
plot(SimOut_lm3glmmrelevoutcount)
## DHARMa:plot used testOutliers with type = binomial for computational reasons (nObs > 500). Note that this method may not have inflated Type I error rates for integer-valued distributions. To get a more exact result, it is recommended to re-run testOutliers with type = 'bootstrap'. See ?testOutliers for details
```

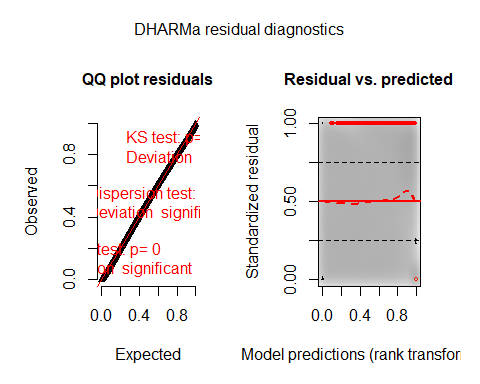

testZeroInflation(SimOut_lm3glmmrelevoutcount)

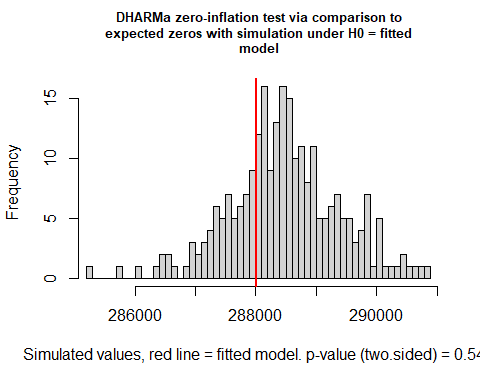

```
##
##  DHARMa zero-inflation test via comparison to expected zeros with
##  simulation under H0 = fitted model
##
## data:  simulationOutput
## ratioObsSim = 0.99841, p-value = 0.544
## alternative hypothesis: two.sided
testOutliers(SimOut_lm3glmmrelevoutcount, type= 'bootstrap')
```

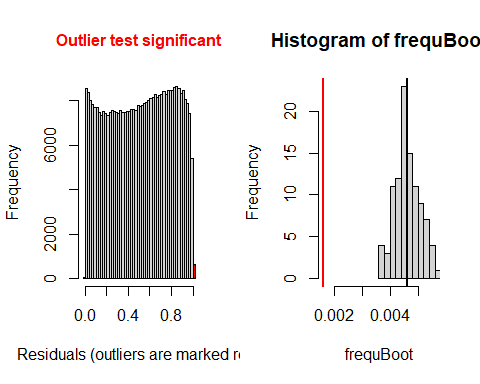

```
##
##  DHARMa bootstrapped outlier test
##
## data:  SimOut_lm3glmmrelevoutcount
## outliers at both margin(s) = 636, observations = 393766, p-value <
## 2.2e-16
## alternative hypothesis: two.sided
##  percent confidence interval:
##  0.003774640 0.005446128
## sample estimates:
## outlier frequency (expected: 0.00462145030297182 )
##                                        0.001615172
simoutrecalc <- recalculateResiduals(SimOut_lm3glmmrelevoutcount, group = df_14outremcount$Date2)
testTemporalAutocorrelation(simoutrecalc, time = unique(df_14outremcount$Date2))
```

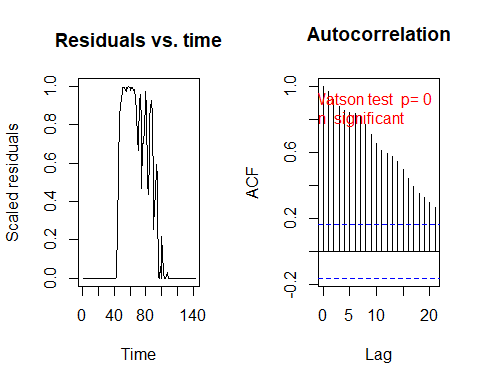

```
##
##  Durbin-Watson test
##
## data:  simulationOutput$scaledResiduals ~ 1
## DW = 0.055966, p-value < 2.2e-16
## alternative hypothesis: true autocorrelation is not 0
```

#### *Removing Outliers Random Slope lm3glmmRandSlope*

```

r <- which(residuals(SimOut_lm3glmmRandSlope) == 1 | residuals(SimOut_lm3glmmRandSlope) == 0)

df_14$row <- c(1:446164)

`%notin%` <- Negate(`%in%`)
#removing the counties
outcount <- df_14$c_FIPS[df_14$row %in% r]
outcount <- unique(outcount)
df_14outremcountrand <- df_14[df_14$c_FIPS %notin% outcount,]


# REMOVING OUTLIERS RANDOM SLOPE
summary(lm3glmmrelevrandslopeoutcount)
##  Family: nbinom2  ( log )
## Formula:
## newcase_nst_14 ~ offset(popoff) + URBinary * c_daterelev + URBinary *
##     Date2 + URBinary * dsahcarried + URBinary * asahcarried +      (1 | c_FIPS)
## Data: df_14outremcountrand
##
##       AIC       BIC    logLik  deviance  df.resid
##  772804.9  772956.9 -386388.5  772776.9    383102
##
## Random effects:
##
## Conditional model:
##  Groups Name        Variance Std.Dev.
##  c_FIPS (Intercept) 1.513    1.23
## Number of obs: 383116, groups:  c_FIPS, 2698
##
## Overdispersion parameter for nbinom2 family (): 0.751
##
## Conditional model:
##                                  Estimate Std. Error z value Pr(>|z|)
## (Intercept)                    -3.7633293  0.0428977  -87.73   <2e-16 ***
## URBinary                       -2.1760163  0.0664835  -32.73   <2e-16 ***
## c_daterelevafter SaH            0.3306234  0.0317482   10.41   <2e-16 ***
## c_daterelevduring SaH           0.5825412  0.0206052   28.27   <2e-16 ***
## Date2                           0.0462531  0.0004294  107.72   <2e-16 ***
## dsahcarried                    -0.0387154  0.0006110  -63.37   <2e-16 ***
## asahcarried                    -0.0154415  0.0009454  -16.33   <2e-16 ***
## URBinary:c_daterelevafter SaH  -1.2598501  0.0409353  -30.78   <2e-16 ***
## URBinary:c_daterelevduring SaH -0.9889043  0.0264444  -37.40   <2e-16 ***
## URBinary:Date2                  0.0610684  0.0007481   81.63   <2e-16 ***
## URBinary:dsahcarried           -0.0609167  0.0009188  -66.30   <2e-16 ***
## URBinary:asahcarried           -0.0600529  0.0013593  -44.18   <2e-16 ***
## ---
## Signif. codes:  0 '***' 0.001 '**' 0.01 '*' 0.05 '.' 0.1 ' ' 1
SimOut_lm3glmmrelevoutcountrandslope <- simulateResiduals(fittedModel = lm3glmmrelevrandslopeoutcount, plot = T)
## DHARMa:plot used testOutliers with type = binomial for computational reasons (nObs > 500). Note that this method may not have inflated Type I error rates for integer-valued distributions. To get a more exact result, it is recommended to re-run testOutliers with type = 'bootstrap'. See ?testOutliers for details
plot(SimOut_lm3glmmrelevoutcountrandslope)
## DHARMa:plot used testOutliers with type = binomial for computational reasons (nObs > 500). Note that this method may not have inflated Type I error rates for integer-valued distributions. To get a more exact result, it is recommended to re-run testOutliers with type = 'bootstrap'. See ?testOutliers for details
```

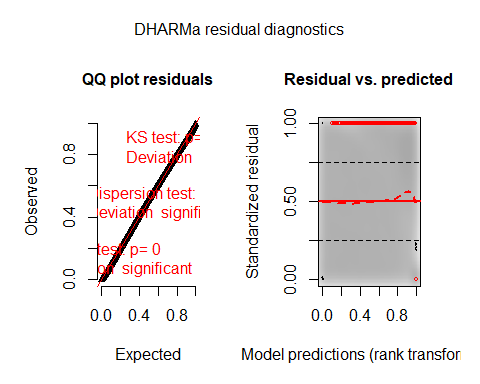

testZeroInflation(SimOut_lm3glmmrelevoutcountrandslope)

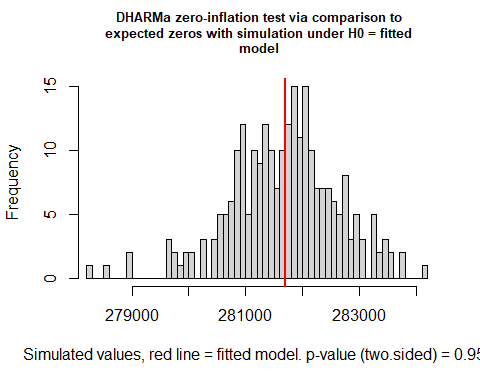

```
##
##  DHARMa zero-inflation test via comparison to expected zeros with
##  simulation under H0 = fitted model
##
## data:  simulationOutput
## ratioObsSim = 1, p-value = 0.952
## alternative hypothesis: two.sided
testOutliers(SimOut_lm3glmmrelevoutcountrandslope, type= 'bootstrap')
```

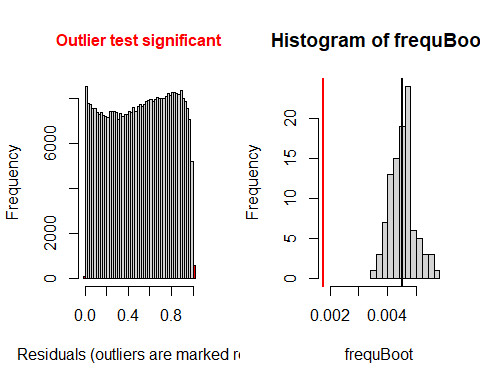

```
##
##  DHARMa bootstrapped outlier test
##
## data:  SimOut_lm3glmmrelevoutcountrandslope
## outliers at both margin(s) = 675, observations = 383116, p-value <
## 2.2e-16
## alternative hypothesis: two.sided
##  percent confidence interval:
##  0.003781622 0.005431057
## sample estimates:
## outlier frequency (expected: 0.00450790361143883 )
##                                        0.001761868
simoutrecalc <- recalculateResiduals(SimOut_lm3glmmrelevoutcountrandslope, group = df_14outremcountrand$Date2)
testTemporalAutocorrelation(simoutrecalc, time = unique(df_14outremcountrand$Date2))
```

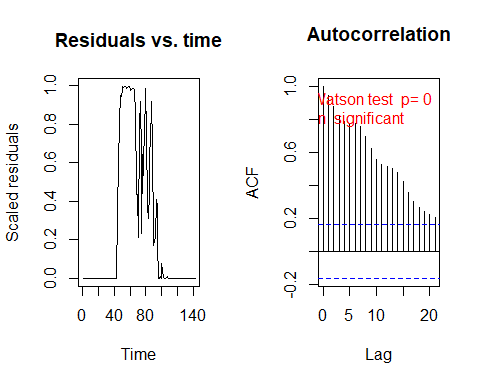

```
##
##  Durbin-Watson test
##
## data:  simulationOutput$scaledResiduals ~ 1
## DW = 0.10231, p-value < 2.2e-16
## alternative hypothesis: true autocorrelation is not 0
```

### Sensitivity Analysis

We conducted a sensitivity analysis surrounding the lag time between daily new cases and time reported. The dependent variable, daily new cases, in this case must be lagged for proper analysis because of several reasons. First, it is well known that the potential incubation period for SARS-CoV-2 is upwards of 14 days, which would imply that an individual being tested positive for the virus could have been exposed to the virus some two weeks earlier, potentially placing them out of range of a particular stay-at-home order, and thus complicating analysis. Second, while stay-at-home orders are declared and in place, it takes time for the orders to be adhered to and enforced for a measurable effect. We initially used the longer 14-day lag due to its being the incubation period. However, other studies have utilized five-to-ten-day lags. Therefore, it becomes necessary to conduct sensitivity analysis, the result of which we report below.

Results of the sensitivity analysis did not change any of the study inferences of conclusions. Moreover, the five-day and ten-day lag analyses exhibited significant overdispersion and zero-inflation, whereas the 14-day lag does not exhibit these characteristics

#### Five-Day Lag

```
#reads in data
setwd("C:\\Users\\Jake\\Desktop\\MAYO\\COVID RURALITY")
df_14 <- read.csv("df_14.csv",header=T)

#installs packages then loads them into the session
library(glmmTMB)
## Warning: package 'glmmTMB' was built under R version 3.6.3
library(DHARMa)
## Warning: package 'DHARMa' was built under R version 3.6.3
## This is DHARMa 0.3.3.0. For overview type '?DHARMa'. For recent changes, type news(package = 'DHARMa') Note: Syntax of plotResiduals has changed in 0.3.0, see ?plotResiduals for details
# Releveling
df_14$c_daterelev <- relevel(df_14$c_date, ref = "before SaH")


#Five Day Lag
n <- 142
D <- 5
for (i in 1:n){
  df_14$newcase_nst_5[df_14$Date2 == i] <- ifelse( i > (n-D), df_14$newcase_nst_14[df_14$Date2 == (i-(14-D))], df_14$newcase_nst[df_14$Date2 == (i+D)])
}

#RENAMING THE VARIABLE TO ALLOW the implementation of the lag

df_14$newcase_nst_14 <- df_14$newcase_nst_5


load("C:/Users/Jake/Desktop/MAYO/COVID RURALITY/5Day.RData")
###########################################
############# SUMMARY RESULTS #############
###########################################

# GLMMTMB mixed effects poisson model
summary(lm1glmmrelev)
##  Family: poisson  ( log )
## Formula:
## newcase_nst_14 ~ offset(popoff) + URBinary * c_daterelev + URBinary *
##     Date2 + URBinary * dsahcarried + URBinary * asahcarried +      (1 | c_FIPS)
## Data: df_14
##
##       AIC       BIC    logLik  deviance  df.resid
## 1433084.9 1433228.0 -716529.5 1433058.9    446151
##
## Random effects:
##
## Conditional model:
##  Groups Name        Variance Std.Dev.
##  c_FIPS (Intercept) 1.533    1.238
## Number of obs: 446164, groups:  c_FIPS, 3142
##
## Conditional model:
##                                  Estimate Std. Error z value Pr(>|z|)
## (Intercept)                    -1.7870967  0.0294820   -60.6  < 2e-16 ***
## URBinary                       -2.1394090  0.0494705   -43.2  < 2e-16 ***
## c_daterelevafter SaH            0.8655903  0.0109259    79.2  < 2e-16 ***
## c_daterelevduring SaH           0.6655302  0.0096345    69.1  < 2e-16 ***
## Date2                           0.0438043  0.0001130   387.5  < 2e-16 ***
## dsahcarried                    -0.0055943  0.0001749   -32.0  < 2e-16 ***
## asahcarried                    -0.0153344  0.0001853   -82.8  < 2e-16 ***
## URBinary:c_daterelevafter SaH  -0.1359608  0.0175846    -7.7 1.06e-14 ***
## URBinary:c_daterelevduring SaH -0.0504590  0.0156094    -3.2  0.00123 **
## URBinary:Date2                  0.0044700  0.0002710    16.5  < 2e-16 ***
## URBinary:dsahcarried           -0.0038140  0.0003344   -11.4  < 2e-16 ***
## URBinary:asahcarried           -0.0037971  0.0003657   -10.4  < 2e-16 ***
## ---
## Signif. codes:  0 '***' 0.001 '**' 0.01 '*' 0.05 '.' 0.1 ' ' 1
# Zero inflated poisson mixed effects (zero inflated using the whole formula)
summary(lm2relev)
##  Family: poisson  ( log )
## Formula:
## newcase_nst_14 ~ offset(popoff) + URBinary * c_daterelev + URBinary *
##     Date2 + URBinary * dsahcarried + URBinary * asahcarried +      (1 | c_FIPS)
## Zero inflation:
## ~URBinary * c_daterelev + URBinary * Date2 + URBinary * dsahcarried +
##     URBinary * asahcarried
## Data: df_14
##
##       AIC       BIC    logLik  deviance  df.resid
## 1310314.5 1310589.7 -655132.2 1310264.5    446139
##
## Random effects:
##
## Conditional model:
##  Groups Name        Variance Std.Dev.
##  c_FIPS (Intercept) 1.419    1.191
## Number of obs: 446164, groups:  c_FIPS, 3142
##
## Conditional model:
##                                  Estimate Std. Error z value Pr(>|z|)
## (Intercept)                     0.6575298  0.0299703   21.94  < 2e-16 ***
## URBinary                       -1.9027802  0.0515287  -36.93  < 2e-16 ***
## c_daterelevafter SaH            0.0031825  0.0123837    0.26 0.797184
## c_daterelevduring SaH          -0.1871174  0.0111842  -16.73  < 2e-16 ***
## Date2                           0.0279289  0.0001427  195.73  < 2e-16 ***
## dsahcarried                    -0.0014594  0.0002064   -7.07 1.55e-12 ***
## asahcarried                    -0.0051106  0.0002063  -24.77  < 2e-16 ***
## URBinary:c_daterelevafter SaH  -0.0664906  0.0197790   -3.36 0.000775 ***
## URBinary:c_daterelevduring SaH  0.0078653  0.0179242    0.44 0.660797
## URBinary:Date2                  0.0011929  0.0003352    3.56 0.000373 ***
## URBinary:dsahcarried           -0.0002050  0.0003994   -0.51 0.607717
## URBinary:asahcarried           -0.0005837  0.0004180   -1.40 0.162626
## ---
## Signif. codes:  0 '***' 0.001 '**' 0.01 '*' 0.05 '.' 0.1 ' ' 1
##
## Zero-inflation model:
##                                  Estimate Std. Error z value Pr(>|z|)
## (Intercept)                     5.9091552  0.0352677  167.55  < 2e-16 ***
## URBinary                        0.2288969  0.0632392    3.62 0.000295 ***
## c_daterelevafter SaH            0.3896047  0.0461149    8.45  < 2e-16 ***
## c_daterelevduring SaH           0.1130996  0.0207635    5.45 5.12e-08 ***
## Date2                          -0.0795411  0.0005083 -156.47  < 2e-16 ***
## dsahcarried                     0.0072807  0.0007160   10.17  < 2e-16 ***
## asahcarried                    -0.0222593  0.0027812   -8.00 1.21e-15 ***
## URBinary:c_daterelevafter SaH   0.0775074  0.0791344    0.98 0.327362
## URBinary:c_daterelevduring SaH  0.1038960  0.0352424    2.95 0.003198 **
## URBinary:Date2                 -0.0050061  0.0009762   -5.13 2.93e-07 ***
## URBinary:dsahcarried            0.0041206  0.0011598    3.55 0.000381 ***
## URBinary:asahcarried            0.0017428  0.0049499    0.35 0.724776
## ---
## Signif. codes:  0 '***' 0.001 '**' 0.01 '*' 0.05 '.' 0.1 ' ' 1
# Zero inflated poisson mixed effects (zero inflated using the rurality and dates)
summary(lm2catziprelev)
##  Family: poisson  ( log )
## Formula:
## newcase_nst_14 ~ offset(popoff) + URBinary * c_daterelev + URBinary *
##     Date2 + URBinary * dsahcarried + URBinary * asahcarried +      (1 | c_FIPS)
## Zero inflation:                  ~URBinary * c_daterelev
## Data: df_14
##
##       AIC       BIC    logLik  deviance  df.resid
## 1385307.0 1385516.1 -692634.5 1385269.0    446145
##
## Random effects:
##
## Conditional model:
##  Groups Name        Variance Std.Dev.
##  c_FIPS (Intercept) 1.546    1.243
## Number of obs: 446164, groups:  c_FIPS, 3142
##
## Conditional model:
##                                  Estimate Std. Error z value Pr(>|z|)
## (Intercept)                    -1.3991715  0.0305346  -45.82  < 2e-16 ***
## URBinary                       -1.7737967  0.0540949  -32.79  < 2e-16 ***
## c_daterelevafter SaH            1.1083193  0.0122324   90.61  < 2e-16 ***
## c_daterelevduring SaH           0.9587011  0.0109879   87.25  < 2e-16 ***
## Date2                           0.0404765  0.0001284  315.14  < 2e-16 ***
## dsahcarried                    -0.0087794  0.0002006  -43.76  < 2e-16 ***
## asahcarried                    -0.0167820  0.0001979  -84.82  < 2e-16 ***
## URBinary:c_daterelevafter SaH  -0.3972812  0.0231576  -17.16  < 2e-16 ***
## URBinary:c_daterelevduring SaH -0.3212096  0.0215502  -14.91  < 2e-16 ***
## URBinary:Date2                  0.0024167  0.0003101    7.79 6.53e-15 ***
## URBinary:dsahcarried           -0.0014551  0.0003835   -3.79 0.000148 ***
## URBinary:asahcarried           -0.0017310  0.0003994   -4.33 1.46e-05 ***
## ---
## Signif. codes:  0 '***' 0.001 '**' 0.01 '*' 0.05 '.' 0.1 ' ' 1
##
## Zero-inflation model:
##                                Estimate Std. Error z value Pr(>|z|)
## (Intercept)                    -1.54573    0.02600  -59.45   <2e-16 ***
## URBinary                        1.05664    0.04693   22.52   <2e-16 ***
## c_daterelevafter SaH           -1.62265    0.03622  -44.80   <2e-16 ***
## c_daterelevduring SaH           0.40845    0.02847   14.35   <2e-16 ***
## URBinary:c_daterelevafter SaH  -1.23150    0.06411  -19.21   <2e-16 ***
## URBinary:c_daterelevduring SaH -1.15857    0.05027  -23.05   <2e-16 ***
## ---
## Signif. codes:  0 '***' 0.001 '**' 0.01 '*' 0.05 '.' 0.1 ' ' 1
# GLMMTMB negative binominal (quadratic version)
summary(lm3glmmrelev)
##  Family: nbinom2  ( log )
## Formula:
## newcase_nst_14 ~ offset(popoff) + URBinary * c_daterelev + URBinary *
##     Date2 + URBinary * dsahcarried + URBinary * asahcarried +      (1 | c_FIPS)
## Data: df_14
##
##       AIC       BIC    logLik  deviance  df.resid
## 1263684.5 1263838.7 -631828.3 1263656.5    446150
##
## Random effects:
##
## Conditional model:
##  Groups Name        Variance Std.Dev.
##  c_FIPS (Intercept) 1.441    1.2
## Number of obs: 446164, groups:  c_FIPS, 3142
##
## Overdispersion parameter for nbinom2 family (): 2.32
##
## Conditional model:
##                                  Estimate Std. Error z value Pr(>|z|)
## (Intercept)                    -2.2865651  0.0301257  -75.90  < 2e-16 ***
## URBinary                       -2.3255120  0.0522028  -44.55  < 2e-16 ***
## c_daterelevafter SaH            0.7262898  0.0144893   50.13  < 2e-16 ***
## c_daterelevduring SaH           0.4488001  0.0113678   39.48  < 2e-16 ***
## Date2                           0.0515785  0.0001851  278.64  < 2e-16 ***
## dsahcarried                    -0.0106564  0.0002756  -38.67  < 2e-16 ***
## asahcarried                    -0.0234832  0.0003504  -67.01  < 2e-16 ***
## URBinary:c_daterelevafter SaH  -0.1897258  0.0234758   -8.08 6.38e-16 ***
## URBinary:c_daterelevduring SaH -0.1100922  0.0185568   -5.93 2.98e-09 ***
## URBinary:Date2                  0.0087959  0.0004420   19.90  < 2e-16 ***
## URBinary:dsahcarried           -0.0081327  0.0005350  -15.20  < 2e-16 ***
## URBinary:asahcarried           -0.0084121  0.0006672  -12.61  < 2e-16 ***
## ---
## Signif. codes:  0 '***' 0.001 '**' 0.01 '*' 0.05 '.' 0.1 ' ' 1
# GLMMTMB negative binomial randomized slope
summary(lm3glmmRandslope)
##  Family: nbinom2  ( log )
## Formula:
## newcase_nst_14 ~ offset(popoff) + URBinary * c_daterelev + URBinary *
##     Date2 + URBinary * dsahcarried + URBinary * asahcarried +
##     (1 + c_daterelev | c_FIPS)
## Data: df_14
##
##      AIC      BIC   logLik deviance df.resid
##       NA       NA       NA       NA   446145
##
## Random effects:
##
## Conditional model:
##  Groups Name                  Variance Std.Dev. Corr
##  c_FIPS (Intercept)           1.483381 1.21794
##         c_daterelevafter SaH  0.003989 0.06316   0.47
##         c_daterelevduring SaH 0.014580 0.12075  -0.50 -1.00
## Number of obs: 446164, groups:  c_FIPS, 3142
##
## Overdispersion parameter for nbinom2 family (): 2.35
##
## Conditional model:
##                                  Estimate Std. Error z value Pr(>|z|)
## (Intercept)                    -2.2863465  0.0304675  -75.04  < 2e-16 ***
## URBinary                       -2.3249281  0.0527023  -44.11  < 2e-16 ***
## c_daterelevafter SaH            0.7184377  0.0146825   48.93  < 2e-16 ***
## c_daterelevduring SaH           0.4167267  0.0118071   35.29  < 2e-16 ***
## Date2                           0.0515031  0.0001844  279.24  < 2e-16 ***
## dsahcarried                    -0.0101321  0.0002778  -36.48  < 2e-16 ***
## asahcarried                    -0.0249385  0.0003593  -69.42  < 2e-16 ***
## URBinary:c_daterelevafter SaH  -0.1864662  0.0236962   -7.87 3.57e-15 ***
## URBinary:c_daterelevduring SaH -0.0970621  0.0190916   -5.08 3.70e-07 ***
## URBinary:Date2                  0.0087402  0.0004402   19.86  < 2e-16 ***
## URBinary:dsahcarried           -0.0081970  0.0005345  -15.34  < 2e-16 ***
## URBinary:asahcarried           -0.0086739  0.0006698  -12.95  < 2e-16 ***
## ---
## Signif. codes:  0 '***' 0.001 '**' 0.01 '*' 0.05 '.' 0.1 ' ' 1
# zero inflated (based on dates) negative binomial mixed effects
summary(lm4catziprelev)
##  Family: nbinom2  ( log )
## Formula:
## newcase_nst_14 ~ offset(popoff) + URBinary * c_daterelev + URBinary *
##     Date2 + URBinary * dsahcarried + URBinary * asahcarried +      (1 | c_FIPS)
## Zero inflation:                  ~URBinary * c_daterelev
## Data: df_14
##
##       AIC       BIC    logLik  deviance  df.resid
## 1260822.9 1261043.1 -630391.5 1260782.9    446144
##
## Random effects:
##
## Conditional model:
##  Groups Name        Variance Std.Dev.
##  c_FIPS (Intercept) 1.447    1.203
## Number of obs: 446164, groups:  c_FIPS, 3142
##
## Overdispersion parameter for nbinom2 family (): 2.76
##
## Conditional model:
##                                  Estimate Std. Error z value Pr(>|z|)
## (Intercept)                    -2.1838911  0.0305875  -71.40   <2e-16 ***
## URBinary                       -2.1493883  0.0543940  -39.52   <2e-16 ***
## c_daterelevafter SaH            0.7968045  0.0147115   54.16   <2e-16 ***
## c_daterelevduring SaH           0.6050528  0.0122626   49.34   <2e-16 ***
## Date2                           0.0504640  0.0001846  273.37   <2e-16 ***
## dsahcarried                    -0.0117809  0.0002795  -42.15   <2e-16 ***
## asahcarried                    -0.0229044  0.0003348  -68.40   <2e-16 ***
## URBinary:c_daterelevafter SaH  -0.2761372  0.0251613  -10.97   <2e-16 ***
## URBinary:c_daterelevduring SaH -0.2094035  0.0213975   -9.79   <2e-16 ***
## URBinary:Date2                  0.0072946  0.0004431   16.46   <2e-16 ***
## URBinary:dsahcarried           -0.0064221  0.0005421  -11.85   <2e-16 ***
## URBinary:asahcarried           -0.0069718  0.0006461  -10.79   <2e-16 ***
## ---
## Signif. codes:  0 '***' 0.001 '**' 0.01 '*' 0.05 '.' 0.1 ' ' 1
##
## Zero-inflation model:
##                                Estimate Std. Error z value Pr(>|z|)
## (Intercept)                     -3.5001     0.1097  -31.91   <2e-16 ***
## URBinary                         1.5671     0.1359   11.53   <2e-16 ***
## c_daterelevafter SaH           -16.0625   185.3833   -0.09    0.931
## c_daterelevduring SaH            1.3823     0.1114   12.41   <2e-16 ***
## URBinary:c_daterelevafter SaH   -1.4518   260.6274   -0.01    0.996
## URBinary:c_daterelevduring SaH  -1.7207     0.1422  -12.10   <2e-16 ***
## ---
## Signif. codes:  0 '***' 0.001 '**' 0.01 '*' 0.05 '.' 0.1 ' ' 1
# #############################################################################
# ############### DISPERSION, RESIDUALS, AND ZERO-INFLATION ###################
# #############################################################################
#
#

SimOut_lm1glmmrelev <- simulateResiduals(fittedModel = lm1glmmrelev, plot = T)
## DHARMa:plot used testOutliers with type = binomial for computational reasons (nObs > 500). Note that this method may not have inflated Type I error rates for integer-valued distributions. To get a more exact result, it is recommended to re-run testOutliers with type = 'bootstrap'. See ?testOutliers for details
plot(SimOut_lm1glmmrelev)
## DHARMa:plot used testOutliers with type = binomial for computational reasons (nObs > 500). Note that this method may not have inflated Type I error rates for integer-valued distributions. To get a more exact result, it is recommended to re-run testOutliers with type = 'bootstrap'. See ?testOutliers for details
```

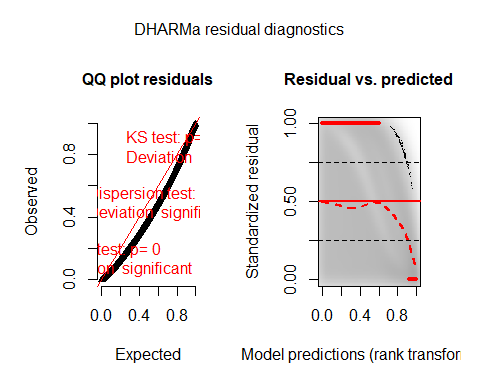

testZeroInflation(SimOut_lm1glmmrelev)

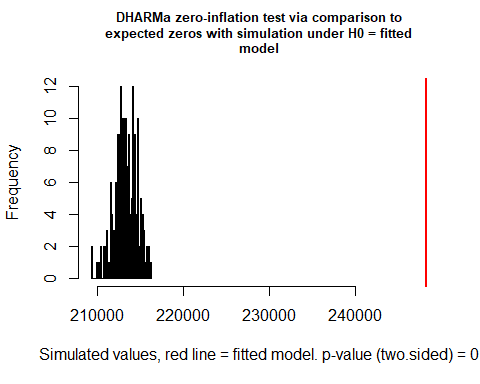

```
##
##  DHARMa zero-inflation test via comparison to expected zeros with
##  simulation under H0 = fitted model
##
## data:  simulationOutput
## ratioObsSim = 1.1636, p-value < 2.2e-16
## alternative hypothesis: two.sided
testOutliers(SimOut_lm1glmmrelev, type= 'bootstrap')
```

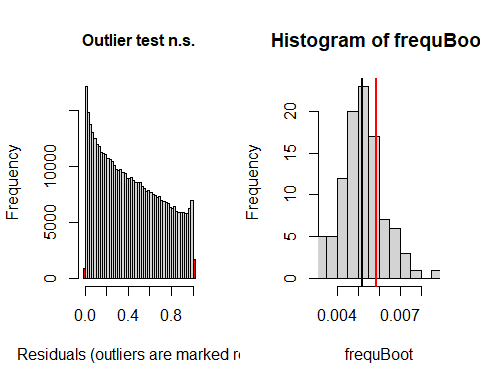

```
##
##  DHARMa bootstrapped outlier test
##
## data:  SimOut_lm1glmmrelev
## outliers at both margin(s) = 2632, observations = 446164, p-value = 0.4
## alternative hypothesis: two.sided
##  percent confidence interval:
##  0.003433379 0.007274399
## sample estimates:
## outlier frequency (expected: 0.00523004993679454 )
##                                        0.005899176
simoutrecalc <- recalculateResiduals(SimOut_lm1glmmrelev, group = df_14$Date2)
testTemporalAutocorrelation(simoutrecalc, time = unique(df_14$Date2))
```

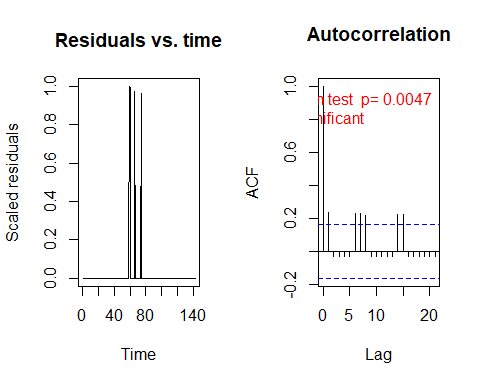

```
##
##  Durbin-Watson test
##
## data:  simulationOutput$scaledResiduals ~ 1
## DW = 1.5288, p-value = 0.004697
## alternative hypothesis: true autocorrelation is not 0
```

\#

SimOut_lm2relev <- simulateResiduals(fittedModel = lm2relev, plot = T)

\## DHARMa:plot used testOutliers with type = binomial for computational reasons (nObs > 500). Note that this method may not have inflated Type I error rates for integer-valued distributions. To get a more exact result, it is recommended to re-run testOutliers with type = 'bootstrap'. See ?testOutliers for details

plot(SimOut_lm2relev)

\## DHARMa:plot used testOutliers with type = binomial for computational reasons (nObs > 500). Note that this method may not have inflated Type I error rates for integer-valued distributions. To get a more exact result, it is recommended to re-run testOutliers with type = 'bootstrap'. See ?testOutliers for details

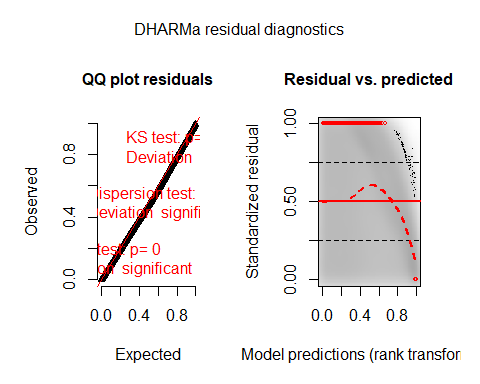

testZeroInflation(SimOut_lm2relev)

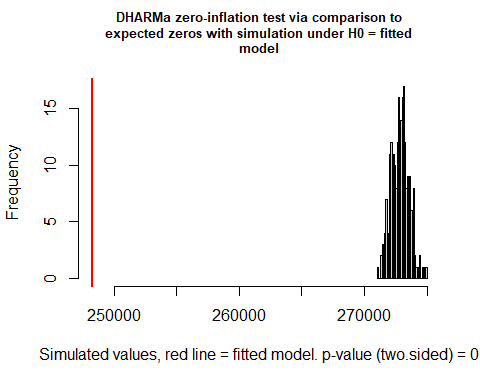

```
##
##  DHARMa zero-inflation test via comparison to expected zeros with
##  simulation under H0 = fitted model
##
## data:  simulationOutput
## ratioObsSim = 0.90973, p-value < 2.2e-16
## alternative hypothesis: two.sided
testOutliers(SimOut_lm2relev, type= 'bootstrap')
```

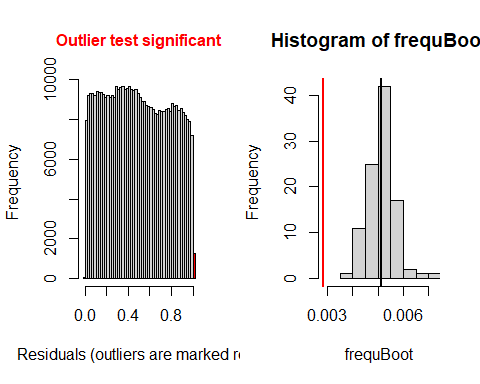

```
##
##  DHARMa bootstrapped outlier test
##
## data:  SimOut_lm2relev
## outliers at both margin(s) = 1262, observations = 446164, p-value <
## 2.2e-16
## alternative hypothesis: two.sided
##  percent confidence interval:
##  0.004220645 0.006071366
## sample estimates:
## outlier frequency (expected: 0.00513037806725778 )
##                                        0.002828556
simoutrecalc <- recalculateResiduals(SimOut_lm2relev, group = df_14$Date2)
testTemporalAutocorrelation(simoutrecalc, time = unique(df_14$Date2))
```

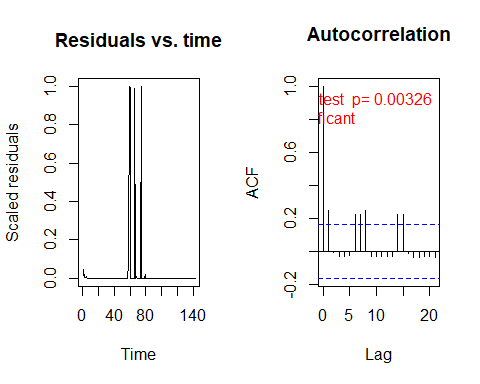

```
##
##  Durbin-Watson test
##
## data:  simulationOutput$scaledResiduals ~ 1
## DW = 1.5096, p-value = 0.003256
## alternative hypothesis: true autocorrelation is not 0
```

\#

SimOut_lm2catziprelev <- simulateResiduals(fittedModel = lm2catziprelev, plot = T)

\## DHARMa:plot used testOutliers with type = binomial for computational reasons (nObs > 500). Note that this method may not have inflated Type I error rates for integer-valued distributions. To get a more exact result, it is recommended to re-run testOutliers with type = 'bootstrap'. See ?testOutliers for details

plot(SimOut_lm2catziprelev)

\## DHARMa:plot used testOutliers with type = binomial for computational reasons (nObs > 500). Note that this method may not have inflated Type I error rates for integer-valued distributions. To get a more exact result, it is recommended to re-run testOutliers with type = 'bootstrap'. See ?testOutliers for details

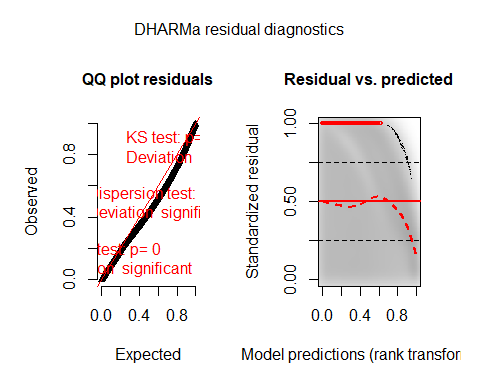

testZeroInflation(SimOut_lm2catziprelev)

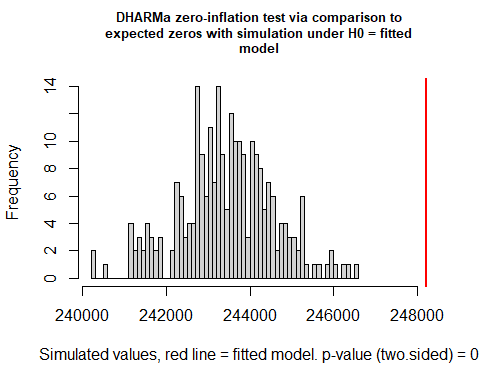

```
##
##  DHARMa zero-inflation test via comparison to expected zeros with
##  simulation under H0 = fitted model
##
## data:  simulationOutput
## ratioObsSim = 1.0196, p-value < 2.2e-16
## alternative hypothesis: two.sided
testOutliers(SimOut_lm2catziprelev, type= 'bootstrap')
```

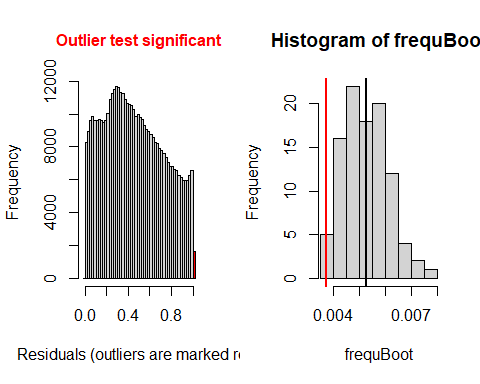

```
##
##  DHARMa bootstrapped outlier test
##
## data:  SimOut_lm2catziprelev
## outliers at both margin(s) = 1660, observations = 446164, p-value =
## 0.04
## alternative hypothesis: two.sided
##  percent confidence interval:
##  0.003838611 0.007088649
## sample estimates:
## outlier frequency (expected: 0.0052525080463686 )
##                                       0.003720605
simoutrecalc <- recalculateResiduals(SimOut_lm2catziprelev, group = df_14$Date2)
testTemporalAutocorrelation(simoutrecalc, time = unique(df_14$Date2))
```

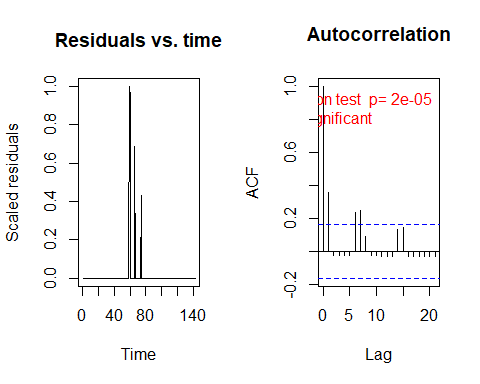

```
##
##  Durbin-Watson test
##
## data:  simulationOutput$scaledResiduals ~ 1
## DW = 1.2925, p-value = 2.181e-05
## alternative hypothesis: true autocorrelation is not 0
```

\###
SimOut_lm3glmmrelev <- simulateResiduals(fittedModel = lm3glmmrelev, plot = T)

\## DHARMa:plot used testOutliers with type = binomial for computational reasons (nObs > 500). Note that this method may not have inflated Type I error rates for integer-valued distributions. To get a more exact result, it is recommended to re-run testOutliers with type = 'bootstrap'. See ?testOutliers for details

plot(SimOut_lm3glmmrelev)

\## DHARMa:plot used testOutliers with type = binomial for computational reasons (nObs > 500). Note that this method may not have inflated Type I error rates for integer-valued distributions. To get a more exact result, it is recommended to re-run testOutliers with type = 'bootstrap'. See ?testOutliers for details

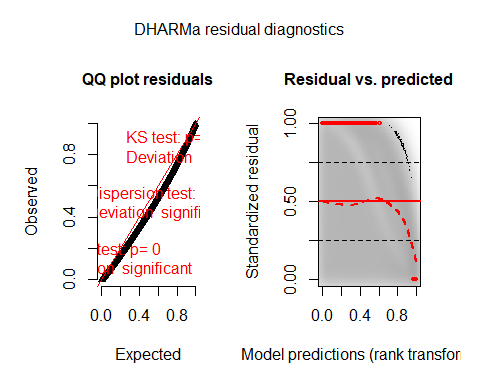

testZeroInflation(SimOut_lm3glmmrelev)

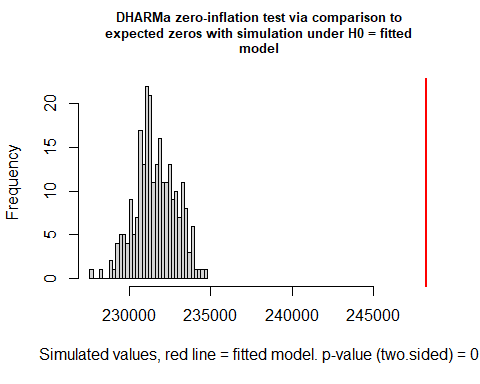

```
##
##  DHARMa zero-inflation test via comparison to expected zeros with
##  simulation under H0 = fitted model
##
## data:  simulationOutput
## ratioObsSim = 1.0716, p-value < 2.2e-16
## alternative hypothesis: two.sided
testOutliers(SimOut_lm3glmmrelev, type= 'bootstrap')
```

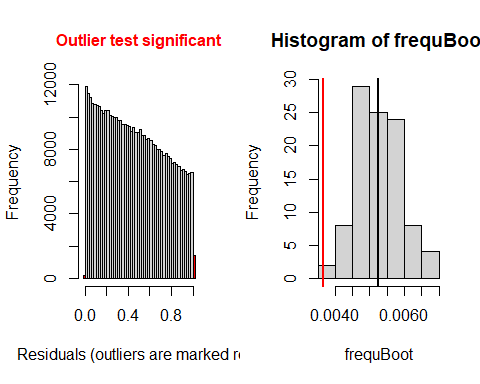

```
##
##  DHARMa bootstrapped outlier test
##
## data:  SimOut_lm3glmmrelev
## outliers at both margin(s) = 1628, observations = 446164, p-value <
## 2.2e-16
## alternative hypothesis: two.sided
##  percent confidence interval:
##  0.004408357 0.006550562
## sample estimates:
## outlier frequency (expected: 0.00526084578764759 )
##                                        0.003648882
simoutrecalc <- recalculateResiduals(SimOut_lm3glmmrelev, group = df_14$Date2)
testTemporalAutocorrelation(simoutrecalc, time = unique(df_14$Date2))
```

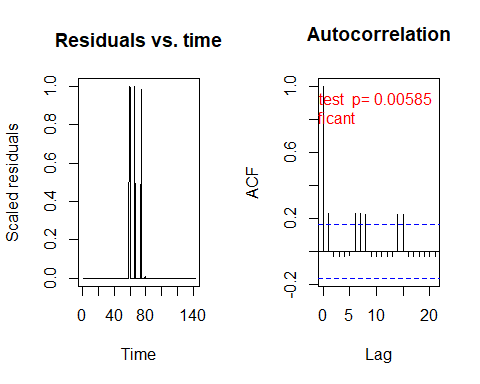

```
##
##  Durbin-Watson test
##
## data:  simulationOutput$scaledResiduals ~ 1
## DW = 1.5407, p-value = 0.005849
## alternative hypothesis: true autocorrelation is not 0
```

SimOut_lm3glmmRandSlope <- simulateResiduals(fittedModel = lm3glmmRandslope, plot = T)

\## DHARMa:plot used testOutliers with type = binomial for computational reasons (nObs > 500). Note that this method may not have inflated Type I error rates for integer-valued distributions. To get a more exact result, it is recommended to re-run testOutliers with type = 'bootstrap'. See ?testOutliers for details

plot(SimOut_lm3glmmRandSlope)

\## DHARMa:plot used testOutliers with type = binomial for computational reasons (nObs > 500). Note that this method may not have inflated Type I error rates for integer-valued distributions. To get a more exact result, it is recommended to re-run testOutliers with type = 'bootstrap'. See ?testOutliers for details

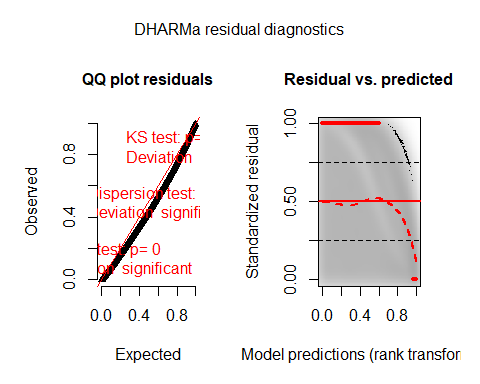

testZeroInflation(SimOut_lm3glmmRandSlope)

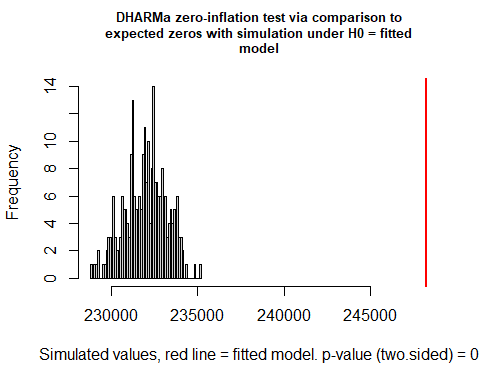

```
##
##  DHARMa zero-inflation test via comparison to expected zeros with
##  simulation under H0 = fitted model
##
## data:  simulationOutput
## ratioObsSim = 1.0701, p-value < 2.2e-16
## alternative hypothesis: two.sided
testOutliers(SimOut_lm3glmmRandSlope, type= 'bootstrap')
```

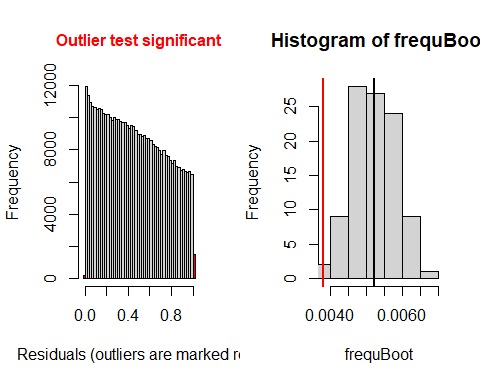

```
##
##  DHARMa bootstrapped outlier test
##
## data:  SimOut_lm3glmmRandSlope
## outliers at both margin(s) = 1690, observations = 446164, p-value <
## 2.2e-16
## alternative hypothesis: two.sided
##  percent confidence interval:
##  0.004183495 0.006223776
## sample estimates:
## outlier frequency (expected: 0.00520227987914758 )
##                                        0.003787845
simoutrecalc <- recalculateResiduals(SimOut_lm3glmmRandSlope, group = df_14$Date2)
testTemporalAutocorrelation(simoutrecalc, time = unique(df_14$Date2))
```

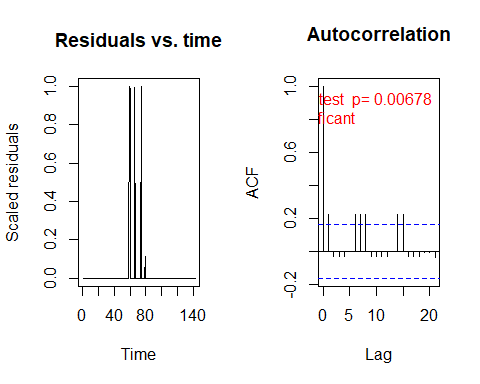

```
##
##  Durbin-Watson test
##
## data:  simulationOutput$scaledResiduals ~ 1
## DW = 1.5488, p-value = 0.006778
## alternative hypothesis: true autocorrelation is not 0
```

\##
SimOut_lm4catziprelev <- simulateResiduals(fittedModel = lm4catziprelev, plot = T)

\## DHARMa:plot used testOutliers with type = binomial for computational reasons (nObs > 500). Note that this method may not have inflated Type I error rates for integer-valued distributions. To get a more exact result, it is recommended to re-run testOutliers with type = 'bootstrap'. See ?testOutliers for details

plot(SimOut_lm4catziprelev)

\## DHARMa:plot used testOutliers with type = binomial for computational reasons (nObs > 500). Note that this method may not have inflated Type I error rates for integer-valued distributions. To get a more exact result, it is recommended to re-run testOutliers with type = 'bootstrap'. See ?testOutliers for details

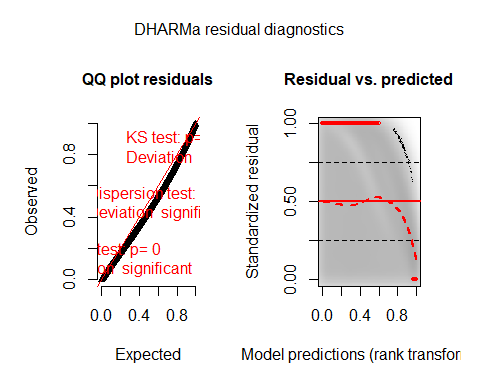

testZeroInflation(SimOut_lm4catziprelev)

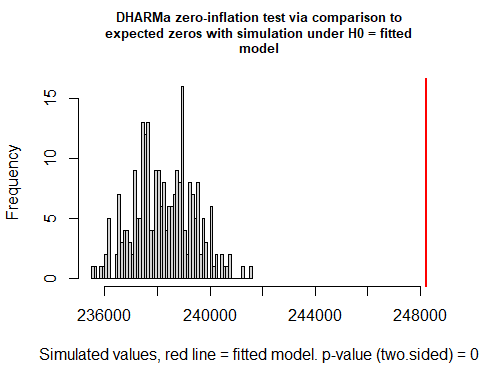

```
##
##  DHARMa zero-inflation test via comparison to expected zeros with
##  simulation under H0 = fitted model
##
## data:  simulationOutput
## ratioObsSim = 1.0419, p-value < 2.2e-16
## alternative hypothesis: two.sided
testOutliers(SimOut_lm4catziprelev, type= 'bootstrap')
```

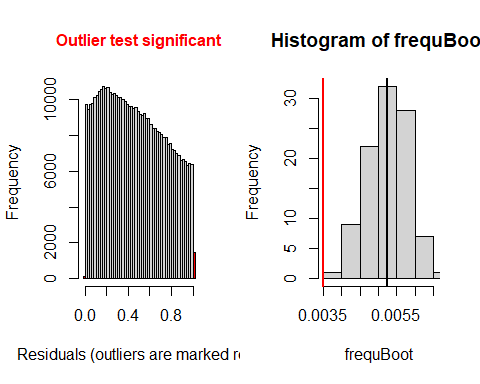

```
##
##  DHARMa bootstrapped outlier test
##
## data:  SimOut_lm4catziprelev
## outliers at both margin(s) = 1557, observations = 446164, p-value <
## 2.2e-16
## alternative hypothesis: two.sided
##  percent confidence interval:
##  0.004233533 0.006230556
## sample estimates:
## outlier frequency (expected: 0.00523818595852646 )
##                                        0.003489748
simoutrecalc <- recalculateResiduals(SimOut_lm4catziprelev, group = df_14$Date2)
testTemporalAutocorrelation(simoutrecalc, time = unique(df_14$Date2))
```

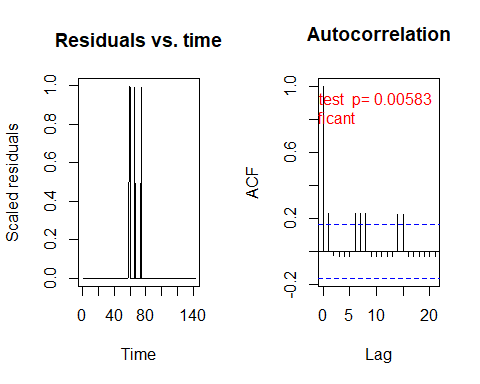

```
##
##  Durbin-Watson test
##
## data:  simulationOutput$scaledResiduals ~ 1
## DW = 1.5405, p-value = 0.005825
## alternative hypothesis: true autocorrelation is not 0
# REMOVING OUTLIERS LM3GLMM
r <- which(residuals(SimOut_lm3glmmrelev) == 1 | residuals(SimOut_lm3glmmrelev) == 0)

df_14$row <- c(1:446164)

`%notin%` <- Negate(`%in%`)
#removing the counties
outcount <- df_14$c_FIPS[df_14$row %in% r]
outcount <- unique(outcount)
df_14outremcount <- df_14[df_14$c_FIPS %notin% outcount,]


# REMOVING OUTLIERS LM3GLMM
summary(lm3glmmrelevoutcount)
##  Family: nbinom2  ( log )
## Formula:
## newcase_nst_14 ~ offset(popoff) + URBinary * c_daterelev + URBinary *
##     Date2 + URBinary * dsahcarried + URBinary * asahcarried +      (1 | c_FIPS)
## Data: df_14outremcount
##
##       AIC       BIC    logLik  deviance  df.resid
## 1122928.3 1123080.8 -561450.1 1122900.3    397018
##
## Random effects:
##
## Conditional model:
##  Groups Name        Variance Std.Dev.
##  c_FIPS (Intercept) 0.8883   0.9425
## Number of obs: 397032, groups:  c_FIPS, 2796
##
## Overdispersion parameter for nbinom2 family (): 2.32
##
## Conditional model:
##                                  Estimate Std. Error z value Pr(>|z|)
## (Intercept)                    -2.4976002  0.0262870  -95.01  < 2e-16 ***
## URBinary                       -2.3869755  0.0486656  -49.05  < 2e-16 ***
## c_daterelevafter SaH            0.7092342  0.0149282   47.51  < 2e-16 ***
## c_daterelevduring SaH           0.4457600  0.0116989   38.10  < 2e-16 ***
## Date2                           0.0521630  0.0002013  259.14  < 2e-16 ***
## dsahcarried                    -0.0111433  0.0002878  -38.71  < 2e-16 ***
## asahcarried                    -0.0239252  0.0003697  -64.71  < 2e-16 ***
## URBinary:c_daterelevafter SaH  -0.2107631  0.0251152   -8.39  < 2e-16 ***
## URBinary:c_daterelevduring SaH -0.1296061  0.0198505   -6.53 6.62e-11 ***
## URBinary:Date2                  0.0098601  0.0005052   19.52  < 2e-16 ***
## URBinary:dsahcarried           -0.0092129  0.0005957  -15.47  < 2e-16 ***
## URBinary:asahcarried           -0.0094811  0.0007393  -12.82  < 2e-16 ***
## ---
## Signif. codes:  0 '***' 0.001 '**' 0.01 '*' 0.05 '.' 0.1 ' ' 1
SimOut_lm3glmmrelevoutcount <- simulateResiduals(fittedModel = lm3glmmrelevoutcount, plot = T)
## DHARMa:plot used testOutliers with type = binomial for computational reasons (nObs > 500). Note that this method may not have inflated Type I error rates for integer-valued distributions. To get a more exact result, it is recommended to re-run testOutliers with type = 'bootstrap'. See ?testOutliers for details
plot(SimOut_lm3glmmrelevoutcount)
## DHARMa:plot used testOutliers with type = binomial for computational reasons (nObs > 500). Note that this method may not have inflated Type I error rates for integer-valued distributions. To get a more exact result, it is recommended to re-run testOutliers with type = 'bootstrap'. See ?testOutliers for details
```

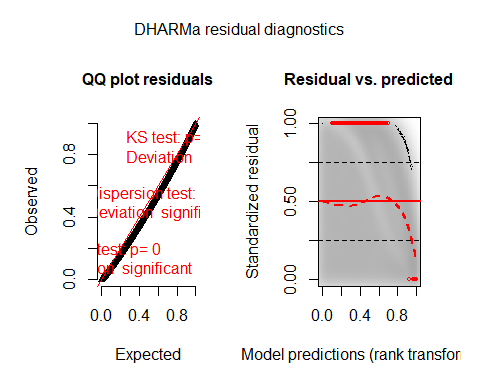

testZeroInflation(SimOut_lm3glmmrelevoutcount)

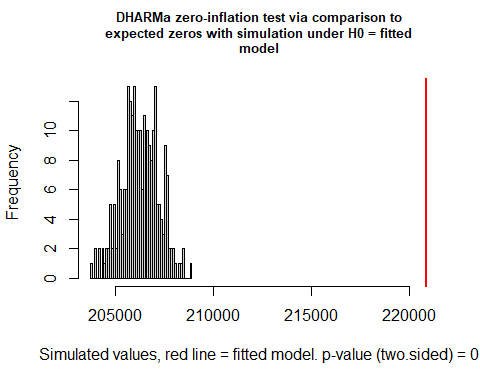

```
##
##  DHARMa zero-inflation test via comparison to expected zeros with
##  simulation under H0 = fitted model
##
## data:  simulationOutput
## ratioObsSim = 1.0709, p-value < 2.2e-16
## alternative hypothesis: two.sided
testOutliers(SimOut_lm3glmmrelevoutcount, type= 'bootstrap')
```

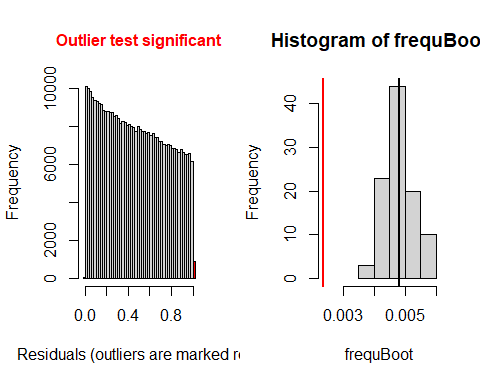

```
##
##  DHARMa bootstrapped outlier test
##
## data:  SimOut_lm3glmmrelevoutcount
## outliers at both margin(s) = 936, observations = 397032, p-value <
## 2.2e-16
## alternative hypothesis: two.sided
##  percent confidence interval:
##  0.003927139 0.005768616
## sample estimates:
## outlier frequency (expected: 0.00480739587741038 )
##                                        0.002357493
simoutrecalc <- recalculateResiduals(SimOut_lm3glmmrelevoutcount, group = df_14outremcount$Date2)
testTemporalAutocorrelation(simoutrecalc, time = unique(df_14outremcount$Date2))
```

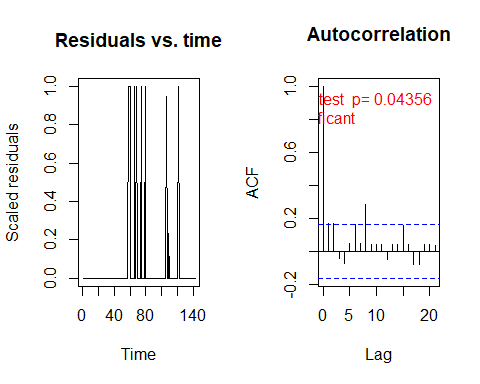

```
##
##  Durbin-Watson test
##
## data:  simulationOutput$scaledResiduals ~ 1
## DW = 1.6636, p-value = 0.04356
## alternative hypothesis: true autocorrelation is not 0
# REMOVING OUTLIERS RANDOM SLOPE
r <- which(residuals(SimOut_lm3glmmRandSlope) == 1 | residuals(SimOut_lm3glmmRandSlope) == 0)

df_14$row <- c(1:446164)

`%notin%` <- Negate(`%in%`)
#removing the counties
outcount <- df_14$c_FIPS[df_14$row %in% r]
outcount <- unique(outcount)
df_14outremcountrand <- df_14[df_14$c_FIPS %notin% outcount,]


# REMOVING OUTLIERS RANDOM SLOPE
summary(lm3glmmrelevoutcountrandslope)
##  Family: nbinom2  ( log )
## Formula:
## newcase_nst_14 ~ offset(popoff) + URBinary * c_daterelev + URBinary *
##     Date2 + URBinary * dsahcarried + URBinary * asahcarried +      (1 | c_FIPS)
## Data: df_14outremcountrand
##
##       AIC       BIC    logLik  deviance  df.resid
## 1116060.2 1116212.6 -558016.1 1116032.2    394604
##
## Random effects:
##
## Conditional model:
##  Groups Name        Variance Std.Dev.
##  c_FIPS (Intercept) 0.8737   0.9347
## Number of obs: 394618, groups:  c_FIPS, 2779
##
## Overdispersion parameter for nbinom2 family (): 2.32
##
## Conditional model:
##                                  Estimate Std. Error z value Pr(>|z|)
## (Intercept)                    -2.5169881  0.0262260  -95.97  < 2e-16 ***
## URBinary                       -2.4038411  0.0485358  -49.53  < 2e-16 ***
## c_daterelevafter SaH            0.7095800  0.0149617   47.43  < 2e-16 ***
## c_daterelevduring SaH           0.4467881  0.0117272   38.10  < 2e-16 ***
## Date2                           0.0521934  0.0002024  257.85  < 2e-16 ***
## dsahcarried                    -0.0111831  0.0002890  -38.69  < 2e-16 ***
## asahcarried                    -0.0239429  0.0003709  -64.56  < 2e-16 ***
## URBinary:c_daterelevafter SaH  -0.2141177  0.0252114   -8.49  < 2e-16 ***
## URBinary:c_daterelevduring SaH -0.1273352  0.0199174   -6.39 1.62e-10 ***
## URBinary:Date2                  0.0097266  0.0005057   19.24  < 2e-16 ***
## URBinary:dsahcarried           -0.0090229  0.0005961  -15.14  < 2e-16 ***
## URBinary:asahcarried           -0.0092761  0.0007424  -12.49  < 2e-16 ***
## ---
## Signif. codes:  0 '***' 0.001 '**' 0.01 '*' 0.05 '.' 0.1 ' ' 1
SimOut_lm3glmmrelevoutcountrandslope <- simulateResiduals(fittedModel = lm3glmmrelevoutcountrandslope, plot = T)
## DHARMa:plot used testOutliers with type = binomial for computational reasons (nObs > 500). Note that this method may not have inflated Type I error rates for integer-valued distributions. To get a more exact result, it is recommended to re-run testOutliers with type = 'bootstrap'. See ?testOutliers for details
plot(SimOut_lm3glmmrelevoutcountrandslope)
## DHARMa:plot used testOutliers with type = binomial for computational reasons (nObs > 500). Note that this method may not have inflated Type I error rates for integer-valued distributions. To get a more exact result, it is recommended to re-run testOutliers with type = 'bootstrap'. See ?testOutliers for details
```

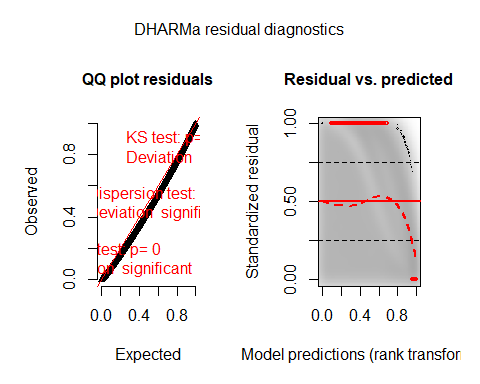

testZeroInflation(SimOut_lm3glmmrelevoutcountrandslope)

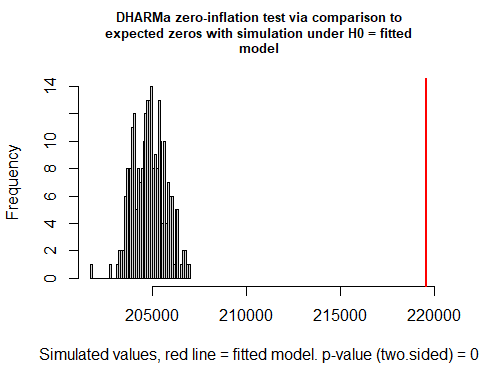

```
##
##  DHARMa zero-inflation test via comparison to expected zeros with
##  simulation under H0 = fitted model
##
## data:  simulationOutput
## ratioObsSim = 1.0717, p-value < 2.2e-16
## alternative hypothesis: two.sided
testOutliers(SimOut_lm3glmmrelevoutcountrandslope, type= 'bootstrap')
```

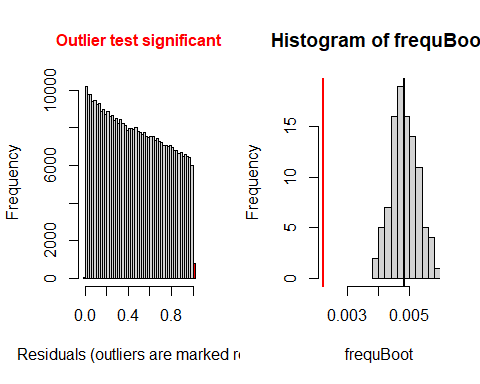

```
##
##  DHARMa bootstrapped outlier test
##
## data:  SimOut_lm3glmmrelevoutcountrandslope
## outliers at both margin(s) = 873, observations = 394618, p-value <
## 2.2e-16
## alternative hypothesis: two.sided
##  percent confidence interval:
##  0.004049106 0.005680937
## sample estimates:
## outlier frequency (expected: 0.00483505567409495 )
##                                        0.002212266
simoutrecalc <- recalculateResiduals(SimOut_lm3glmmrelevoutcountrandslope, group = df_14outremcountrand$Date2)
testTemporalAutocorrelation(simoutrecalc, time = unique(df_14outremcountrand$Date2))
```

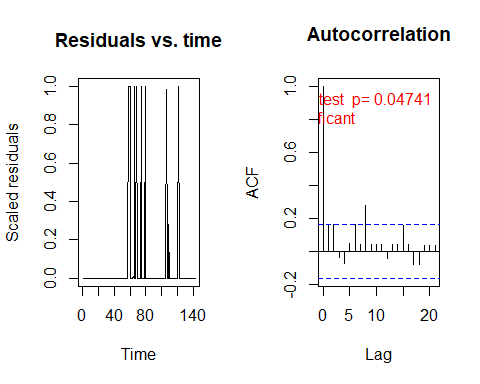

```
##
##  Durbin-Watson test
##
## data:  simulationOutput$scaledResiduals ~ 1
## DW = 1.6696, p-value = 0.04741
## alternative hypothesis: true autocorrelation is not 0
df_sah <- data.frame(cbind(df_14$FIPS, df_14$Date2, df_14$URBinary, df_14$daysSaH))
df_sahs <- df_sah[df_sah$X2 == 1,]
colnames(df_sahs) <- c("FIPS", "Date", "County_Type", "Days_under_SAH")
df_sahs$County_Type <- factor(df_sahs$County_Type, levels = c(0,1), labels =c("Rural", "Urban"))
wilcoxon <- wilcox.test(Days_under_SAH ~ County_Type,data= df_sahs)
wilcoxon
##
##  Wilcoxon rank sum test with continuity correction
##
## data:  Days_under_SAH by County_Type
## W = 897959, p-value < 2.2e-16
## alternative hypothesis: true location shift is not equal to 0
```

#### Ten-Day Lag

```
#reads in data
setwd("C:\\Users\\Jake\\Desktop\\MAYO\\COVID RURALITY")
df_14 <- read.csv("df_14.csv",header=T)

#installs packages then loads them into the session
library(glmmTMB)
## Warning: package 'glmmTMB' was built under R version 3.6.3
library(DHARMa)
## Warning: package 'DHARMa' was built under R version 3.6.3
## This is DHARMa 0.3.3.0. For overview type '?DHARMa'. For recent changes, type news(package = 'DHARMa') Note: Syntax of plotResiduals has changed in 0.3.0, see ?plotResiduals for details
# Releveling
df_14$c_daterelev <- relevel(df_14$c_date, ref = "before SaH")


#ten Day Lag
n <- 142
D <- 10
for (i in 1:n){
  df_14$newcase_nst_10[df_14$Date2 == i] <- ifelse( i > (n-D), df_14$newcase_nst_14[df_14$Date2 == (i-(14-D))], df_14$newcase_nst[df_14$Date2 == (i+D)])
}

#RENAMING THE VARIABLE TO ALLOW the implementation of the lag

df_14$newcase_nst_14 <- df_14$newcase_nst_10


load("C:/Users/Jake/Desktop/MAYO/COVID RURALITY/10day.RData")
###########################################
############# SUMMARY RESULTS #############
###########################################

# GLMMTMB mixed effects poisson model
summary(lm1glmmrelev)
##  Family: poisson  ( log )
## Formula:
## newcase_nst_14 ~ offset(popoff) + URBinary * c_daterelev + URBinary *
##     Date2 + URBinary * dsahcarried + URBinary * asahcarried +      (1 | c_FIPS)
## Data: df_14
##
##       AIC       BIC    logLik  deviance  df.resid
## 1625845.6 1625988.7 -812909.8 1625819.6    446151
##
## Random effects:
##
## Conditional model:
##  Groups Name        Variance Std.Dev.
##  c_FIPS (Intercept) 1.533    1.238
## Number of obs: 446164, groups:  c_FIPS, 3142
##
## Conditional model:
##                                  Estimate Std. Error z value Pr(>|z|)
## (Intercept)                    -1.305e+00  2.904e-02   -44.9  < 2e-16 ***
## URBinary                       -2.131e+00  4.847e-02   -44.0  < 2e-16 ***
## c_daterelevafter SaH            7.029e-01  9.413e-03    74.7  < 2e-16 ***
## c_daterelevduring SaH           5.687e-01  8.149e-03    69.8  < 2e-16 ***
## Date2                           4.148e-02  9.992e-05   415.2  < 2e-16 ***
## dsahcarried                    -3.916e-03  1.567e-04   -25.0  < 2e-16 ***
## asahcarried                    -1.561e-02  1.699e-04   -91.8  < 2e-16 ***
## URBinary:c_daterelevafter SaH  -1.193e-01  1.519e-02    -7.9 3.99e-15 ***
## URBinary:c_daterelevduring SaH -3.804e-02  1.325e-02    -2.9  0.00408 **
## URBinary:Date2                  4.453e-03  2.382e-04    18.7  < 2e-16 ***
## URBinary:dsahcarried           -4.388e-03  2.968e-04   -14.8  < 2e-16 ***
## URBinary:asahcarried           -3.570e-03  3.312e-04   -10.8  < 2e-16 ***
## ---
## Signif. codes:  0 '***' 0.001 '**' 0.01 '*' 0.05 '.' 0.1 ' ' 1
# Zero inflated poisson mixed effects (zero inflated using the whole formula)
summary(lm2relev)
##  Family: poisson  ( log )
## Formula:
## newcase_nst_14 ~ offset(popoff) + URBinary * c_daterelev + URBinary *
##     Date2 + URBinary * dsahcarried + URBinary * asahcarried +      (1 | c_FIPS)
## Zero inflation:
## ~URBinary * c_daterelev + URBinary * Date2 + URBinary * dsahcarried +
##     URBinary * asahcarried
## Data: df_14
##
##       AIC       BIC    logLik  deviance  df.resid
## 1490527.7 1490802.9 -745238.8 1490477.7    446139
##
## Random effects:
##
## Conditional model:
##  Groups Name        Variance Std.Dev.
##  c_FIPS (Intercept) 1.439    1.2
## Number of obs: 446164, groups:  c_FIPS, 3142
##
## Conditional model:
##                                  Estimate Std. Error z value Pr(>|z|)
## (Intercept)                     0.9012932  0.0291932   30.87  < 2e-16 ***
## URBinary                       -1.8854025  0.0495544  -38.05  < 2e-16 ***
## c_daterelevafter SaH           -0.0602801  0.0104942   -5.74 9.24e-09 ***
## c_daterelevduring SaH          -0.2367721  0.0092925  -25.48  < 2e-16 ***
## Date2                           0.0268085  0.0001248  214.79  < 2e-16 ***
## dsahcarried                     0.0005851  0.0001823    3.21  0.00133 **
## asahcarried                    -0.0063314  0.0001880  -33.68  < 2e-16 ***
## URBinary:c_daterelevafter SaH  -0.0540179  0.0167620   -3.22  0.00127 **
## URBinary:c_daterelevduring SaH  0.0185206  0.0149098    1.24  0.21417
## URBinary:Date2                  0.0009268  0.0002920    3.17  0.00151 **
## URBinary:dsahcarried           -0.0005445  0.0003502   -1.55  0.11997
## URBinary:asahcarried           -0.0001513  0.0003754   -0.40  0.68699
## ---
## Signif. codes:  0 '***' 0.001 '**' 0.01 '*' 0.05 '.' 0.1 ' ' 1
##
## Zero-inflation model:
##                                  Estimate Std. Error z value Pr(>|z|)
## (Intercept)                     5.6303031  0.0327253  172.05  < 2e-16 ***
## URBinary                        0.2101251  0.0582938    3.60 0.000313 ***
## c_daterelevafter SaH            1.6899759  0.0503264   33.58  < 2e-16 ***
## c_daterelevduring SaH           0.2283812  0.0209249   10.91  < 2e-16 ***
## Date2                          -0.0809809  0.0005073 -159.64  < 2e-16 ***
## dsahcarried                     0.0023634  0.0007779    3.04 0.002380 **
## asahcarried                    -0.1238826  0.0051268  -24.16  < 2e-16 ***
## URBinary:c_daterelevafter SaH   0.3146067  0.0853122    3.69 0.000226 ***
## URBinary:c_daterelevduring SaH  0.0852503  0.0350038    2.44 0.014873 *
## URBinary:Date2                 -0.0046190  0.0009543   -4.84  1.3e-06 ***
## URBinary:dsahcarried            0.0043723  0.0012304    3.55 0.000380 ***
## URBinary:asahcarried           -0.0363792  0.0096597   -3.77 0.000166 ***
## ---
## Signif. codes:  0 '***' 0.001 '**' 0.01 '*' 0.05 '.' 0.1 ' ' 1
# Zero inflated poisson mixed effects (zero inflated using the rurality and dates)
summary(lm2catziprelev)
##  Family: poisson  ( log )
## Formula:
## newcase_nst_14 ~ offset(popoff) + URBinary * c_daterelev + URBinary *
##     Date2 + URBinary * dsahcarried + URBinary * asahcarried +      (1 | c_FIPS)
## Zero inflation:                  ~URBinary * c_daterelev
## Data: df_14
##
##       AIC       BIC    logLik  deviance  df.resid
## 1580358.4 1580567.6 -790160.2 1580320.4    446145
##
## Random effects:
##
## Conditional model:
##  Groups Name        Variance Std.Dev.
##  c_FIPS (Intercept) 1.526    1.236
## Number of obs: 446164, groups:  c_FIPS, 3142
##
## Conditional model:
##                                  Estimate Std. Error z value Pr(>|z|)
## (Intercept)                    -0.8388483  0.0300594   -27.9  < 2e-16 ***
## URBinary                       -1.5289178  0.0517319   -29.6  < 2e-16 ***
## c_daterelevafter SaH            0.8518576  0.0111095    76.7  < 2e-16 ***
## c_daterelevduring SaH           0.7121948  0.0099498    71.6  < 2e-16 ***
## Date2                           0.0377610  0.0001168   323.2  < 2e-16 ***
## dsahcarried                    -0.0059850  0.0001798   -33.3  < 2e-16 ***
## asahcarried                    -0.0165647  0.0001840   -90.0  < 2e-16 ***
## URBinary:c_daterelevafter SaH  -0.5690112  0.0200518   -28.4  < 2e-16 ***
## URBinary:c_daterelevduring SaH -0.4933506  0.0184905   -26.7  < 2e-16 ***
## URBinary:Date2                  0.0016374  0.0002762     5.9 3.07e-09 ***
## URBinary:dsahcarried           -0.0013485  0.0003407    -4.0 7.56e-05 ***
## URBinary:asahcarried           -0.0007427  0.0003652    -2.0    0.042 *
## ---
## Signif. codes:  0 '***' 0.001 '**' 0.01 '*' 0.05 '.' 0.1 ' ' 1
##
## Zero-inflation model:
##                                Estimate Std. Error z value Pr(>|z|)
## (Intercept)                    -1.25889    0.02310  -54.50  < 2e-16 ***
## URBinary                        1.34931    0.03367   40.08  < 2e-16 ***
## c_daterelevafter SaH           -2.00159    0.03468  -57.71  < 2e-16 ***
## c_daterelevduring SaH          -0.18711    0.02623   -7.13 9.84e-13 ***
## URBinary:c_daterelevafter SaH  -1.47527    0.05563  -26.52  < 2e-16 ***
## URBinary:c_daterelevduring SaH -1.43643    0.03877  -37.05  < 2e-16 ***
## ---
## Signif. codes:  0 '***' 0.001 '**' 0.01 '*' 0.05 '.' 0.1 ' ' 1
# GLMMTMB negative binominal (quadratic version)
summary(lm3glmmrelev)
##  Family: nbinom2  ( log )
## Formula:
## newcase_nst_14 ~ offset(popoff) + URBinary * c_daterelev + URBinary *
##     Date2 + URBinary * dsahcarried + URBinary * asahcarried +      (1 | c_FIPS)
## Data: df_14
##
##      AIC      BIC   logLik deviance df.resid
##  1379454  1379608  -689713  1379426   446150
##
## Random effects:
##
## Conditional model:
##  Groups Name        Variance Std.Dev.
##  c_FIPS (Intercept) 1.442    1.201
## Number of obs: 446164, groups:  c_FIPS, 3142
##
## Overdispersion parameter for nbinom2 family (): 2.11
##
## Conditional model:
##                                  Estimate Std. Error z value Pr(>|z|)
## (Intercept)                    -1.8557621  0.0295613  -62.78  < 2e-16 ***
## URBinary                       -2.3146332  0.0507335  -45.62  < 2e-16 ***
## c_daterelevafter SaH            0.5048157  0.0135887   37.15  < 2e-16 ***
## c_daterelevduring SaH           0.3117852  0.0102445   30.43  < 2e-16 ***
## Date2                           0.0501722  0.0001733  289.53  < 2e-16 ***
## dsahcarried                    -0.0089371  0.0002640  -33.85  < 2e-16 ***
## asahcarried                    -0.0243228  0.0003475  -69.99  < 2e-16 ***
## URBinary:c_daterelevafter SaH  -0.1864333  0.0220662   -8.45  < 2e-16 ***
## URBinary:c_daterelevduring SaH -0.1003363  0.0167657   -5.98 2.17e-09 ***
## URBinary:Date2                  0.0089095  0.0004070   21.89  < 2e-16 ***
## URBinary:dsahcarried           -0.0089243  0.0005023  -17.77  < 2e-16 ***
## URBinary:asahcarried           -0.0080379  0.0006506  -12.36  < 2e-16 ***
## ---
## Signif. codes:  0 '***' 0.001 '**' 0.01 '*' 0.05 '.' 0.1 ' ' 1
# GLMMTMB negative binomial randomized slope
summary(lm3glmmRandslope)
##  Family: nbinom2  ( log )
## Formula:
## newcase_nst_14 ~ offset(popoff) + URBinary * c_daterelev + URBinary *
##     Date2 + URBinary * dsahcarried + URBinary * asahcarried +
##     (1 + c_daterelev | c_FIPS)
## Data: df_14
##
##      AIC      BIC   logLik deviance df.resid
##       NA       NA       NA       NA   446145
##
## Random effects:
##
## Conditional model:
##  Groups Name                  Variance Std.Dev. Corr
##  c_FIPS (Intercept)           1.403543 1.18471
##         c_daterelevafter SaH  0.006503 0.08064   0.80
##         c_daterelevduring SaH 0.004194 0.06476  -0.17 -0.72
## Number of obs: 446164, groups:  c_FIPS, 3142
##
## Overdispersion parameter for nbinom2 family (): 2.12
##
## Conditional model:
##                                  Estimate Std. Error z value Pr(>|z|)
## (Intercept)                    -1.8561592  0.0296052  -62.70  < 2e-16 ***
## URBinary                       -2.3171274  0.0502043  -46.15  < 2e-16 ***
## c_daterelevafter SaH            0.4971476  0.0139280   35.69  < 2e-16 ***
## c_daterelevduring SaH           0.2923361  0.0105142   27.80  < 2e-16 ***
## Date2                           0.0501534  0.0001733  289.43  < 2e-16 ***
## dsahcarried                    -0.0085366  0.0002666  -32.02  < 2e-16 ***
## asahcarried                    -0.0253004  0.0003572  -70.82  < 2e-16 ***
## URBinary:c_daterelevafter SaH  -0.1850089  0.0226273   -8.18 2.93e-16 ***
## URBinary:c_daterelevduring SaH -0.0927658  0.0170413   -5.44 5.22e-08 ***
## URBinary:Date2                  0.0089269  0.0004063   21.97  < 2e-16 ***
## URBinary:dsahcarried           -0.0090932  0.0005042  -18.03  < 2e-16 ***
## URBinary:asahcarried           -0.0082176  0.0006544  -12.56  < 2e-16 ***
## ---
## Signif. codes:  0 '***' 0.001 '**' 0.01 '*' 0.05 '.' 0.1 ' ' 1
# zero inflated (based on dates) negative binomial mixed effects
summary(lm4catziprelev)
##  Family: nbinom2  ( log )
## Formula:
## newcase_nst_14 ~ offset(popoff) + URBinary * c_daterelev + URBinary *
##     Date2 + URBinary * dsahcarried + URBinary * asahcarried +      (1 | c_FIPS)
## Zero inflation:                  ~URBinary * c_daterelev
## Data: df_14
##
##       AIC       BIC    logLik  deviance  df.resid
## 1378848.8 1379068.9 -689404.4 1378808.8    446144
##
## Random effects:
##
## Conditional model:
##  Groups Name        Variance Std.Dev.
##  c_FIPS (Intercept) 1.442    1.201
## Number of obs: 446164, groups:  c_FIPS, 3142
##
## Overdispersion parameter for nbinom2 family (): 2.22
##
## Conditional model:
##                                  Estimate Std. Error z value Pr(>|z|)
## (Intercept)                    -1.8317151  0.0301027  -60.85   <2e-16 ***
## URBinary                       -2.0121840  0.0533612  -37.71   <2e-16 ***
## c_daterelevafter SaH            0.5302388  0.0138961   38.16   <2e-16 ***
## c_daterelevduring SaH           0.3573428  0.0111221   32.13   <2e-16 ***
## Date2                           0.0498785  0.0001776  280.90   <2e-16 ***
## dsahcarried                    -0.0093296  0.0002690  -34.68   <2e-16 ***
## asahcarried                    -0.0241742  0.0003443  -70.22   <2e-16 ***
## URBinary:c_daterelevafter SaH  -0.3706921  0.0244454  -15.16   <2e-16 ***
## URBinary:c_daterelevduring SaH -0.2899238  0.0203126  -14.27   <2e-16 ***
## URBinary:Date2                  0.0069571  0.0004225   16.47   <2e-16 ***
## URBinary:dsahcarried           -0.0068333  0.0005190  -13.17   <2e-16 ***
## URBinary:asahcarried           -0.0060872  0.0006531   -9.32   <2e-16 ***
## ---
## Signif. codes:  0 '***' 0.001 '**' 0.01 '*' 0.05 '.' 0.1 ' ' 1
##
## Zero-inflation model:
##                                Estimate Std. Error z value Pr(>|z|)
## (Intercept)                     -5.4348     0.7506  -7.240 4.48e-13 ***
## URBinary                         3.9864     0.7508   5.310 1.10e-07 ***
## c_daterelevafter SaH           -14.8661   188.2534  -0.079   0.9371
## c_daterelevduring SaH            1.9216     0.7479   2.569   0.0102 *
## URBinary:c_daterelevafter SaH   -0.3234   191.6502  -0.002   0.9987
## URBinary:c_daterelevduring SaH  -4.2609     0.7649  -5.571 2.54e-08 ***
## ---
## Signif. codes:  0 '***' 0.001 '**' 0.01 '*' 0.05 '.' 0.1 ' ' 1
# #############################################################################
# ############### DISPERSION, RESIDUALS, AND ZERO-INFLATION ###################
# #############################################################################
#
#

SimOut_lm1glmmrelev <- simulateResiduals(fittedModel = lm1glmmrelev, plot = T)
## DHARMa:plot used testOutliers with type = binomial for computational reasons (nObs > 500). Note that this method may not have inflated Type I error rates for integer-valued distributions. To get a more exact result, it is recommended to re-run testOutliers with type = 'bootstrap'. See ?testOutliers for details
plot(SimOut_lm1glmmrelev)
## DHARMa:plot used testOutliers with type = binomial for computational reasons (nObs > 500). Note that this method may not have inflated Type I error rates for integer-valued distributions. To get a more exact result, it is recommended to re-run testOutliers with type = 'bootstrap'. See ?testOutliers for details
```

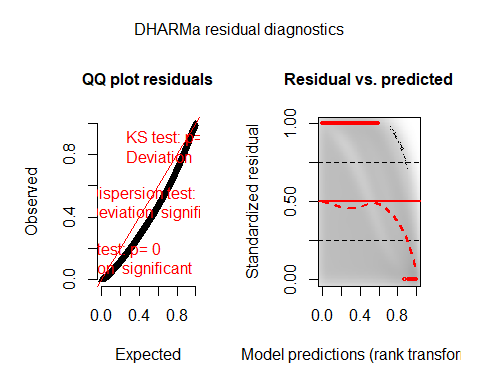

testZeroInflation(SimOut_lm1glmmrelev)

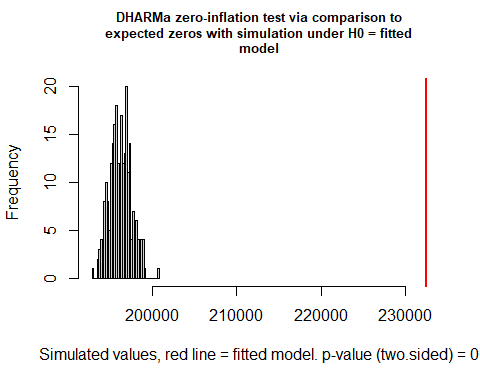

```
##
##  DHARMa zero-inflation test via comparison to expected zeros with
##  simulation under H0 = fitted model
##
## data:  simulationOutput
## ratioObsSim = 1.1853, p-value < 2.2e-16
## alternative hypothesis: two.sided
testOutliers(SimOut_lm1glmmrelev, type= 'bootstrap')
```

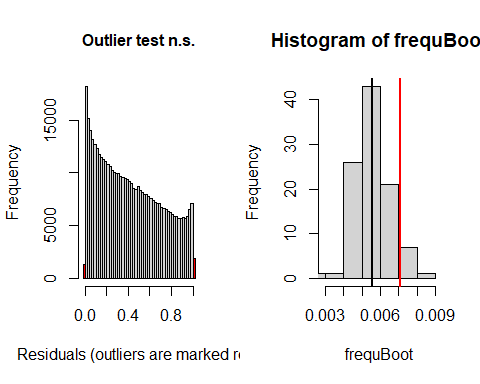

```
##
##  DHARMa bootstrapped outlier test
##
## data:  SimOut_lm1glmmrelev
## outliers at both margin(s) = 3164, observations = 446164, p-value =
## 0.16
## alternative hypothesis: two.sided
##  percent confidence interval:
##  0.004089808 0.007597823
## sample estimates:
## outlier frequency (expected: 0.0055858383912642 )
##                                       0.007091563
simoutrecalc <- recalculateResiduals(SimOut_lm1glmmrelev, group = df_14$Date2)
testTemporalAutocorrelation(simoutrecalc, time = unique(df_14$Date2))
```

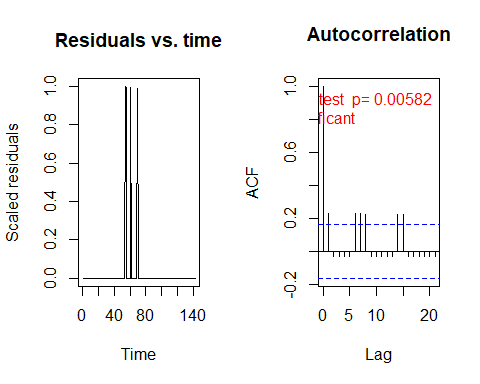

```
##
##  Durbin-Watson test
##
## data:  simulationOutput$scaledResiduals ~ 1
## DW = 1.5404, p-value = 0.005817
## alternative hypothesis: true autocorrelation is not 0
#

SimOut_lm2relev <- simulateResiduals(fittedModel = lm2relev, plot = T)
plot(SimOut_lm2relev)
```

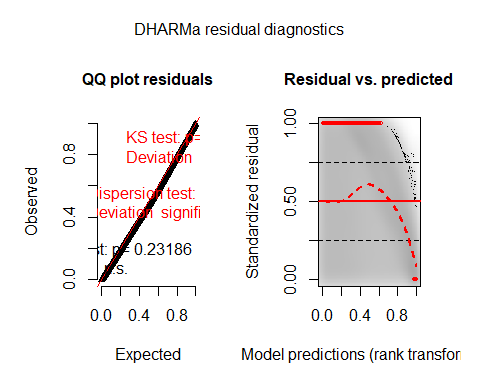

testZeroInflation(SimOut_lm2relev)

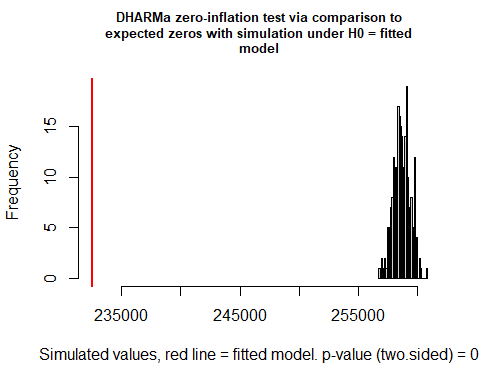

```
##
##  DHARMa zero-inflation test via comparison to expected zeros with
##  simulation under H0 = fitted model
##
## data:  simulationOutput
## ratioObsSim = 0.89895, p-value < 2.2e-16
## alternative hypothesis: two.sided
testOutliers(SimOut_lm2relev, type= 'bootstrap')
```

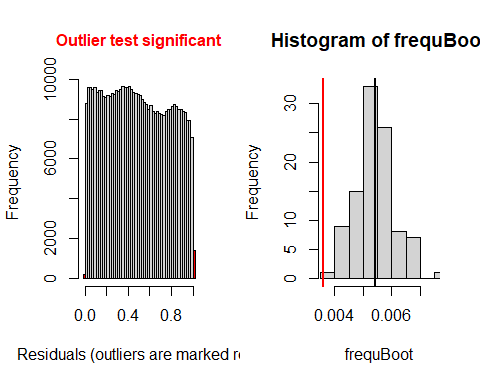

```
##
##  DHARMa bootstrapped outlier test
##
## data:  SimOut_lm2relev
## outliers at both margin(s) = 1603, observations = 446164, p-value <
## 2.2e-16
## alternative hypothesis: two.sided
##  percent confidence interval:
##  0.004377090 0.006769484
## sample estimates:
## outlier frequency (expected: 0.00541536296070503 )
##                                        0.003592849
simoutrecalc <- recalculateResiduals(SimOut_lm2relev, group = df_14$Date2)
testTemporalAutocorrelation(simoutrecalc, time = unique(df_14$Date2))
```

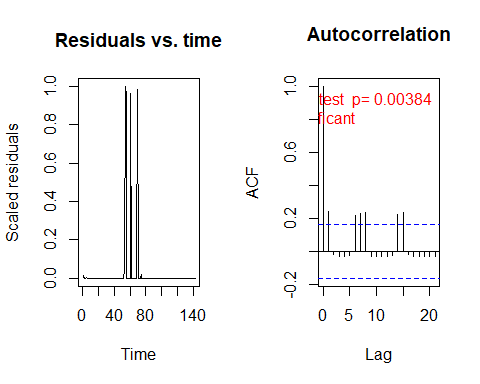

```
##
##  Durbin-Watson test
##
## data:  simulationOutput$scaledResiduals ~ 1
## DW = 1.5182, p-value = 0.003841
## alternative hypothesis: true autocorrelation is not 0
```

\#

SimOut_lm2catziprelev <- simulateResiduals(fittedModel = lm2catziprelev, plot = T)

\## DHARMa:plot used testOutliers with type = binomial for computational reasons (nObs > 500). Note that this method may not have inflated Type I error rates for integer-valued distributions. To get a more exact result, it is recommended to re-run testOutliers with type = 'bootstrap'. See ?testOutliers for details

plot(SimOut_lm2catziprelev)

\## DHARMa:plot used testOutliers with type = binomial for computational reasons (nObs > 500). Note that this method may not have inflated Type I error rates for integer-valued distributions. To get a more exact result, it is recommended to re-run testOutliers with type = 'bootstrap'. See ?testOutliers for details

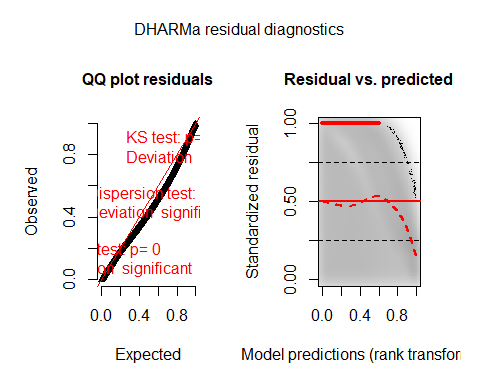

testZeroInflation(SimOut_lm2catziprelev)

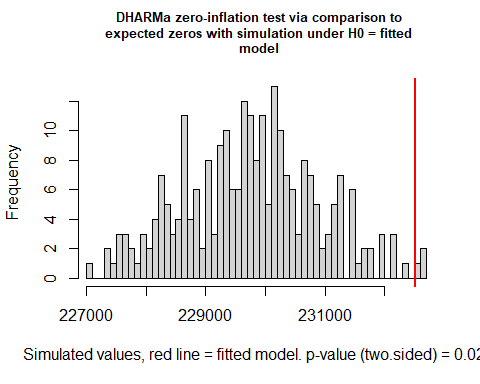

```
##
##  DHARMa zero-inflation test via comparison to expected zeros with
##  simulation under H0 = fitted model
##
## data:  simulationOutput
## ratioObsSim = 1.012, p-value = 0.024
## alternative hypothesis: two.sided
testOutliers(SimOut_lm2catziprelev, type= 'bootstrap')
```

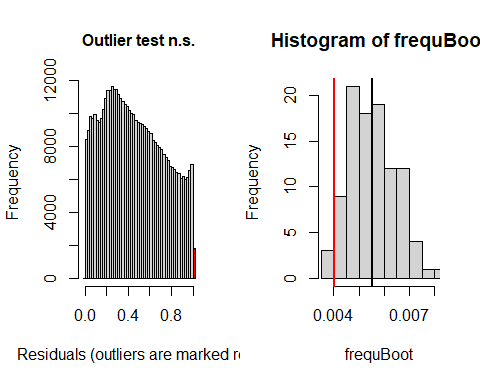

```
##
##  DHARMa bootstrapped outlier test
##
## data:  SimOut_lm2catziprelev
## outliers at both margin(s) = 1802, observations = 446164, p-value =
## 0.06
## alternative hypothesis: two.sided
##  percent confidence interval:
##  0.004060951 0.007104675
## sample estimates:
## outlier frequency (expected: 0.00551676065303341 )
##                                        0.004038874
simoutrecalc <- recalculateResiduals(SimOut_lm2catziprelev, group = df_14$Date2)
testTemporalAutocorrelation(simoutrecalc, time = unique(df_14$Date2))
```

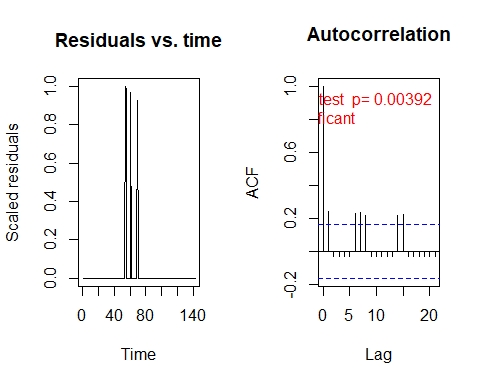

```
##
##  Durbin-Watson test
##
## data:  simulationOutput$scaledResiduals ~ 1
## DW = 1.5192, p-value = 0.003916
## alternative hypothesis: true autocorrelation is not 0
```

\###
SimOut_lm3glmmrelev <- simulateResiduals(fittedModel = lm3glmmrelev, plot = T)

\## DHARMa:plot used testOutliers with type = binomial for computational reasons (nObs > 500). Note that this method may not have inflated Type I error rates for integer-valued distributions. To get a more exact result, it is recommended to re-run testOutliers with type = 'bootstrap'. See ?testOutliers for details

plot(SimOut_lm3glmmrelev)

\## DHARMa:plot used testOutliers with type = binomial for computational reasons (nObs > 500). Note that this method may not have inflated Type I error rates for integer-valued distributions. To get a more exact result, it is recommended to re-run testOutliers with type = 'bootstrap'. See ?testOutliers for details

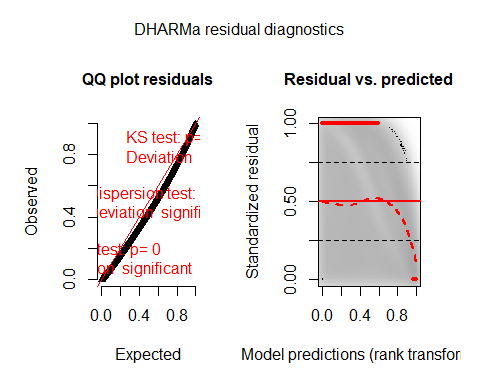

testZeroInflation(SimOut_lm3glmmrelev)

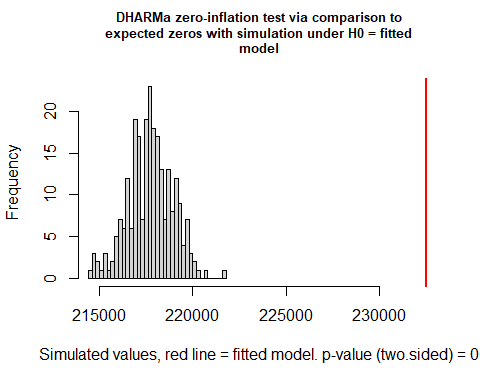

```
##
##  DHARMa zero-inflation test via comparison to expected zeros with
##  simulation under H0 = fitted model
##
## data:  simulationOutput
## ratioObsSim = 1.0679, p-value < 2.2e-16
## alternative hypothesis: two.sided
testOutliers(SimOut_lm3glmmrelev, type= 'bootstrap')
```

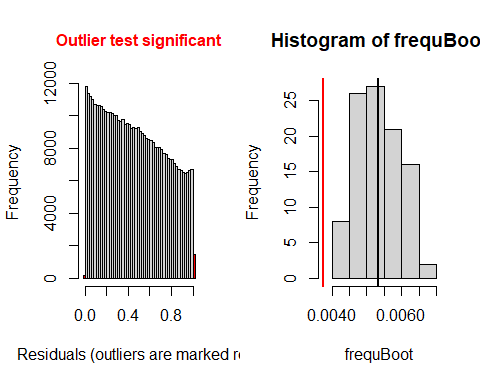

```
##
##  DHARMa bootstrapped outlier test
##
## data:  SimOut_lm3glmmrelev
## outliers at both margin(s) = 1671, observations = 446164, p-value <
## 2.2e-16
## alternative hypothesis: two.sided
##  percent confidence interval:
##  0.004189155 0.006475366
## sample estimates:
## outlier frequency (expected: 0.00534637487560628 )
##                                         0.00374526
simoutrecalc <- recalculateResiduals(SimOut_lm3glmmrelev, group = df_14$Date2)
testTemporalAutocorrelation(simoutrecalc, time = unique(df_14$Date2))
```

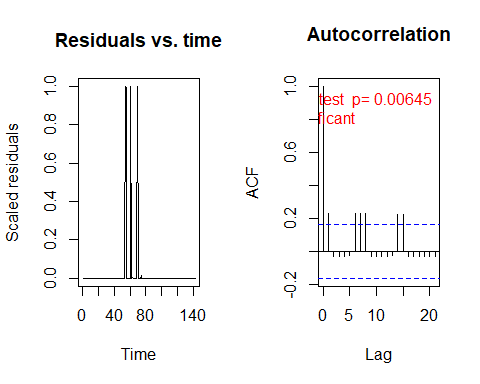

```
##
##  Durbin-Watson test
##
## data:  simulationOutput$scaledResiduals ~ 1
## DW = 1.546, p-value = 0.00645
## alternative hypothesis: true autocorrelation is not 0
```

SimOut_lm3glmmRandSlope <- simulateResiduals(fittedModel = lm3glmmRandslope, plot = T)

\## DHARMa:plot used testOutliers with type = binomial for computational reasons (nObs > 500). Note that this method may not have inflated Type I error rates for integer-valued distributions. To get a more exact result, it is recommended to re-run testOutliers with type = 'bootstrap'. See ?testOutliers for details

plot(SimOut_lm3glmmRandSlope)

\## DHARMa:plot used testOutliers with type = binomial for computational reasons (nObs > 500). Note that this method may not have inflated Type I error rates for integer-valued distributions. To get a more exact result, it is recommended to re-run testOutliers with type = 'bootstrap'. See ?testOutliers for details

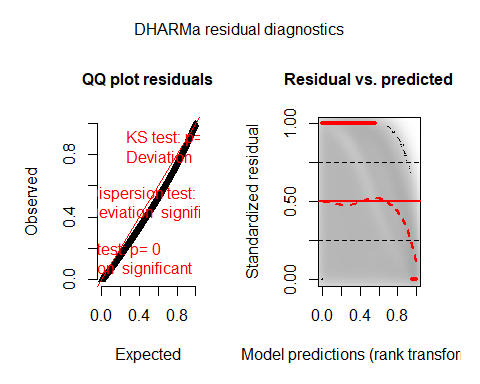

testZeroInflation(SimOut_lm3glmmRandSlope)

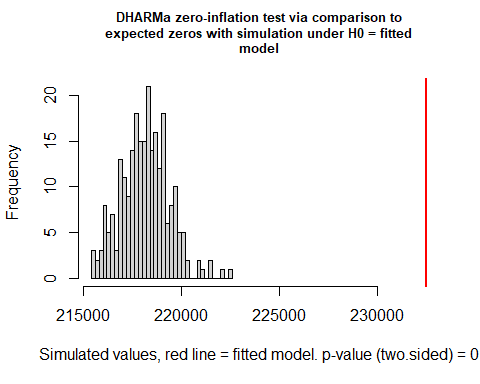

```
##
##  DHARMa zero-inflation test via comparison to expected zeros with
##  simulation under H0 = fitted model
##
## data:  simulationOutput
## ratioObsSim = 1.0657, p-value < 2.2e-16
## alternative hypothesis: two.sided
testOutliers(SimOut_lm3glmmRandSlope, type= 'bootstrap')
```

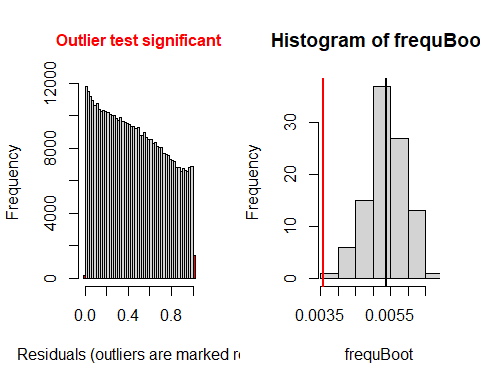

```
##
##  DHARMa bootstrapped outlier test
##
## data:  SimOut_lm3glmmRandSlope
## outliers at both margin(s) = 1593, observations = 446164, p-value <
## 2.2e-16
## alternative hypothesis: two.sided
##  percent confidence interval:
##  0.004222494 0.006330352
## sample estimates:
## outlier frequency (expected: 0.0053729122026878 )
##                                       0.003570436
simoutrecalc <- recalculateResiduals(SimOut_lm3glmmRandSlope, group = df_14$Date2)
testTemporalAutocorrelation(simoutrecalc, time = unique(df_14$Date2))
```

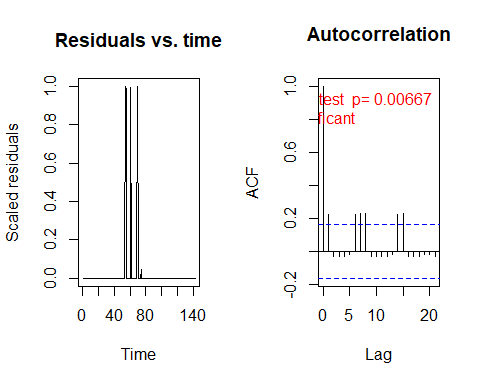

```
##
##  Durbin-Watson test
##
## data:  simulationOutput$scaledResiduals ~ 1
## DW = 1.5479, p-value = 0.006666
## alternative hypothesis: true autocorrelation is not 0
```

\##
SimOut_lm4catziprelev <- simulateResiduals(fittedModel = lm4catziprelev, plot = T)

\## DHARMa:plot used testOutliers with type = binomial for computational reasons (nObs > 500). Note that this method may not have inflated Type I error rates for integer-valued distributions. To get a more exact result, it is recommended to re-run testOutliers with type = 'bootstrap'. See ?testOutliers for details

plot(SimOut_lm4catziprelev)

\## DHARMa:plot used testOutliers with type = binomial for computational reasons (nObs > 500). Note that this method may not have inflated Type I error rates for integer-valued distributions. To get a more exact result, it is recommended to re-run testOutliers with type = 'bootstrap'. See ?testOutliers for details

testZeroInflation(SimOut_lm4catziprelev)

```
##
##  DHARMa zero-inflation test via comparison to expected zeros with
##  simulation under H0 = fitted model
##
## data:  simulationOutput
## ratioObsSim = 1.0509, p-value < 2.2e-16
## alternative hypothesis: two.sided
testOutliers(SimOut_lm4catziprelev, type= 'bootstrap')
```

```
##
##  DHARMa bootstrapped outlier test
##
## data:  SimOut_lm4catziprelev
## outliers at both margin(s) = 1520, observations = 446164, p-value <
## 2.2e-16
## alternative hypothesis: two.sided
##  percent confidence interval:
##  0.004346216 0.006524507
## sample estimates:
## outlier frequency (expected: 0.00542166109323029 )
##                                        0.003406819
simoutrecalc <- recalculateResiduals(SimOut_lm4catziprelev, group = df_14$Date2)
testTemporalAutocorrelation(simoutrecalc, time = unique(df_14$Date2))
```

```
##
##  Durbin-Watson test
##
## data:  simulationOutput$scaledResiduals ~ 1
## DW = 1.5436, p-value = 0.006167
## alternative hypothesis: true autocorrelation is not 0
# REMOVING OUTLIERS LM3GLMM
r <- which(residuals(SimOut_lm3glmmrelev) == 1 | residuals(SimOut_lm3glmmrelev) == 0)

df_14$row <- c(1:446164)

`%notin%` <- Negate(`%in%`)
#removing the counties
outcount <- df_14$c_FIPS[df_14$row %in% r]
outcount <- unique(outcount)
df_14outremcount <- df_14[df_14$c_FIPS %notin% outcount,]


# REMOVING OUTLIERS LM3GLMM
summary(lm3glmmrelevoutcount)
##  Family: nbinom2  ( log )
## Formula:
## newcase_nst_14 ~ offset(popoff) + URBinary * c_daterelev + URBinary *
##     Date2 + URBinary * dsahcarried + URBinary * asahcarried +      (1 | c_FIPS)
## Data: df_14outremcount
##
##       AIC       BIC    logLik  deviance  df.resid
## 1225932.8 1226085.3 -612952.4 1225904.8    397018
##
## Random effects:
##
## Conditional model:
##  Groups Name        Variance Std.Dev.
##  c_FIPS (Intercept) 0.8733   0.9345
## Number of obs: 397032, groups:  c_FIPS, 2796
##
## Overdispersion parameter for nbinom2 family (): 2.11
##
## Conditional model:
##                                  Estimate Std. Error z value Pr(>|z|)
## (Intercept)                    -2.0656642  0.0253779  -81.40  < 2e-16 ***
## URBinary                       -2.3324511  0.0461256  -50.57  < 2e-16 ***
## c_daterelevafter SaH            0.4821471  0.0140357   34.35  < 2e-16 ***
## c_daterelevduring SaH           0.3030943  0.0105631   28.69  < 2e-16 ***
## Date2                           0.0507715  0.0001879  270.14  < 2e-16 ***
## dsahcarried                    -0.0094679  0.0002757  -34.34  < 2e-16 ***
## asahcarried                    -0.0247484  0.0003665  -67.52  < 2e-16 ***
## URBinary:c_daterelevafter SaH  -0.1981276  0.0236522   -8.38  < 2e-16 ***
## URBinary:c_daterelevduring SaH -0.1055584  0.0179513   -5.88  4.1e-09 ***
## URBinary:Date2                  0.0095086  0.0004584   20.74  < 2e-16 ***
## URBinary:dsahcarried           -0.0095222  0.0005523  -17.24  < 2e-16 ***
## URBinary:asahcarried           -0.0085088  0.0007165  -11.88  < 2e-16 ***
## ---
## Signif. codes:  0 '***' 0.001 '**' 0.01 '*' 0.05 '.' 0.1 ' ' 1
SimOut_lm3glmmrelevoutcount <- simulateResiduals(fittedModel = lm3glmmrelevoutcount, plot = T)
## DHARMa:plot used testOutliers with type = binomial for computational reasons (nObs > 500). Note that this method may not have inflated Type I error rates for integer-valued distributions. To get a more exact result, it is recommended to re-run testOutliers with type = 'bootstrap'. See ?testOutliers for details
plot(SimOut_lm3glmmrelevoutcount)
## DHARMa:plot used testOutliers with type = binomial for computational reasons (nObs > 500). Note that this method may not have inflated Type I error rates for integer-valued distributions. To get a more exact result, it is recommended to re-run testOutliers with type = 'bootstrap'. See ?testOutliers for details
```

testZeroInflation(SimOut_lm3glmmrelevoutcount)

```
##
##  DHARMa zero-inflation test via comparison to expected zeros with
##  simulation under H0 = fitted model
##
## data:  simulationOutput
## ratioObsSim = 1.0698, p-value < 2.2e-16
## alternative hypothesis: two.sided
testOutliers(SimOut_lm3glmmrelevoutcount, type= 'bootstrap')
```

```
##
##  DHARMa bootstrapped outlier test
##
## data:  SimOut_lm3glmmrelevoutcount
## outliers at both margin(s) = 851, observations = 397032, p-value <
## 2.2e-16
## alternative hypothesis: two.sided
##  percent confidence interval:
##  0.004032987 0.006420137
## sample estimates:
## outlier frequency (expected: 0.00502468314896532 )
##                                        0.002143404
simoutrecalc <- recalculateResiduals(SimOut_lm3glmmrelevoutcount, group = df_14outremcount$Date2)
testTemporalAutocorrelation(simoutrecalc, time = unique(df_14outremcount$Date2))
```

```
##
##  Durbin-Watson test
##
## data:  simulationOutput$scaledResiduals ~ 1
## DW = 1.6828, p-value = 0.05699
## alternative hypothesis: true autocorrelation is not 0
# REMOVING OUTLIERS RANDOM SLOPE
r <- which(residuals(SimOut_lm3glmmRandSlope) == 1 | residuals(SimOut_lm3glmmRandSlope) == 0)

df_14$row <- c(1:446164)

`%notin%` <- Negate(`%in%`)
#removing the counties
outcount <- df_14$c_FIPS[df_14$row %in% r]
outcount <- unique(outcount)
df_14outremcountrand <- df_14[df_14$c_FIPS %notin% outcount,]


# REMOVING OUTLIERS RANDOM SLOPE
summary(lm3glmmrelevoutcountrandslope)
##  Family: nbinom2  ( log )
## Formula:
## newcase_nst_14 ~ offset(popoff) + URBinary * c_daterelev + URBinary *
##     Date2 + URBinary * dsahcarried + URBinary * asahcarried +      (1 | c_FIPS)
## Data: df_14outremcountrand
##
##      AIC      BIC   logLik deviance df.resid
##  1220576  1220728  -610274  1220548   395314
##
## Random effects:
##
## Conditional model:
##  Groups Name        Variance Std.Dev.
##  c_FIPS (Intercept) 0.8801   0.9382
## Number of obs: 395328, groups:  c_FIPS, 2784
##
## Overdispersion parameter for nbinom2 family (): 2.11
##
## Conditional model:
##                                  Estimate Std. Error z value Pr(>|z|)
## (Intercept)                    -2.0695923  0.0254751  -81.24  < 2e-16 ***
## URBinary                       -2.4055968  0.0467563  -51.45  < 2e-16 ***
## c_daterelevafter SaH            0.4832032  0.0140579   34.37  < 2e-16 ***
## c_daterelevduring SaH           0.3055253  0.0105775   28.88  < 2e-16 ***
## Date2                           0.0507448  0.0001880  269.98  < 2e-16 ***
## dsahcarried                    -0.0094657  0.0002753  -34.38  < 2e-16 ***
## asahcarried                    -0.0246969  0.0003676  -67.19  < 2e-16 ***
## URBinary:c_daterelevafter SaH  -0.2133346  0.0237230   -8.99  < 2e-16 ***
## URBinary:c_daterelevduring SaH -0.1267896  0.0180448   -7.03 2.12e-12 ***
## URBinary:Date2                  0.0105569  0.0004730   22.32  < 2e-16 ***
## URBinary:dsahcarried           -0.0105118  0.0005651  -18.60  < 2e-16 ***
## URBinary:asahcarried           -0.0096821  0.0007246  -13.36  < 2e-16 ***
## ---
## Signif. codes:  0 '***' 0.001 '**' 0.01 '*' 0.05 '.' 0.1 ' ' 1
SimOut_lm3glmmrelevoutcountrandslope <- simulateResiduals(fittedModel = lm3glmmrelevoutcountrandslope, plot = T)
## DHARMa:plot used testOutliers with type = binomial for computational reasons (nObs > 500). Note that this method may not have inflated Type I error rates for integer-valued distributions. To get a more exact result, it is recommended to re-run testOutliers with type = 'bootstrap'. See ?testOutliers for details
plot(SimOut_lm3glmmrelevoutcountrandslope)
## DHARMa:plot used testOutliers with type = binomial for computational reasons (nObs > 500). Note that this method may not have inflated Type I error rates for integer-valued distributions. To get a more exact result, it is recommended to re-run testOutliers with type = 'bootstrap'. See ?testOutliers for details
```

testZeroInflation(SimOut_lm3glmmrelevoutcountrandslope)

```
##
##  DHARMa zero-inflation test via comparison to expected zeros with
##  simulation under H0 = fitted model
##
## data:  simulationOutput
## ratioObsSim = 1.0694, p-value < 2.2e-16
## alternative hypothesis: two.sided
testOutliers(SimOut_lm3glmmrelevoutcountrandslope, type= 'bootstrap')
```

```
##
##  DHARMa bootstrapped outlier test
##
## data:  SimOut_lm3glmmrelevoutcountrandslope
## outliers at both margin(s) = 882, observations = 395328, p-value <
## 2.2e-16
## alternative hypothesis: two.sided
##  percent confidence interval:
##  0.003981947 0.006070908
## sample estimates:
## outlier frequency (expected: 0.00499182956937024 )
##                                        0.002231059
simoutrecalc <- recalculateResiduals(SimOut_lm3glmmrelevoutcountrandslope, group = df_14outremcountrand$Date2)
testTemporalAutocorrelation(simoutrecalc, time = unique(df_14outremcountrand$Date2))
```

```
##
##  Durbin-Watson test
##
## data:  simulationOutput$scaledResiduals ~ 1
## DW = 1.6755, p-value = 0.05152
## alternative hypothesis: true autocorrelation is not 0
df_sah <- data.frame(cbind(df_14$FIPS, df_14$Date2, df_14$URBinary, df_14$daysSaH))
df_sahs <- df_sah[df_sah$X2 == 1,]
colnames(df_sahs) <- c("FIPS", "Date", "County_Type", "Days_under_SAH")
df_sahs$County_Type <- factor(df_sahs$County_Type, levels = c(0,1), labels =c("Rural", "Urban"))
wilcoxon <- wilcox.test(Days_under_SAH ~ County_Type,data= df_sahs)
wilcoxon
##
##  Wilcoxon rank sum test with continuity correction
##
## data:  Days_under_SAH by County_Type
## W = 897959, p-value < 2.2e-16
## alternative hypothesis: true location shift is not equal to 0
```

### Mobility Data Analysis

Community Mobility Reports from Google Inc^4^ were used to examine county-level mobility trends .The data shows movement trends by individuals within U.S. counties across several categories of places as well as the percent change of movement relative to a baseline period. According to Google, “The data shows how visitors to (or time spent in) categorized places change compared to our baseline days. A baseline day represents a *normal* value for that day of the week. The baseline day is the median value from the 5‑week period Jan 3 – Feb 6, 2020.” The categories of places include grocery & pharmacy, parks, transit stations, retail & recreation, residential, and workplaces. However, due to the fact that not every county reports parks and transit stations, those were not included in our analysis.

Google did not report a change in baseline for every county for every day. However, since the measured outcome is the change from baseline for each individual county relative to itself, we were able to average the percent changes across county types (i.e. rural and urban counties). For each day, the numbers of counties included in the analysis each day by county type are shown in the table below. There are a total of 1,976 rural and 1,166 urban counties in the United States.

| Date | Rural Counties | Urban Counties |
| --- | --- | --- |
| 2/15/20 | 1450 | 1111 |
| 2/16/20 | 1355 | 1098 |
| 2/17/20 | 1594 | 1150 |
| 2/18/20 | 1577 | 1146 |
| 2/19/20 | 1583 | 1146 |
| 2/20/20 | 1580 | 1147 |
| 2/21/20 | 1567 | 1141 |
| 2/22/20 | 1449 | 1113 |
| 2/23/20 | 1352 | 1096 |
| 2/24/20 | 1564 | 1142 |
| 2/25/20 | 1579 | 1145 |
| 2/26/20 | 1574 | 1146 |
| 2/27/20 | 1573 | 1146 |
| 2/28/20 | 1562 | 1141 |
| 2/29/20 | 1442 | 1109 |
| 3/1/20 | 1332 | 1090 |
| 3/2/20 | 1559 | 1142 |
| 3/3/20 | 1572 | 1146 |
| 3/4/20 | 1576 | 1146 |
| 3/5/20 | 1572 | 1146 |
| 3/6/20 | 1566 | 1142 |
| 3/7/20 | 1437 | 1109 |
| 3/8/20 | 1337 | 1092 |
| 3/9/20 | 1560 | 1143 |
| 3/10/20 | 1570 | 1146 |
| 3/11/20 | 1577 | 1147 |
| 3/12/20 | 1575 | 1146 |
| 3/13/20 | 1568 | 1142 |
| 3/14/20 | 1442 | 1109 |
| 3/15/20 | 1338 | 1090 |
| 3/16/20 | 1575 | 1146 |
| 3/17/20 | 1603 | 1151 |
| 3/18/20 | 1612 | 1151 |
| 3/19/20 | 1611 | 1152 |
| 3/20/20 | 1607 | 1150 |
| 3/21/20 | 1460 | 1110 |
| 3/22/20 | 1375 | 1102 |
| 3/23/20 | 1612 | 1151 |
| 3/24/20 | 1623 | 1152 |
| 3/25/20 | 1625 | 1152 |
| 3/26/20 | 1629 | 1152 |
| 3/27/20 | 1609 | 1150 |
| 3/28/20 | 1470 | 1116 |
| 3/29/20 | 1387 | 1102 |
| 3/30/20 | 1618 | 1152 |
| 3/31/20 | 1632 | 1152 |
| 4/1/20 | 1630 | 1152 |
| 4/2/20 | 1637 | 1152 |
| 4/3/20 | 1617 | 1151 |
| 4/4/20 | 1471 | 1115 |
| 4/5/20 | 1395 | 1102 |
| 4/6/20 | 1607 | 1152 |
| 4/7/20 | 1628 | 1152 |
| 4/8/20 | 1627 | 1152 |
| 4/9/20 | 1628 | 1153 |
| 4/10/20 | 1606 | 1152 |
| 4/11/20 | 1177 | 1075 |
| 4/12/20 | 1124 | 1061 |
| 4/13/20 | 1609 | 1152 |
| 4/14/20 | 1621 | 1152 |
| 4/15/20 | 1624 | 1152 |
| 4/16/20 | 1620 | 1152 |
| 4/17/20 | 1587 | 1151 |
| 4/18/20 | 1165 | 1072 |
| 4/19/20 | 1087 | 1054 |
| 4/20/20 | 1604 | 1152 |
| 4/21/20 | 1616 | 1152 |
| 4/22/20 | 1622 | 1152 |
| 4/23/20 | 1619 | 1152 |
| 4/24/20 | 1586 | 1151 |
| 4/25/20 | 1156 | 1072 |
| 4/26/20 | 1078 | 1051 |
| 4/27/20 | 1597 | 1151 |
| 4/28/20 | 1616 | 1152 |
| 4/29/20 | 1621 | 1152 |
| 4/30/20 | 1613 | 1152 |
| 5/1/20 | 1581 | 1151 |
| 5/2/20 | 1137 | 1069 |
| 5/3/20 | 1067 | 1046 |
| 5/4/20 | 1593 | 1152 |
| 5/5/20 | 1613 | 1152 |
| 5/6/20 | 1610 | 1152 |
| 5/7/20 | 1607 | 1152 |
| 5/8/20 | 1579 | 1151 |
| 5/9/20 | 1139 | 1067 |
| 5/10/20 | 1055 | 1044 |
| 5/11/20 | 1589 | 1151 |
| 5/12/20 | 1611 | 1152 |
| 5/13/20 | 1606 | 1152 |
| 5/14/20 | 1603 | 1152 |
| 5/15/20 | 1574 | 1150 |
| 5/16/20 | 1135 | 1067 |
| 5/17/20 | 1058 | 1042 |
| 5/18/20 | 1581 | 1152 |
| 5/19/20 | 1605 | 1152 |
| 5/20/20 | 1608 | 1152 |
| 5/21/20 | 1602 | 1152 |
| 5/22/20 | 1574 | 1151 |
| 5/23/20 | 1126 | 1062 |
| 5/24/20 | 1052 | 1042 |
| 5/25/20 | 1610 | 1146 |
| 5/26/20 | 1607 | 1151 |
| 5/27/20 | 1605 | 1152 |
| 5/28/20 | 1597 | 1152 |
| 5/29/20 | 1567 | 1149 |
| 5/30/20 | 1111 | 1054 |
| 5/31/20 | 1015 | 1028 |
| 6/1/20 | 1577 | 1152 |
| 6/2/20 | 1604 | 1152 |
| 6/3/20 | 1602 | 1152 |
| 6/4/20 | 1596 | 1152 |
| 6/5/20 | 1567 | 1148 |
| 6/6/20 | 1112 | 1055 |
| 6/7/20 | 1020 | 1036 |
| 6/8/20 | 1580 | 1151 |
| 6/9/20 | 1605 | 1152 |
| 6/10/20 | 1599 | 1152 |
| 6/11/20 | 1597 | 1152 |
| 6/12/20 | 1571 | 1148 |
| 6/13/20 | 1102 | 1055 |
| 6/14/20 | 1012 | 1031 |

```
#information on the number of counties on which data was collected
counties <- NA
for (i in 1:130) {
  counties[i] <- sum(mob$X_FREQ_[mob$date2==i])
}
#Urban and rural combined
mean(counties)
## [1] 2615.846
median(counties)
## [1] 2729
#UR separated
mean(mob$X_FREQ_[mob$URBinary=="Urban"])
## [1] 1127.9
median(mob$X_FREQ_[mob$URBinary=="Rural"])
## [1] 1579.5
mean(mob$X_FREQ_[mob$URBinary=="Urban"])
## [1] 1127.9
median(mob$X_FREQ_[mob$URBinary=="Rural"])
## [1] 1579.5
```

Repeated measures ANOVA analysis was performed on the Google mobility data using the rstatix package.^5^ The dependent variable was the mean % change from baseline mobility on a given day (mean of the counties with data on a given day). The “subjects” were the individual days and the “within-subject factor” was the county type (urban or rural). This approach was chosen because each outcome is the change from baseline (each county acts as its own control and null hypothesis that all change equally) and thus minimizes the bias of treating outcomes of rural and urban counties on the same day as independent.

Each category of mobility data were tested for outliers and normality. The anova_test function of the rstatix package tests for sphericity and automatically applies the Greenhouse-Geisser sphericity correction.

Outliers were classified as observations outside of 1.5 times the interquartile range (IQR) of their respective distribution (mobility type and rurality). Grocery/pharmacy and workplace were the only categories with outliers, with 8 outliers (4 days) and 2 outliers (1 day) removed for these categories, respectively. Below are the boxplots of every mobility category by type of county.

out3 <- boxplot(mob$work ~ mob$URBinary )$out

The assumption of normality in the case of this analysis is a given based on the large number of observations, 130 days for each of the mobility types. To ensure that this was not incorrectly assumed normality was assessed by county type and mobility type using QQ-plots. Based on these QQ-plots, residential and work seem to not be perfectly normally distributed, but there are a large number of observations (n>50) thus alleviating this concern. Below are the QQ-plots.

```

ret.aov <- anova_test(data = mob_ret, dv = retail_rec, wid =date2 , within = URBinary)
## ANOVA Table (type III tests) Retail and Recreation
##
##     Effect DFn DFd       F        p p<.05   ges
## 1 URBinary   1 129 415.405 3.71e-42     * 0.038
groc.aov <- anova_test(data = mob_groc, dv = groc_pha, wid =date2 , within = URBinary)
## ANOVA Table (type III tests) Grocery and Pharmacy
##
##     Effect DFn DFd       F        p p<.05   ges
## 1 URBinary   1 125 317.158 4.28e-36     * 0.072
work.aov <- anova_test(data = mob_work, dv = work, wid =date2 , within = URBinary)
## ANOVA Table (type III tests) Work
##
##     Effect DFn DFd       F       p p<.05   ges
## 1 URBinary   1 128 340.928 6.7e-38     * 0.035
res.aov <- anova_test(data = mob_res, dv = residential, wid =date2 , within = URBinary)
get_anova_table(res.aov)
## ANOVA Table (type III tests) Residential
##
##     Effect DFn DFd       F        p p<.05   ges
## 1 URBinary   1 129 381.282 2.44e-40     * 0.042
```

All of the repeated measures ANOVA tests resulted in a significant p-value indicating that all of the percentage change in mobility from baseline categories are statistically significantly different between Rural and Urban counties.

### Stay-at-Home Orders Start and End Dates

Individual state governments started stay-at-home at different times and ended at different times, ascertained by review of each state’s executive order by the study team. Four states (Arkansas, Iowa, North Dakota, and South Dakota) did not issue stay at home orders. Three others (Oklahoma, Utah, and Wyoming) allowed the county and local governments to make such determinations. The following table displays the start and end dates of statewide stay-at-home orders, while the subsequent table displays that of locales.

| State | Start | End |
| --- | --- | --- |
| Alabama | 4/4/20 | 4/30/20 |
| Alaska | 3/28/20 | 4/24/20 |
| Arizona | 3/31/20 | 5/15/20 |
| Arkansas | Did Not Issue SAH | |
| California | 3/19/20 | Ongoing |
| Colorado | 3/26/20 | 4/26/20 |
| Connecticut | 3/23/20 | 5/20/20 |
| Delaware | 3/24/20 | 5/31/20 |
| District of Columbia | 4/1/20 | 5/29/20 |
| Florida | 4/3/20 | 5/4/20 |
| Georgia | 4/3/20 | 4/30/20 |
| Hawaii | 3/25/20 | 5/31/20 |
| Idaho | 3/25/20 | 4/30/20 |
| Illinois | 3/21/20 | 5/29/20 |
| Indiana | 3/24/20 | 5/4/20 |
| Iowa | Did Not Issue SAH | |
| Kansas | 3/30/20 | 5/3/20 |
| Kentucky | 3/26/20 | Ongoing |
| Louisiana | 3/23/20 | 5/15/20 |
| Maine | 4/2/20 | 5/31/20 |
| Maryland | 3/30/20 | 5/15/20 |
| Massachusetts | 3/24/20 | 5/18/20 |
| Michigan | 3/24/20 | 6/1/20 |
| Minnesota | 3/27/20 | 5/13/20 |
| Mississippi | 4/3/20 | 4/27/20 |
| Missouri | 4/6/20 | 5/3/20 |
| Montana | 3/28/20 | 4/26/20 |
| Nebraska | Did Not Issue SAH | |
| Nevada | 4/1/20 | 4/29/20 |
| New Hampshire | 3/27/20 | Ongoing |
| New Jersey | 3/21/20 | 6/9/20 |
| New Mexico | 3/24/20 | 5/31/20 |
| New York | 3/22/20 | 5/28/20 |
| North Carolina | 3/30/20 | 5/22/20 |
| North Dakota | Did Not Issue SAH | |
| Ohio | 3/23/20 | 5/29/20 |
| Oklahoma | Local Decision | |
| Oregon | 3/23/20 | Ongoing |
| Pennsylvania | 4/1/20 | 6/4/20 |
| Rhode Island | 3/28/20 | 5/8/20 |
| South Carolina | 4/7/20 | 5/4/20 |
| South Dakota | Did Not Issue SAH | |
| Tennessee | 3/31/20 | 4/30/20 |
| Texas | 4/2/20 | 4/30/20 |
| Utah | Local Decision | |
| Vermont | 3/25/20 | 5/10/20 |
| Virginia | 3/30/20 | 6/10/20 |
| Washington | 3/23/20 | 5/31/20 |
| West Virginia | 3/24/20 | 5/3/20 |
| Wisconsin | 3/25/20 | 5/13/20 |
| Wyoming | Local Decision | |

| County | State | FIPS | Start | End |
| --- | --- | --- | --- | --- |
| Carter County | OK | 40019 | 4/6/20 | 4/24/20 |
| Rogers County | OK | 40131 | 4/6/20 | 4/24/20 |
| Cleveland County | OK | 40027 | 3/25/20 | 4/24/20 |
| Seqouyah County | OK | 40135 | 4/4/20 | 4/24/20 |
| Payne County | OK | 40119 | 3/30/20 | 4/24/20 |
| Tulsa County | OK | 40143 | 3/28/20 | 4/24/20 |
| Oklahoma County | OK | 40109 | 3/28/20 | 4/24/20 |
| Davis County | UT | 49011 | 4/1/20 | 5/1/20 |
| Salt Lake County | UT | 49035 | 3/30/20 | 5/1/20 |
| Summit County | UT | 49043 | 3/27/20 | 5/1/20 |
| Teton County | WY | 56039 | 3/28/20 | 5/1/20 |
